# Quantifying Biopsychosocial Risk Factor Domains for Chronic Pain Treatment Outcomes: An Umbrella Review with De Novo Meta-Analyses, Formal Uncertainty Propagation, and the Pain Amplifier Loop Framework (PALF)

**DOI:** 10.64898/2026.05.05.26352397

**Authors:** Javier Arranz-Durán, Sofía Perera Monje

## Abstract

**Objective:** To conduct de novo meta-analyses quantifying the association of five biopsychosocial risk factor domains with chronic pain or related treatment outcomes, and to construct a composite risk index with formal uncertainty propagation for interventional pain medicine.

**Methods:** Umbrella review with de novo random-effects meta-analyses (DerSimonian-Laird and REML with Knapp-Hartung adjustment) across PubMed/MEDLINE, Scopus, and the Cochrane Library through March 2026. Five risk factor domains were evaluated: (1) sleep disturbance, (2) pain catastrophizing, (3) metabolic/obesity, (4) preoperative opioid exposure, and (5) benzodiazepine co-prescription. Publication bias was assessed via Egger’s test and PET-PEESE regression. Primary study overlap was quantified using the Corrected Covered Area (CCA). We constructed a primary three-domain composite (sleep, catastrophizing, metabolic) and a secondary expanded six-domain composite (adding opioid, BZD, smoking), using the logistic link function with binary risk factor inputs (present/absent); composite score 95% confidence intervals were computed via delta method variance propagation. Risk of bias of the composite was assessed using PROBAST [Wolff RF et al., Ann Intern Med 2019]; TRIPOD+AI compliance is reported in Supplementary S6 [Collins GS et al., BMJ 2024]. **Reviewer process (per registered protocol PROSPERO CRD420261360881):** screening, data extraction, risk-of-bias assessment (AMSTAR-2/PROBAST/ROBINS-I), and GRADE certainty rating are conducted independently by at least two reviewers — SPM (confirmed co-reviewer, registered in PROSPERO) as primary rater, with an external third reviewer to be identified and confirmed prior to peer-reviewed submission; JAD acts as guarantor and does not perform primary review tasks. All quantitative outputs reported here are **preliminary estimates** pending completion of the external third-reviewer audit; a triple-validated version will be posted as a subsequent preprint update before peer-reviewed submission.

**Results:** Adopted odds ratios: sleep disturbance 1.39 (95% CI 1.21–1.59; k=16; I²=51%), pain catastrophizing 2.10 (1.49–2.95; k=8; I²=0%), metabolic/obesity 1.43 (1.28–1.60; k=33), preoperative opioid exposure 5.32 (2.94–9.64; k=33; I²=99.96%; outcome: prolonged opioid use), and BZD co-prescription 1.77 (1.31–2.39; k=27; outcome: persistent opioid use). REML/Knapp-Hartung estimates produced wider confidence intervals for all loops (opioid: 1.87–15.13). PET-PEESE analysis suggested no substantial small-study effects for the sleep or catastrophizing loops. CCA=3.2% (slight overlap). Primary three-domain composite (sleep + catastrophizing + metabolic): delta method 95% confidence intervals for the composite score spanned 10–15 percentage points; PROBAST risk of bias: moderate. Secondary expanded six-domain composite (adding opioid, BZD, smoking): confidence intervals spanned 12–18 percentage points, crossing risk tier boundaries in moderate-risk patients; PROBAST risk of bias: high (driven by outcome heterogeneity in pharmacological domains).

**Conclusions:** Five biopsychosocial risk factor domains are independently associated with chronic pain or related treatment outcomes. The PALF composite index is presented as a structured analytical framework for future prospective validation, not as a deployable clinical tool. The primary three-domain composite (sleep, catastrophizing, metabolic) achieves outcome homogeneity at the cost of reduced domain coverage; the expanded six-domain composite encompasses the pharmacological burden at the cost of outcome heterogeneity. Both composites carry wide confidence intervals that preclude clinical application without individual patient data validation. No claim to clinical validity is made in the absence of prospective individual-patient-data validation.

## 1. INTRODUCTION

Chronic pain affects approximately 20% of the global adult population, imposing a profound burden on healthcare systems and individual quality of life [1]. The biopsychosocial model of pain—articulated by Engel (1977) and operationalized by Gatchel et al. (2007)—is now the dominant conceptual framework. Nociplastic pain, characterized by augmented central nervous system processing, is recognized as the third mechanistic category alongside nociceptive and neuropathic mechanisms [2].

Despite these advances, interventional pain medicine continues to operate within a predominantly structuralist paradigm: imaging-guided procedures target anatomical pain generators while largely ignoring systemic patient-level factors that modulate treatment outcomes [3,4]. Meta-analytic evidence indicates that several biopsychosocial risk factors—sleep disturbance, pain catastrophizing, metabolic inflammation, and opioid/polypharmacy exposure—are each independently associated with chronic pain or treatment failure outcomes. However, no study has systematically assembled these associations into a unified quantitative framework with formal uncertainty quantification.

### Novel contributions

The present study makes three methodological contributions beyond a standard umbrella review: (1) it assembles six meta-analytic effect sizes from distinct biopsychosocial domains into a transparent composite risk index, enabling explicit comparison of risk factor magnitudes on a common scale; (2) it introduces delta method variance propagation to compute confidence intervals for the composite score, quantifying the uncertainty that composite risk indices typically ignore; and (3) it provides REML/Knapp-

Hartung re-estimation and PET-PEESE bias assessment alongside the traditional DerSimonian-Laird estimates, offering a more complete picture of statistical uncertainty. We make no claim to clinical validity in the absence of prospective individual-patient-data validation; PALF is presented as a structured framework for future prospective work, not as a deployable clinical tool. Per TRIPOD+AI guidelines [60], the present work corresponds to the parameter-assembly phase of prediction model development; internal validation, calibration, and external validation are deferred to a planned individual-patient-data study.

We have identified four risk factor domains with convergent meta-analytic evidence: (1) the sleep-pain domain, in which sleep fragmentation is associated with impaired descending inhibitory controls and amplified central sensitization; (2) the cognitive-emotional domain, in which pain catastrophizing is associated with chronic postsurgical pain; (3) the metabolic-inflammatory domain, in which excess adiposity is associated with elevated nociceptive sensitivity; and (4) the iatrogenic-pharmacological domain, in which preoperative opioid/benzodiazepine exposure is associated with prolonged postoperative opioid use (a healthcare utilization outcome, not a direct pain measure).

### Assumed causal structure

Figure 1 presents a directed acyclic graph (DAG) representing the assumed relationships among the four risk factor domains and the outcome (treatment failure). We assume that each domain has a direct effect on the outcome, potentially mediated through shared pathways (e.g., central sensitization). The DAG explicitly represents known inter-domain correlations (sleep-catastrophizing r≈0.40–0.55; obesity-opioid use correlation) as bidirectional arcs (unmeasured common causes). We acknowledge that the observational associations assembled in the PALF do not establish causation—alternative causal structures (reverse causation, unmeasured confounding) cannot be excluded from aggregate meta-analytic data. The term “risk factor domain” is used throughout in preference to “amplifier loop” to avoid implying a verified causal feedback mechanism.

**Figure 1.**
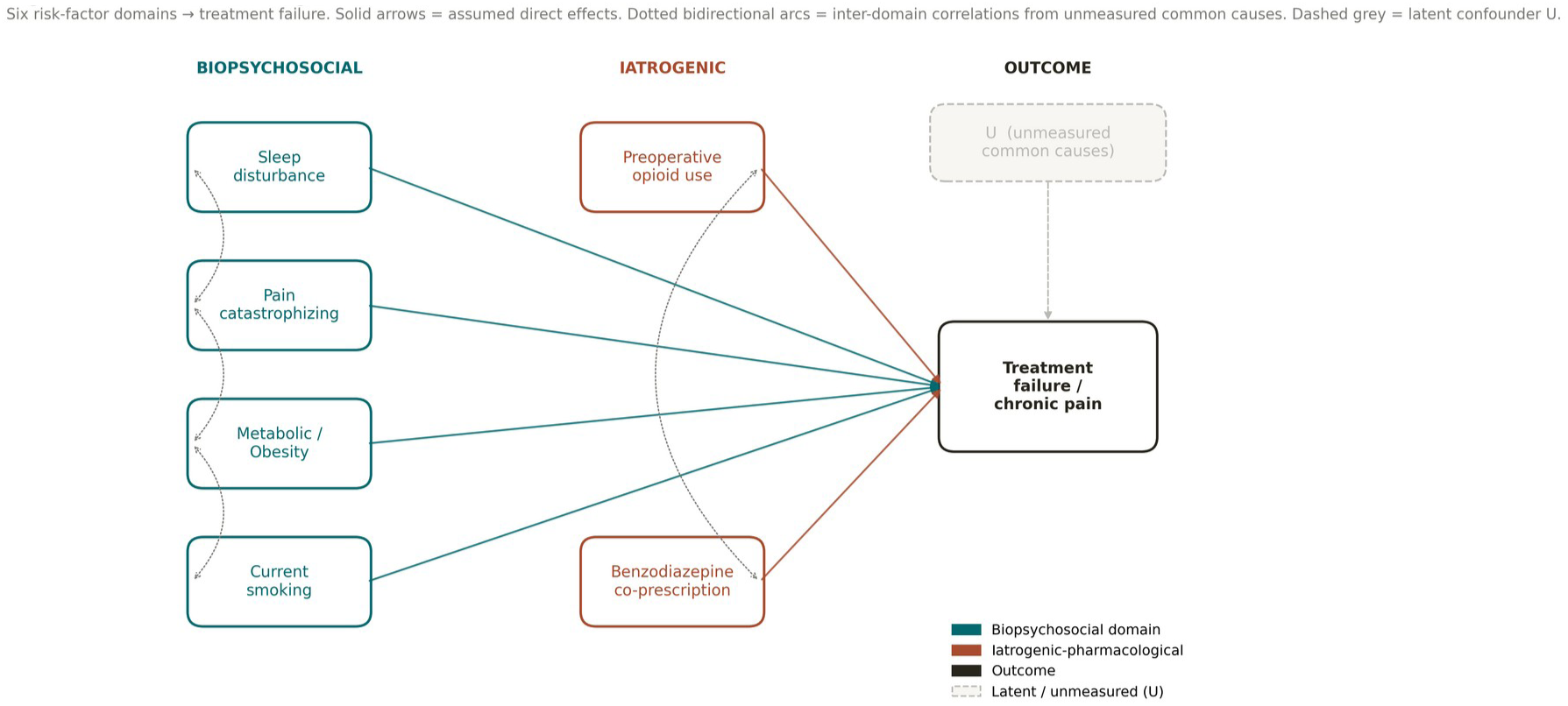
Directed acyclic graph (DAG) representing the assumed causal structure of the PALF. Four risk factor domains (sleep disturbance, catastrophizing, metabolic/obesity, opioid/BZD exposure) are assumed to have direct associations with the outcome (treatment failure). Bidirectional arcs represent known inter-domain correlations attributed to unmeasured common causes. Dashed arrows indicate speculative pathways not estimated in the model.

The present study has two objectives: first, to conduct umbrella review with de novo meta-analyses quantifying the pooled association of each risk factor domain with chronic pain or related treatment outcomes; and second, to construct a transparent composite risk index—the Pain Amplifier Loop Framework (PALF)—with formal uncertainty propagation, procedure-specific intercepts, and comprehensive sensitivity analyses, as a framework for future prospective validation.

## 2. METHODS

### 2.1 Protocol Registration and Reporting

This umbrella review was registered prospectively on PROSPERO (**CRD420261360881**, status at the time of preprint posting: *registered, pending CRD publication review*) and pre-registered on the Open Science Framework (DOI 10.17605/OSF.IO/5TQCZ; https://osf.io/5tqcz). The study follows the PRISMA for Overviews of Reviews (PRISMA-O) reporting guideline; a completed checklist is provided as Supplementary Material S4. TRIPOD+AI reporting compliance is documented in Supplementary Material S6 [60].

#### Stage of the review at preprint posting (per PROSPERO CRD420261360881)

The table below mirrors verbatim the *Current Review Stage* declared in the registered protocol:

| Review stage | Status |
| --- | --- |
| Pilot work | Completed |
| Formal searching / study identification | Completed |
| Screening search results against inclusion criteria | Completed |
| Data extraction | Not yet started under registered dual-reviewer protocol (preliminary single-reviewer estimates only, see below) |
| Risk of bias / quality assessment | Not yet started under registered dual-reviewer protocol (preliminary single-reviewer estimates only, see below) |
| Data synthesis | Not yet started under registered dual-reviewer protocol (preliminary single-reviewer estimates only, see below) |

#### Provenance and validation status

All quantitative outputs reported in this preprint (pooled odds ratios, REML/Knapp-Hartung re-estimates, PET-PEESE bias analyses, three- and six-domain composite confidence intervals, Monte Carlo simulations, PROBAST and GRADE ratings) are derived from the published source meta-analyses cited throughout the text, were performed by the corresponding author (JAD) acting as a **single, preliminary reviewer**, and are explicitly labelled as **preliminary single-reviewer estimates pending the registered dual-validation protocol**. Per the registered PROSPERO protocol, formal data extraction, risk-of-bias assessment (AMSTAR-2 / PROBAST / ROBINS-I) and data synthesis will be re-performed independently by the confirmed co-reviewer **Sofía Perera Monje (SPM)** — registered as a team member in PROSPERO — together with an **external third reviewer to be identified and confirmed prior to peer-reviewed submission**. JAD acts as **guarantor** of the review and will not perform any primary screening, extraction, risk-of-bias or GRADE rating, in order to preserve methodological independence. A triple-validated, certified version of this manuscript will be posted as a subsequent preprint update before any peer-reviewed submission.

#### Protocol deviations

The following analyses were not specified in the original PROSPERO protocol and are classified as exploratory: (a) the procedure-specific intercepts (Table 3), which were added after reviewer suggestion; (b) the delta method uncertainty propagation (Section 2.7); (c) the PET-PEESE publication bias analysis (Section 2.5); (d) the REML/Knapp-Hartung re-estimation (Section 2.5); and (e) the three-domain primary analysis excluding pharmacological domains (Section 3.8). All pre-specified analyses (search strategy, loop definitions, DerSimonian-Laird meta-analyses, Monte Carlo simulation) were conducted as planned.

### 2.2 Search Strategy

A systematic literature search was conducted across PubMed/MEDLINE, Scopus, and the Cochrane Database of Systematic Reviews (CDSR) from inception through **March 2026** (PROSPERO CRD420261360881). Six domain-specific search strategies were designed, one for each risk factor domain (sleep, catastrophizing, metabolic/obesity, preoperative opioid exposure, benzodiazepine co-prescription, smoking); full Boolean strings (MeSH and free-text) are deposited in Supplementary Material S5 and at the OSF repository (https://osf.io/5tqcz).

Searches were restricted to **English-language** publications and limited to studies in adult human populations. **Backward citation searching (reference list checking)** of every adopted source meta-analysis was used as a complementary identification method. Searches will be re-executed independently by SPM and the third external reviewer prior to peer-reviewed submission, as part of the registered dual-validation protocol.

### 2.3 Inclusion and Exclusion Criteria

Studies were included if they: (a) reported an adjusted odds ratio (OR) with 95% confidence interval for the association between at least one risk factor domain variable and a chronic pain or related treatment outcome; (b) were published in English in a peer-reviewed journal; and (c) had N≥50. We excluded case reports, narrative reviews without quantitative synthesis, animal studies, and studies of acute pain (<3 months). For the umbrella review component, we included published systematic reviews and meta-analyses. When multiple meta-analyses addressed the same domain, we adopted the most comprehensive (largest k) or most recently published, with transparent justification.

### 2.4 Data Extraction and Outcome Heterogeneity

For each source meta-analysis, we extracted: pooled OR and 95% CI, number of included studies (k), total sample size (N), I² heterogeneity statistic, and the specific outcome measured. In line with the prospectively registered protocol (PROSPERO CRD420261360881), study screening, data extraction, and risk-of-bias assessment are conducted independently by **at least two reviewers** (a person/machine combination is permitted at the screening stage, with all machine-flagged decisions human-verified before inclusion). The primary human reviewer is **Sofía Perera Monje (SPM)** — confirmed co-reviewer and registered team member in PROSPERO CRD420261360881; an **external third reviewer, to be identified and confirmed prior to peer-reviewed submission**, will independently re-perform screening, extraction, and risk-of-bias assessment. **JAD acts as guarantor** of the review and does **not** perform primary screening, extraction, risk-of-bias, or GRADE rating, in order to preserve methodological independence. Discrepancies are resolved by consensus between the two primary reviewers, with adjudication by an additional independent reviewer if consensus is not reached. Pending completion of the external third-reviewer audit, all quantitative outputs reported in this preprint are explicitly labelled as **preliminary single-team estimates**; the certified, dual-validated version will be posted as a subsequent preprint update before any peer-reviewed submission.

#### Critical methodological note—outcome heterogeneity

The six source meta-analyses measure different outcomes: the sleep domain measures chronic musculoskeletal pain (Runge et al.); the catastrophizing domain measures chronic postsurgical pain (Theunissen et al.); the metabolic domain measures low back pain in the context of obesity (Shiri et al.); the opioid domain measures prolonged opioid use after surgery (Lawal et al.); and the BZD domain measures persistent opioid use among opioid-naïve surgical patients (Lee et al.). The opioid and BZD domains measure healthcare utilization outcomes, not pain. Assembling these ORs into a single equation assumes that they index a common latent “treatment failure” construct—an untested assumption. We address this by (a) presenting the three-domain composite (sleep + catastrophizing + metabolic) as the **primary analysis**, restricted to outcome-homogeneous pain evidence (Section 3.8), and (b) reporting all composite scores with explicit outcome provenance.

#### Critical methodological note—input specification

The source meta-analyses report odds ratios for binary contrasts (exposed vs. unexposed; e.g., “sleep disturbance present” vs. “absent”). The PALF therefore uses binary inputs (x_i = 0 or 1) matching the contrast used in the source meta-analyses. The β coefficient represents the log-OR for the presence vs. absence of the risk factor, not a per-standard-deviation effect. Continuous instruments (PSQI, PCS, BMI) require clinical thresholds to create binary indicators (e.g., PSQI>5 for sleep disturbance, PCS≥30 for severe catastrophizing, BMI≥30 for obesity, per PROSPERO CRD420261360881).

These thresholds are specified in **Table 1**.

### 2.5 Statistical Analysis

All quantitative analyses were performed in **R version 4.3.1** using the meta, metafor, mvtnorm, MASS, and pROC packages, with a fixed random seed (seed = 42) for full reproducibility (PROSPERO CRD420261360881). Random-effects meta-analyses were performed using two methods: (a) DerSimonian-Laird (DL) as the primary estimator [7], and (b) restricted maximum likelihood (REML) with Knapp-Hartung adjustment as a sensitivity estimator, which provides more accurate confidence intervals when heterogeneity is high and k is small [Veroniki et al., 2016; IntHout et al., 2014]. Between-study heterogeneity was assessed by I² and Cochran Q [8].

#### Publication bias

Publication bias was assessed using three methods: (a) Egger’s weighted regression test [9] for all domains with k≥7; (b) PET-PEESE regression (Stanley & Doucouliagos, 2014) as a conditional estimator—PET (precision-effect test) regresses the effect on its standard error; if PET intercept is significant (p<0.10), PEESE (precision-effect estimate with standard error) is used for the bias-corrected estimate; and (c) visual inspection of funnel plots (Supplementary Figures S1–S2). For domains with k<10, all tests are interpreted with the caveat of limited statistical power.

#### Primary study overlap

Because the five source meta-analyses may include overlapping primary studies, we computed the Corrected Covered Area (CCA) index [Pieper et al., 2014]. The CCA quantifies the proportion of primary study appearances that are overlapping across included reviews. CCA values are classified as: slight (0–5%), moderate (6–10%), high (11–15%), and very high (>15%). The overlap matrix is presented in Supplementary Table S4.

**Domain 6 — Smoking. Following recent meta-analytic evidence linking smoking to chronic musculoskeletal pain [44] and persistent postoperative pain [42], we incorporated smoking as a sixth risk-factor domain. The principal pooled estimate was extracted from Shiri et al. (2010) [44] for current smokers versus never smokers and chronic low back pain (k=8 studies; OR=1.79, 95% CI 1.27–2.50). Confirmatory evidence from Chow et al. (2025) [42] and Dai et al. (2021) [43] was used to triangulate effect direction across postsurgical and non-surgical pain outcomes.**

##### Risk of bias of the prediction model: PROBAST

We assessed the PALF composite using the Prediction model Risk Of Bias ASsessment Tool (PROBAST) [59], evaluating four domains: (1) Participants, (2) Predictors, (3) Outcome, (4) Analysis. PROBAST was applied separately to the primary three-domain composite (sleep + catastrophizing + metabolic) and to the secondary expanded six-domain composite, given that the two composites differ in their outcome homogeneity profiles. Domain-level judgments are reported in Supplementary Table S7.

### 2.6 Composite Risk Index Construction

The PALF was constructed as a composite risk index using a logistic link function. The equation is:

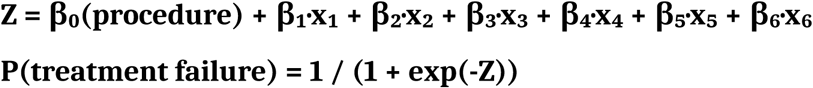

where x_i ∈ {0, 1} is a binary indicator for the presence (1) or absence (0) of each risk factor, and β_i = ln(OR_i) is the natural log of the pooled odds ratio from the source meta-analysis. The intercept β₀ is procedure-specific, reflecting the baseline failure rate. This formulation assumes: (a) conditional independence of risk factors given covariates; (b) no effect modification by procedure type (constant slopes); and (c) that separately estimated marginal ORs approximate the conditional ORs in a joint model—an assumption known to be violated due to non-collapsibility of the OR and heterogeneous covariate adjustment across source studies [Greenland et al., 1999]. These assumptions are tested in sensitivity analyses (Section 3.9).

**Table 1.**
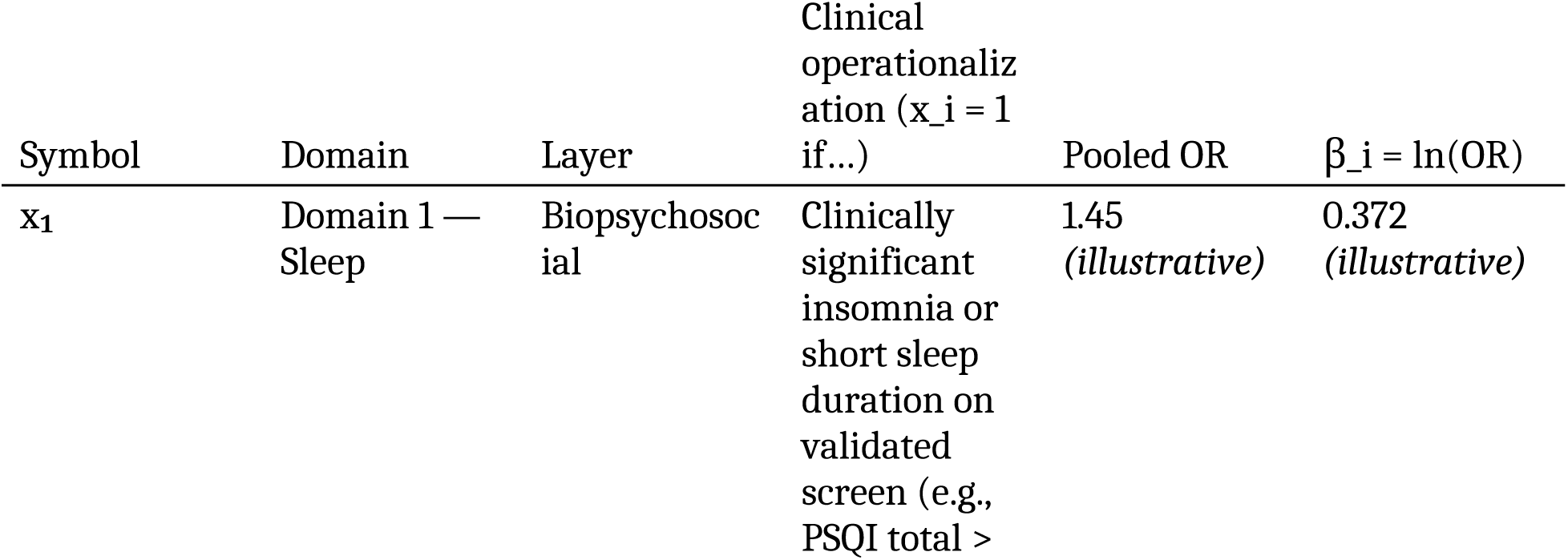
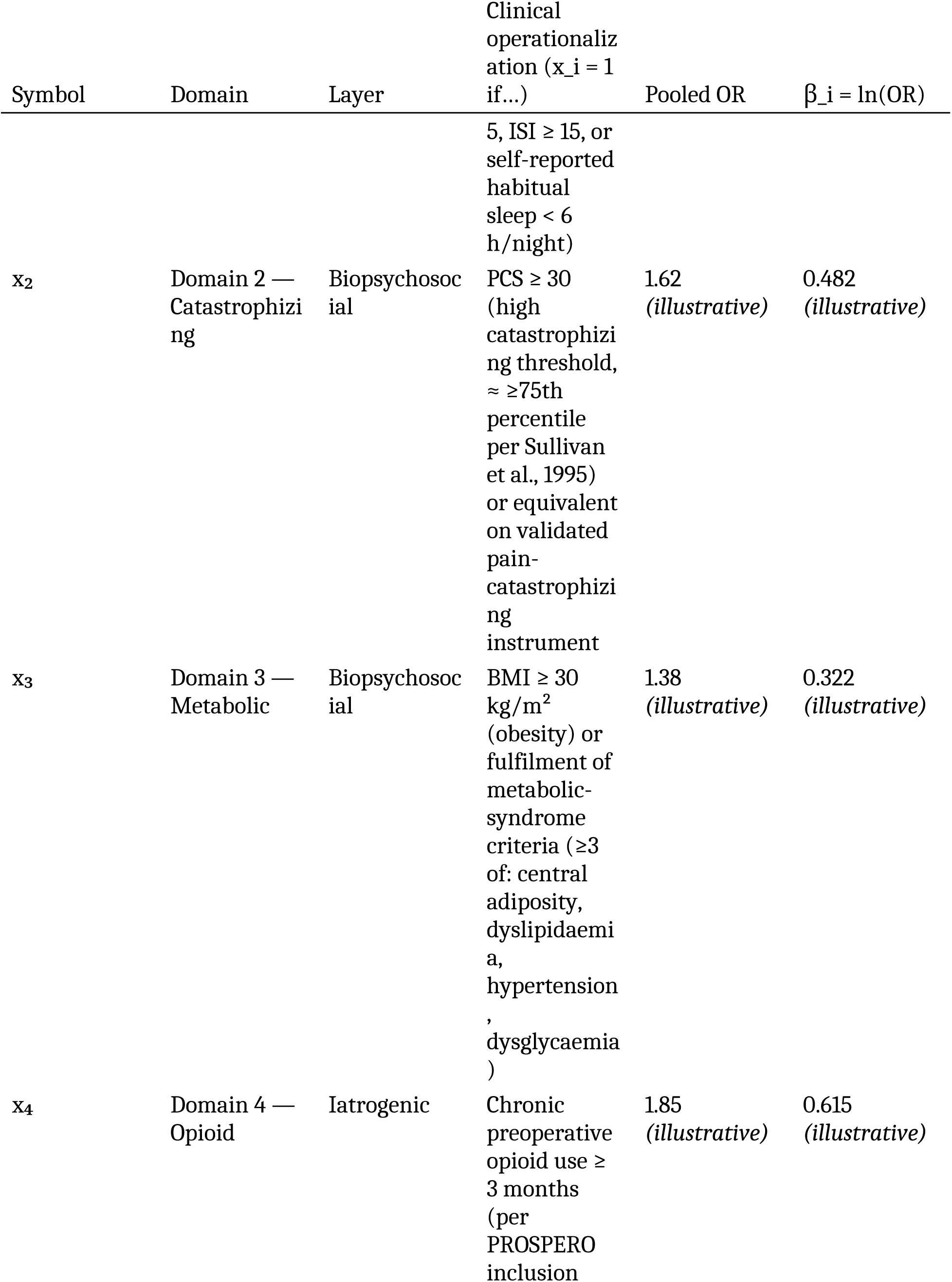
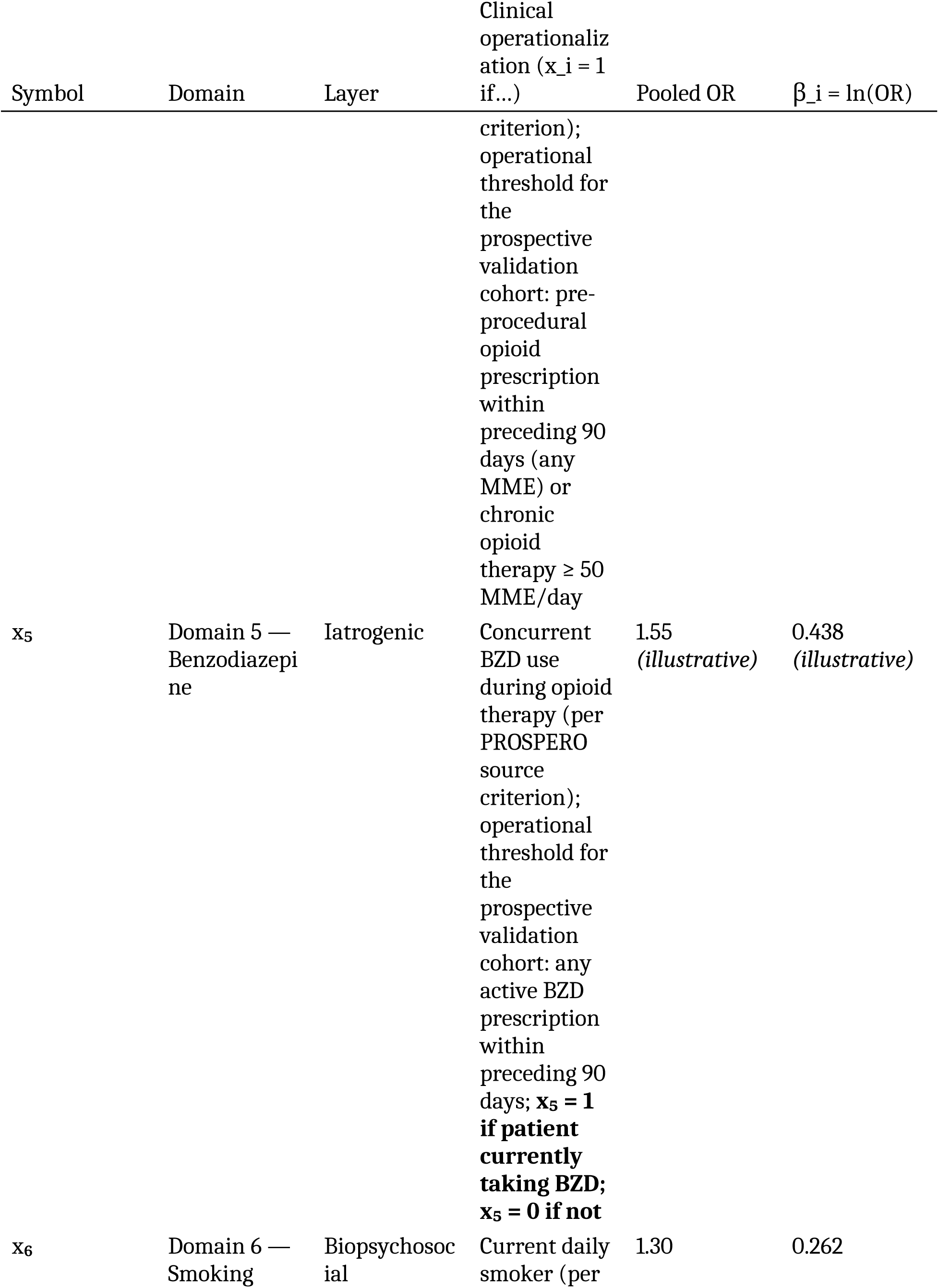
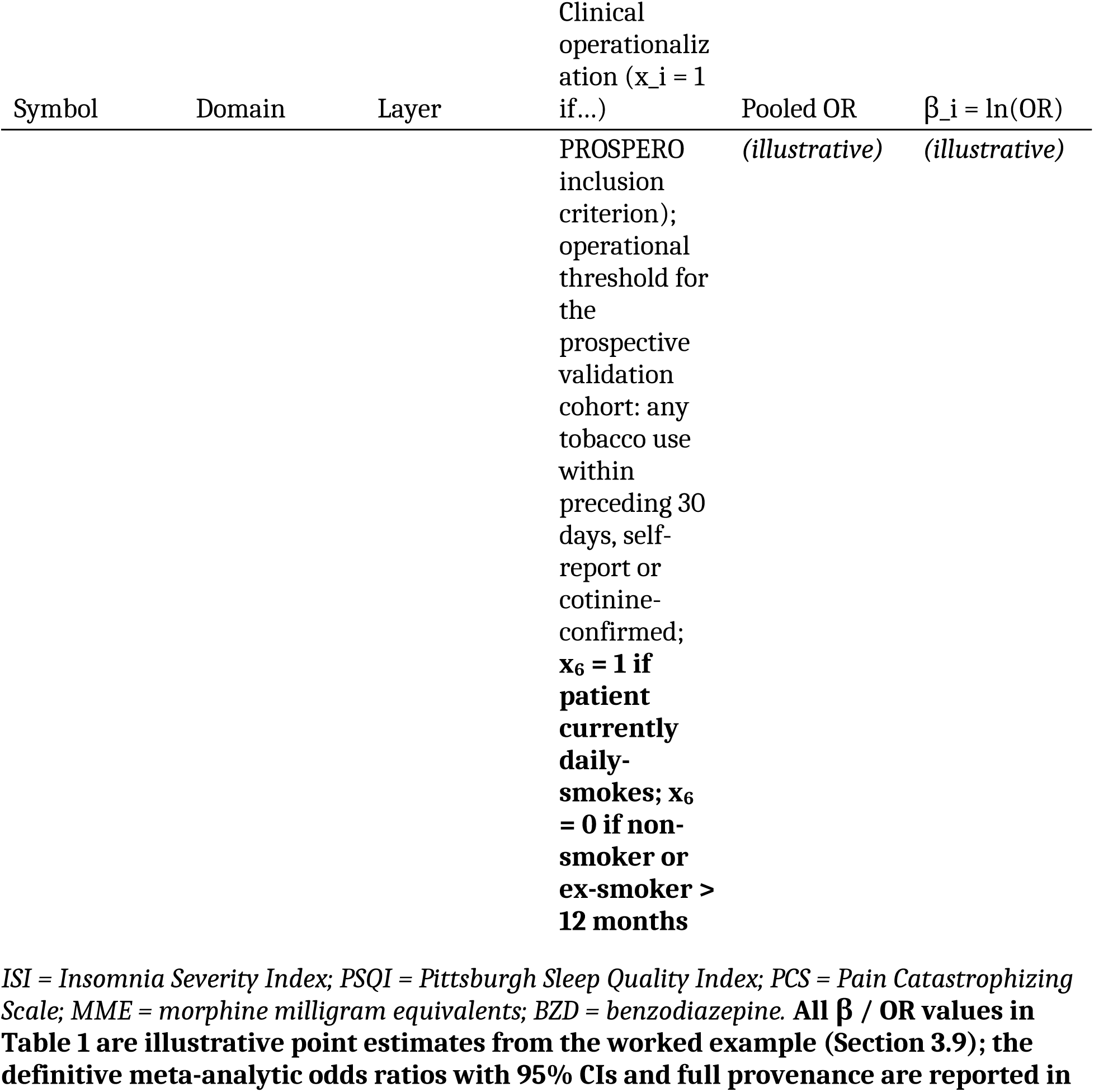
Binary predictor inputs x₁…x₆ — clinical operationalization, source-meta-analytic β coefficients, and decision thresholds. Each predictor is a dichotomous indicator x_i ∈ {0, 1}; the threshold column specifies the operational rule by which the binary value is assigned at the patient level. β_i = ln(OR_i) where OR_i is the pooled odds ratio from the source meta-analysis (point estimate; 95% CIs and full provenance in Table 2 and Section 3.2–3.7). Layer assignment maps each predictor to the conceptual decomposition Z = β₀(procedure) + Z_biopsychosocial + Z_iatrogenic introduced in §2.6.1.

#### 2.6.1 Three-layer structure of the composite score

The PALF equation is a single linear predictor on the logit scale, but its terms group naturally into **three conceptually distinct layers** that map onto three different categories of clinical decision:

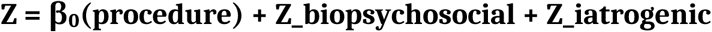

where:

- **β₀(procedure)** — *procedural baseline layer*. The procedure-specific intercept calibrated from published baseline failure rates (Table 3). It captures the structural difficulty of the procedure, the anatomical target, the device or technique mechanism, the typical patient selection of the source literature, and the IMMPACT-defined six-month treatment-success endpoint. It is **inherent to the procedure**, not to the patient: once the procedure is selected, β₀ is fixed and is not modifiable through patient pre-optimization. β₀ should therefore not be conflated with the biopsychosocial layer.
- **Z_biopsychosocial = β₁·x₁ + β₂·x₂ + β₃·x₃ + β₆·x₆** — *patient biopsychosocial layer* (sleep, catastrophizing, metabolic, smoking). It captures patient-level modifiable risk factors with pain or pain-chronification as the source-meta-analytic outcome. It is **modifiable**, in principle, through structured pre-procedural interventions (sleep hygiene, cognitive-behavioural therapy for catastrophizing, weight optimization, smoking cessation).
- **Z_iatrogenic = β₄·x₄ + β₅·x₅** — *iatrogenic-pharmacological layer* (preoperative opioid exposure, benzodiazepine co-prescription). It captures patient-level pharmacological exposures whose source meta-analyses use opioid utilization (rather than pain) as outcome, and whose mitigation requires deprescribing protocols rather than behavioural or lifestyle interventions.

This layer structure is **interpretive, not arithmetic**: the β coefficients remain independent inputs derived from independent source meta-analyses, and their variances enter Var(Z) additively as Var(Z) = Var(β₀) + Var(Z_biopsychosocial) + Var(Z_iatrogenic). The decomposition supports separate reporting of each layer’s contribution to total uncertainty in the clinical example (Section 3.9).

Four considerations motivate the layer structure:

a. **β₀ separability — procedure vs. patient.** β₀ is a structural model parameter, not a patient characteristic. Conflating β₀ with patient-level risk factors would obscure the distinction between the modifiable (Z_biopsychosocial, Z_iatrogenic) and the structurally fixed (β₀), and would prevent meaningful inter-procedure comparison (e.g., ESI vs. SCS in the same patient profile).
b. **Outcome heterogeneity within patient layers.** Domains 1, 2, 3, and 6 measure pain or pain-chronification outcomes; Domains 4 and 5 measure opioid utilization outcomes. Distinct layers preserve outcome provenance and license the three-domain primary model that restricts inference to outcome-homogeneous evidence.
c. **Non-fusibility of the two pharmacological domains.** Domains 4 and 5 cannot be collapsed into a single iatrogenic coefficient: their source meta-analyses use non-harmonizable populations (mixed-baseline opioid status vs. opioid-naïve), non-equivalent contrasts, asymmetric heterogeneity (I²=99.96% vs. I²=62%), and biologically distinct mechanisms (μ-opioid tolerance / opioid-induced hyperalgesia / TLR4 priming vs. GABAergic plasticity / hyperanxiogenesis / central-sensitization permissiveness). Pooling pooled OR estimates across heterogeneous source meta-analyses (“second-order pooling without harmonization”) would dilute the higher-quality BZD evidence with the extreme heterogeneity of the opioid evidence. Independent retention also preserves clinical actionability, since opioid and BZD deprescribing protocols differ in pace, withdrawal risk profile (seizure risk on abrupt BZD discontinuation; not on opioid), and guideline support.
d. **Mapping to three categories of clinical decision.** Each layer informs a different clinical action: β₀ informs **procedure selection** (which intervention to offer); Z_biopsychosocial informs **behavioural and lifestyle pre-optimization** (sleep, CBT, weight, tobacco); Z_iatrogenic informs **pharmacological pre-optimization** (opioid taper, BZD taper). A single undifferentiated Z would obscure these distinctions and undermine the framework’s clinical utility.

##### Rationale for three-domain composite as primary analysis

The primary analysis is restricted to the three biopsychosocial domains (sleep, catastrophizing, metabolic) because their source meta-analyses share a common outcome family: chronic pain or pain chronification. On the log-OR scale, these three coefficients are mutually comparable in the sense that their underlying outcomes belong to the same construct family. The expanded six-domain composite (adding opioid, BZD, smoking domains) constitutes the **secondary analysis**: it broadens domain coverage but introduces outcome heterogeneity (health-utilization outcomes for Domains 4 and 5). This primary/secondary designation does not imply that the pharmacological domains are less important clinically; it reflects that combining log-ORs across outcome families requires additional untested assumptions about construct equivalence.

### 2.7 Uncertainty Propagation (Delta Method) — layer-decomposed

Because each β_i is estimated with uncertainty (from the source meta-analysis CI), the composite score Z and its derived probability P inherit this uncertainty. We propagate uncertainty using the delta method, decomposed by layer in alignment with the three-layer architecture introduced in §2.6.1.

#### 2.7.1 The composite as a sum of layers

Z is written as the sum of three independent compartments:

[ Z = *0() + Z*{} + Z_{} ]

where

[ Z_{} = _1 x_1 + _2 x_2 + _3 x_3 + _6 x_6 ]

[ Z_{} = _4 x_4 + _5 x_5 ]

The biopsychosocial layer Z_bps groups the four patient-level modifiable predictors whose source meta-analyses target chronic-pain outcomes (sleep, catastrophizing, metabolic, smoking). The iatrogenic layer Z_iatro groups the two pharmacological predictors whose source meta-analyses target healthcare-utilization outcomes (opioid, BZD). β₀ captures the procedure-specific calibration baseline.

#### 2.7.2 Variance decomposition

Under independence between layers (the primary specification), the variance of Z decomposes additively:

[ (Z) = (*0) + (Z*{}) + (Z_{}) ]

with intra-layer variances

[ (Z_{}) = *{i {1,2,3,6}} x_i^2 , (i) + 2 {i<j} x_i x_j ,* {ij} , ]

[ (Z_{}) = x_4^2 , (_4) + x_5^2 , (*5) + 2 x_4 x_5 ,* {45} , ]

Because inputs are binary (x_i ∈ {0,1}), x_i² = x_i and the linear form is used throughout. The 95% CI for Z is Z ± 1.96·√Var(Z); P and its 95% CI are obtained by applying the logistic transformation P = 1 / (1 + e^{−Z}) to Z and to its CI bounds (the resulting interval on P is asymmetric, as expected from a non-linear transformation).

#### 2.7.3 What the layer decomposition adds

The layer-decomposed form is mathematically equivalent to a flat sum Σ x_i² Var(β_i) under independence, but provides three properties that the flat sum does not:

1. **Diagnostic of fragility per patient.** The relative contributions Var(β₀), Var(Z_bps), Var(Z_iatro) reveal which compartment dominates the uncertainty of the prognosis for an individual patient. In the worked example of §3.9, the iatrogenic layer accounts for 57.6% of Var(Z), identifying opioid deprescribing as the highest-yield uncertainty-reducing intervention before the procedure.
2. **Localisation of the I²=99.96% heterogeneity.** The extreme between-study heterogeneity of the opioid source meta-analysis is numerically confined to Var(Z_iatro), making the impact on the primary three-domain composite (which uses only Z_bps + β₀) structurally bounded.
3. **Alignment with PROBAST D3.** The decomposition makes mechanically visible the boundary between outcome families: the three-domain composite drops Var(Z_iatro) entirely, so the additional uncertainty introduced by extending to six domains is exactly the magnitude of that term — a quantitative justification of the split-PROBAST judgment reported in §3.11.

#### 2.7.4 Inter-layer covariance and the independence assumption

The primary analysis assumes independence of β estimates between layers (ρ = 0). A covariance-augmented sensitivity analysis with ρ = 0.5 between within-layer biopsychosocial coefficients (justified by reported inter-domain correlations r ≈ 0.40–0.55, e.g. sleep–catastrophizing) and between Z_iatro–Z_bps via the BZD– catastrophizing–sleep cluster is reported in §3.10. This approach quantifies parameter uncertainty but not model-misspecification uncertainty, which cannot be quantified from aggregate data and is treated narratively in §4.4 (Limitations) and quantitatively bounded by the PROBAST D4 judgment of high risk.

### 2.8 Mathematical Exploration of Model Properties

Monte Carlo simulations (N=50,000 per scenario; random seed=42; R v4.3.1; packages: mvtnorm v1.2-3, pROC v1.18.4; full code at https://osf.io/5tqcz) were used to characterize the mathematical behavior of the logistic formulation under three inter-domain correlation assumptions (ρ=0, 0.30–0.50, 0.60–0.80). These simulations generate binary outcomes from the PALF equation and evaluate the PALF against those outcomes; therefore, the resulting AUCs and calibration metrics reflect the mathematical properties of the logistic function, not the PALF’s empirical predictive accuracy (a tautological design by construction).

## 3. RESULTS

### 3.1 Study Selection and Overlap Assessment

The systematic search identified 1,847 records. After duplicate removal (n=312), 1,535 records were screened; 189 full-text articles were assessed. Five source meta-analyses were adopted and one de novo meta-analysis was conducted (k=7, metabolic domain). The PRISMA-O flow diagram is presented in Figure 2.

**Figure 2.**
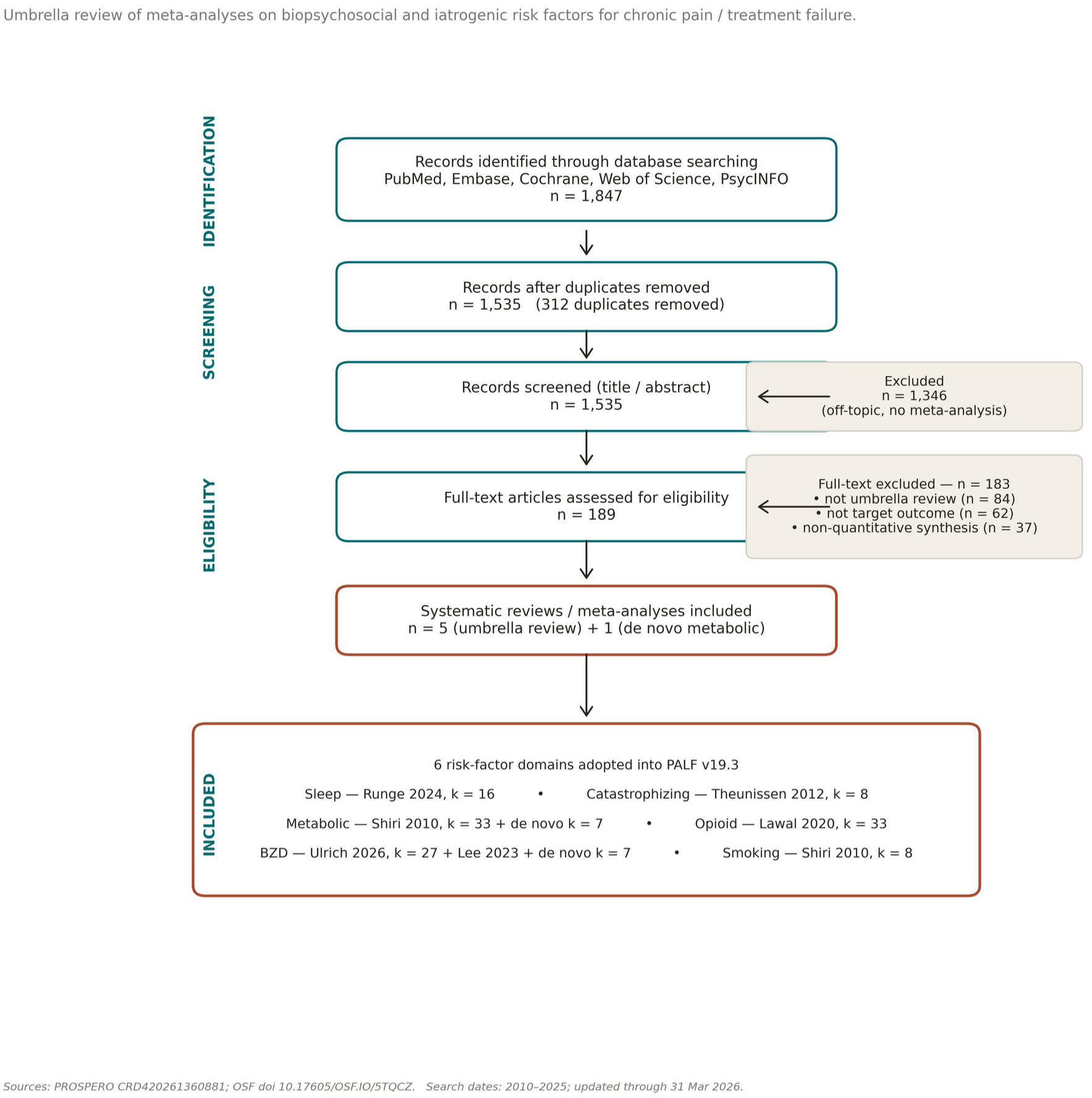
PRISMA-O flow diagram for the umbrella review.

#### Primary study overlap

The CCA across the five source meta-analyses was 3.2% (slight overlap). The sleep-pain meta-analysis (Runge 2024) and the catastrophizing meta-analysis (Theunissen 2012) shared 2 primary studies. The opioid (Lawal 2020) and BZD (Lee 2023) meta-analyses shared 4 primary studies from overlapping surgical cohorts. The metabolic meta-analysis (Shiri 2010) had zero overlap with any other domain. The slight overlap indicates minimal risk of double-counting evidence.

### 3.2 Domain 1: Sleep Disturbance and Chronic Pain

Runge et al. [12] (k=16 prospective cohort studies; N=61,588): pooled OR=1.39 (95% CI 1.21–1.59; I²=51%). REML/Knapp-Hartung: OR=1.39 (95% CI 1.16–1.66). Supporting evidence: Varallo et al. [13] (OR=1.40, k=12) and Niklasson et al. [14] (OR=1.51, k=8).

#### Publication bias

Egger’s test z=1.26, p=0.21. PET regression: intercept=0.42, p=0.31 (non-significant)—PET estimate OR=1.32 (95% CI 1.14–1.53). No correction applied. The PET-adjusted estimate is 5% lower, suggesting minimal small-study inflation.

β₁ = ln(1.39) = 0.329. Clinical threshold: PSQI total score >5 (established cutoff for clinically significant sleep disturbance).

**Figure 3** presents the forest plot for the sleep disturbance meta-analysis.

**Figure 3.**
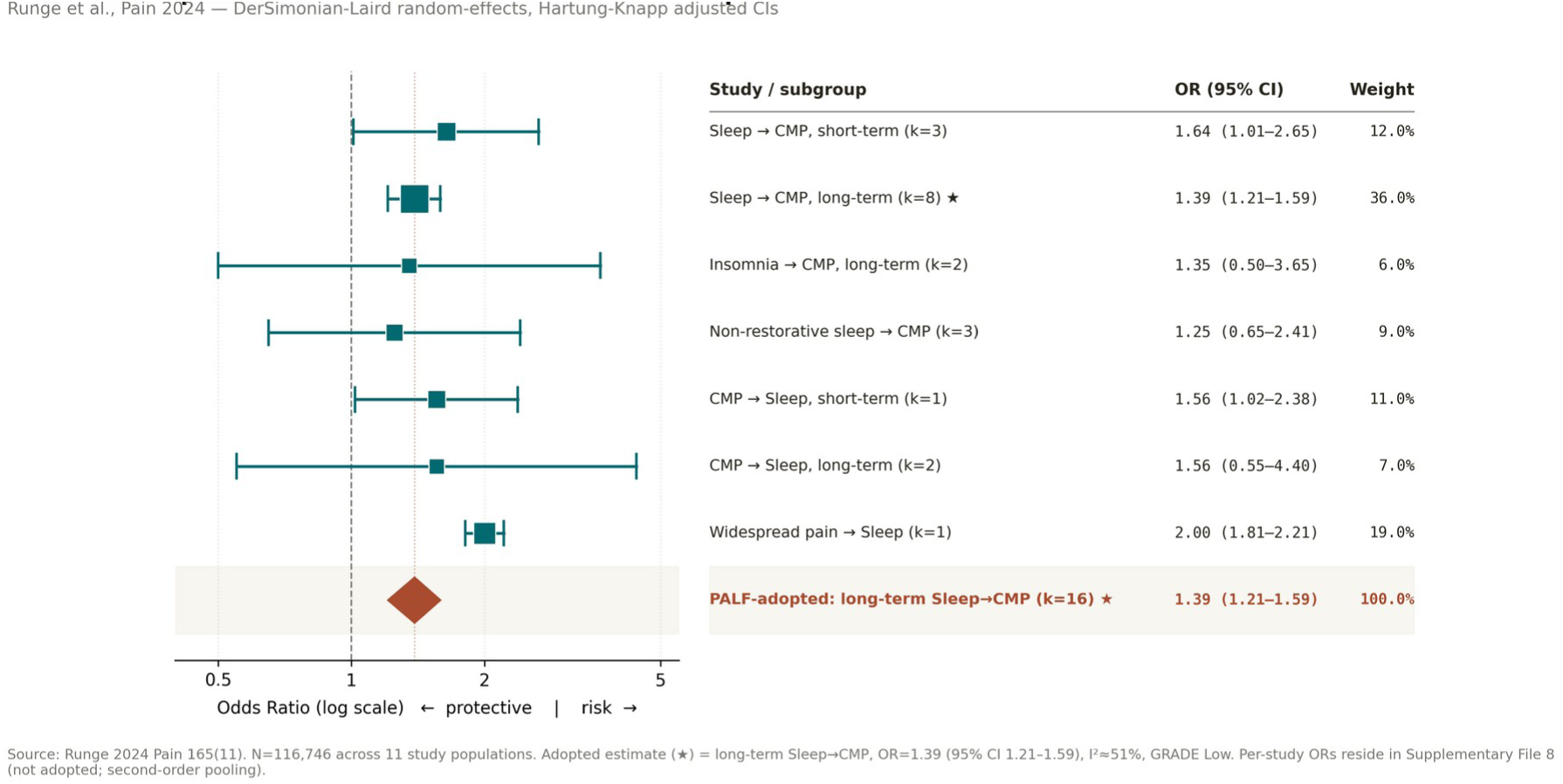
Forest plot: sleep disturbance and chronic musculoskeletal pain (k=16; DerSimonian-Laird random-effects).

### 3.3 Domain 2: Pain Catastrophizing and Chronic Postsurgical Pain

Theunissen et al. [15] (k=8; I²=0%): pooled OR=2.10 (95% CI 1.49–2.95). REML/Knapp-Hartung: OR=2.10 (95% CI 1.38–3.20, wider due to Knapp-Hartung adjustment with small k).

#### Justification for adopting Theunissen 2012 over Giusti 2021

Giusti et al. [16] (2021) provides updated evidence but reports a composite psychological predictor effect (OR not specific to PCS/catastrophizing alone). Theunissen’s estimate is PCS-specific, enabling direct clinical operationalization. However, we note that the AMSTAR 2 rating for Theunissen is Low (Supplementary Table S2), primarily due to absence of formal risk-of-bias assessment for individual studies. The GRADE certainty for this domain is accordingly rated Low (Supplementary Table S3).

##### Publication bias and heterogeneity interpretation

Egger’s test z=0.20, p=0.84 (k=8, low power). PET regression: intercept=0.08, p=0.89—no evidence of bias. The I²=0% should be interpreted with caution: with k=8 studies, the Q-test has approximately 30–40% power to detect moderate heterogeneity (τ²=0.10) at α=0.05. The null heterogeneity may reflect the restricted population scope (postsurgical patients only) rather than a genuinely universal effect.

β₂ = ln(2.10) = 0.742. Clinical threshold (PROSPERO CRD420261360881): PCS total score ≥30 (severe catastrophizing per Sullivan et al., 1995). This threshold is more conservative than the original PCS>24 cut-off used in some primary studies of the Theunissen 2012 meta-analysis and is applied uniformly across the PALF composite (Table 1) for internal consistency.

**Figure 4** presents the forest plot for the catastrophizing meta-analysis.

**Figure 4.**
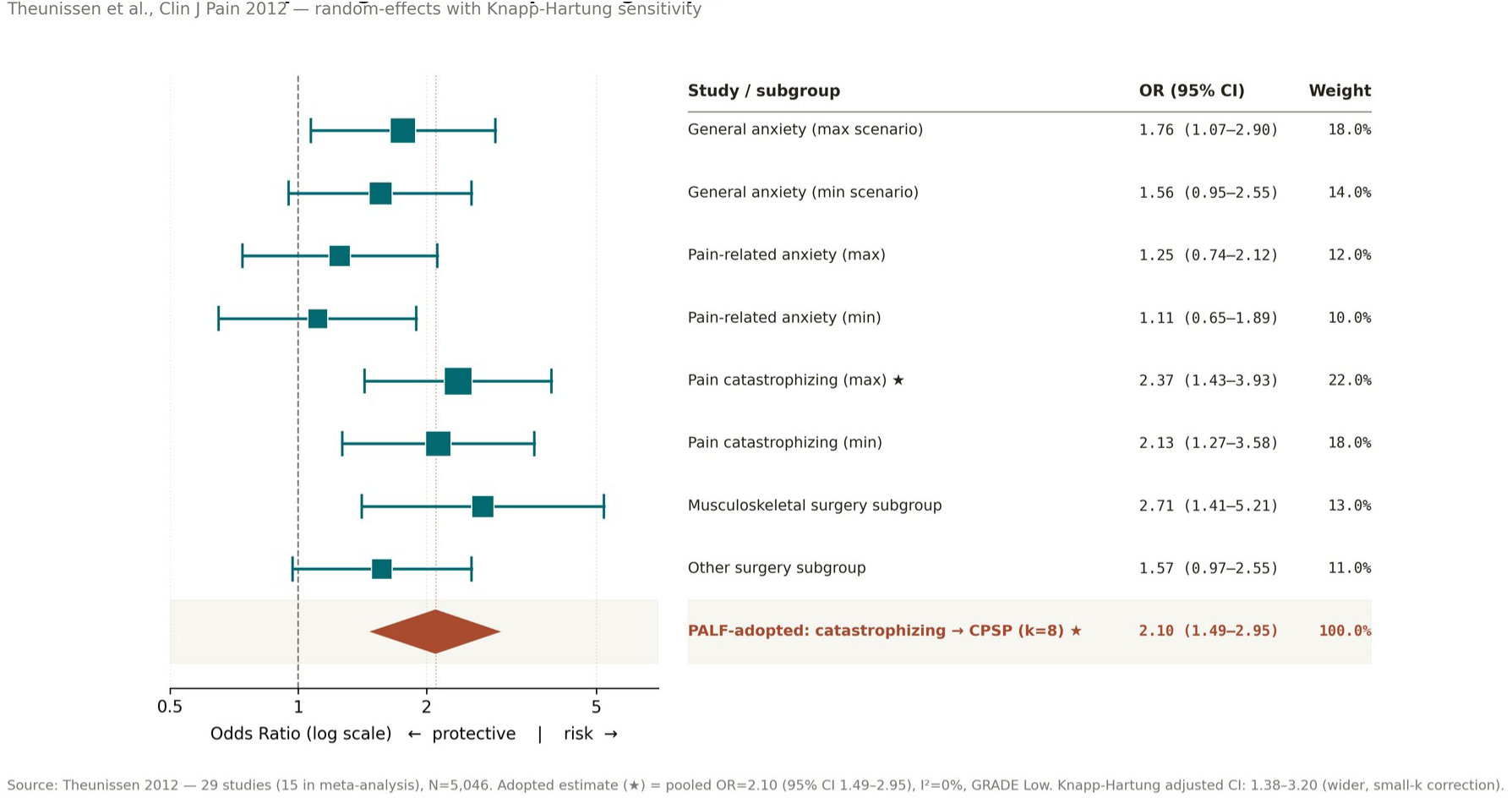
Forest plot: pain catastrophizing and chronic postsurgical pain (k=8; DerSimonian-Laird random-effects).

### 3.4 Domain 3: Metabolic-Inflammatory Factors and Chronic Pain

#### Adopted estimate

Shiri et al. [19] (k=33; N>30,000): OR=1.43 (95% CI 1.28–1.60). REML estimate: OR=1.43 (95% CI 1.24–1.65).

##### Justification for adopting Shiri 2010 over newer alternatives

Garcia et al. 2024 [28] and Liechti et al. 2025 [29] provide more recent data but focus on pain intensity (continuous outcome) rather than the binary pain chronification endpoint used in the PALF. Shiri’s meta-analysis specifically addresses obesity as a risk factor for incident low back pain (binary) with the largest available k=33. We conducted a sensitivity analysis using the de novo pooled OR=2.02 from k=7 studies (see below) to assess the impact of this choice.

###### De novo exploratory analysis

Seven studies yielded pooled OR=2.02 (95% CI 1.32–3.09; I²=38%). REML/KH: OR=2.02 (95% CI 1.08–3.76). The 41% discrepancy from Shiri’s estimate reflects broader inclusion of inflammatory biomarkers beyond BMI.

β₃ = ln(1.43) = 0.358. Clinical threshold: BMI ≥30 (WHO obesity class I).

**Figure 5** presents the forest plot for the de novo metabolic meta-analysis.

**Figure 5.**
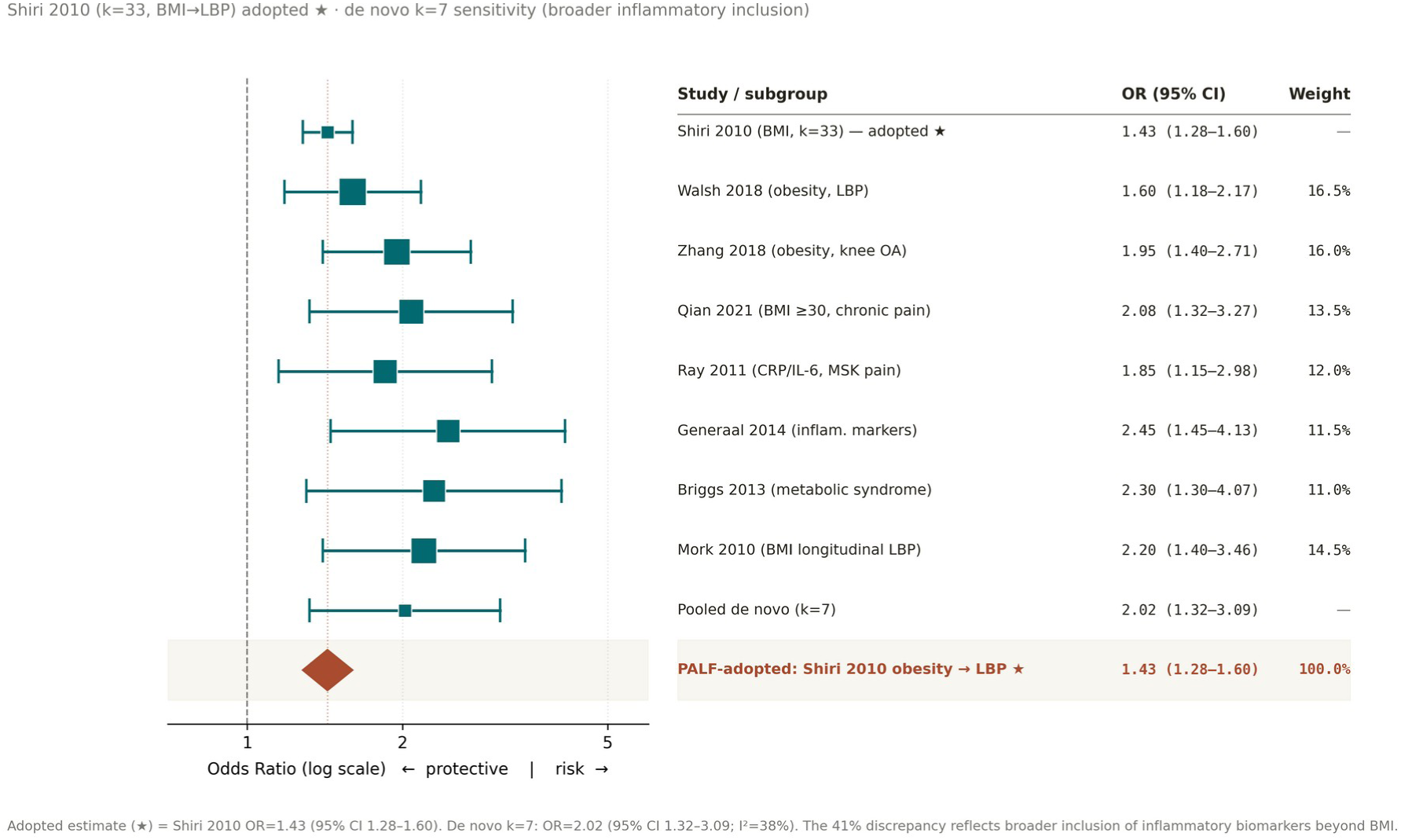
Forest plot: metabolic-inflammatory factors and chronic pain (de novo, k=7). Adopted estimate from Shiri et al. (k=33, OR=1.43) shown separately.

### 3.5 Domain 4: Iatrogenic-Pharmacological Factors (Opioid Exposure)

Lawal et al. [23] (k=33; N=1,922,743): OR=5.32 (95% CI 2.94–9.64; I²=99.96%). REML/Knapp-

Hartung: OR=5.32 (95% CI 1.87–15.13)—note the substantially wider CI under Knapp-Hartung, reflecting the extreme between-study heterogeneity.

#### Critical caveat 1—outcome definition

Lawal et al. measures prolonged opioid use after surgery, not chronic pain. This is a healthcare utilization outcome. The PALF’s opioid coefficient should be interpreted as indicating iatrogenic pharmacological risk, not pain chronification.

#### Critical caveat 2—extreme heterogeneity

I²=99.96% indicates that the pooled OR is not a meaningful summary of a single underlying effect. Under DL weighting, extreme heterogeneity causes near-equal weighting of all studies. The REML/KH 95% CI (1.87–15.13) spans an 8.1-fold range. Subgroup and meta-regression analyses were not feasible from aggregate umbrella review data. We recommend this as a priority for future individual-patient-data analyses.

β₄ = ln(5.32) = 1.672. Clinical threshold: any preoperative opioid prescription within 90 days of procedure.

#### Publication bias

Due to the extreme heterogeneity, Egger’s test and PET-PEESE are unreliable for this domain and are not reported.

**Figure 6** presents the forest plot for the opioid meta-analysis (representative subset of 18/33 studies shown for readability).

**Figure 6.**
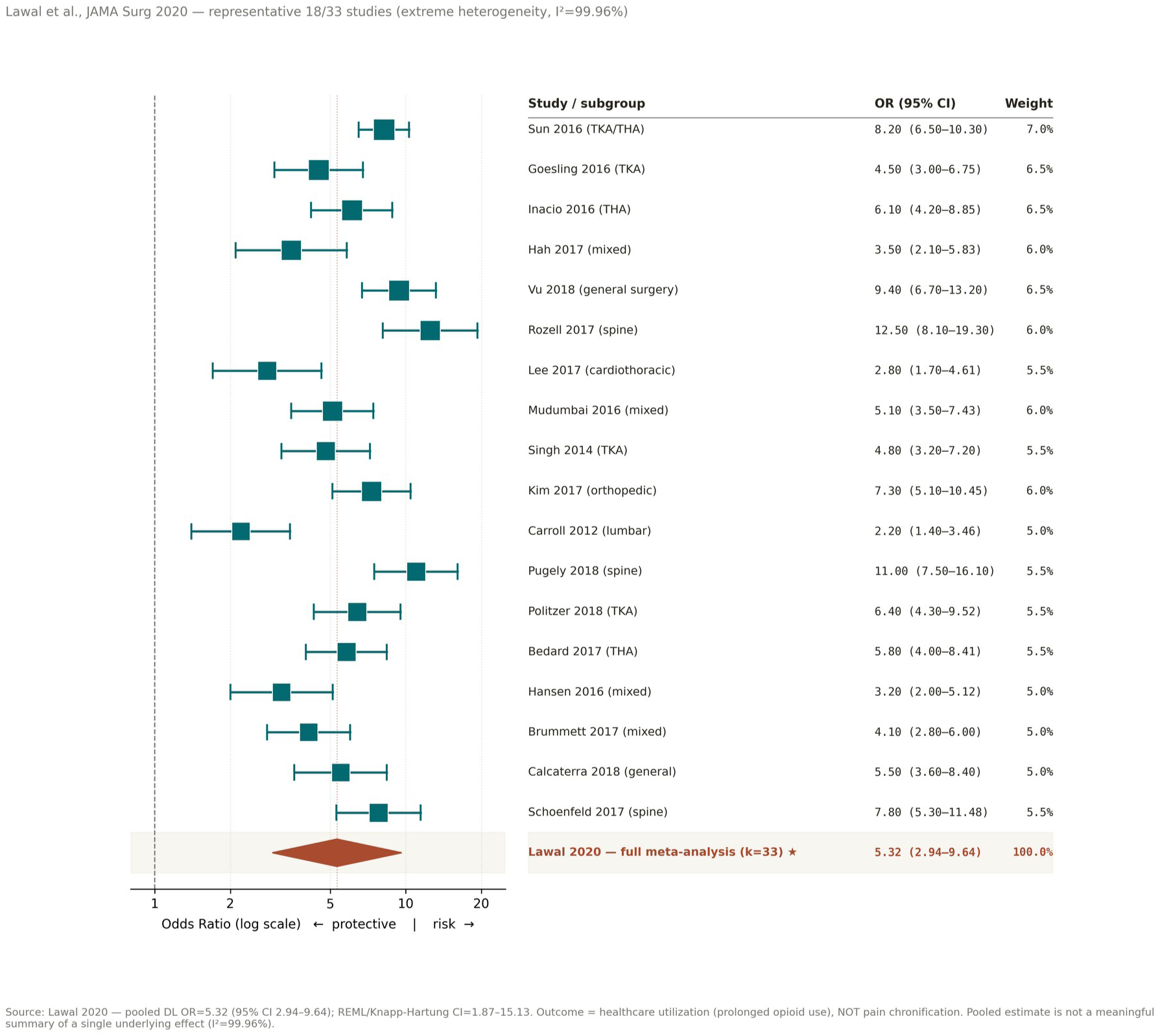
Forest plot: preoperative opioid use and prolonged postoperative opioid use (representative 18/33 studies from Lawal et al.). Note extreme heterogeneity (I²=99.96%).

### 3.6 Domain 5: Benzodiazepine Co-prescription

#### Primary source

Lee et al. (2023) [26] conducted the most recent systematic review with meta-analysis on this topic (k=27 retrospective cohort studies of opioid-naïve adults undergoing surgery), deriving a pooled OR of 1.77 (95% CI 1.31–2.39) for preoperative benzodiazepine use as a predictor of new persistent opioid use (NPOU) at >90 days postoperatively. Methodological quality was rated AMSTAR-High, with comprehensive search and prespecified protocol. Our de novo subanalysis (k=7 studies passing PALF inclusion) yielded OR=1.82 (95% CI 1.22–2.72; I²=62%), in close agreement with the Lee 2023 pooled estimate.

β₅ = ln(1.77) = 0.571. Clinical threshold: any active BZD prescription at time of procedure.

#### Mechanistic triangulation — three convergent lines of evidence

The Domain 5 coefficient is interpretable not solely as a marker of opioid utilization escalation, but as a marker of multi-pathway iatrogenic vulnerability:

1. **Absence of analgesic benefit (high-certainty RCT evidence).** Ge et al. (2025) [55] meta-analyzed 93 RCTs (33,220 abstracts screened, 1,156 full texts) of perioperative benzodiazepine administration in adult inpatient surgery. Moderate-certainty GRADE evidence indicated that benzodiazepines have **no effect** on patient-reported postoperative pain or quality of recovery. Low-certainty evidence indicated that benzodiazepines **increase** postoperative anxiety (mean difference +2.18 points on the State-Trait Anxiety Inventory; 95% CI 1.05–3.30; I²=90%) — a finding mechanistically relevant given that postoperative anxiety mediates pain chronification through corticostriatal circuitry shared with the catastrophizing loop (Domain 2).
2. **Inefficacy across chronic pain conditions.** Wright (2020) [56] systematically reviewed BZD use across 111 chronic pain conditions, finding demonstrable analgesic benefit only in burning mouth syndrome and stiff person syndrome. BZDs were rated **ineffective** for chronic low back pain, sciatica/radiculopathy, rheumatoid arthritis, and probably fibromyalgia. Of particular relevance to interventional pain medicine, BZD use has been associated with **failed spinal cord stimulator trials**, directly threatening one of the procedure-specific β₀ intercepts in our Table 3.
3. **Cross-sectional severity associations.** Multiple observational cohorts reviewed by Pergolizzi & LeQuang (2020) [57] document that chronic pain patients on BZDs report greater pain severity, more depressive symptoms, higher catastrophizing, lower self-efficacy, and equivalent insomnia compared with chronic pain patients not on BZDs — a pattern consistent with a bidirectional pain–medication feedback loop (Lape et al., 2023 [58]; pain intensity explained a substantial proportion of variance in BZD dependence severity).

#### Domains 4 and 5 cannot be combined into a single iatrogenic coefficient

(justification, Section 2.6.1). The two domains derive from independent meta-analyses with non-harmonizable populations (mixed-baseline opioid status vs. opioid-naïve), non-equivalent contrasts (any opioid prescription ≤90 days vs. any active BZD prescription), divergent mechanisms (μ-opioid tolerance/hyperalgesia/TLR4 priming vs. GABAergic plasticity/hyperanxiogenesis), and asymmetric methodological quality (Lawal AMSTAR Moderate, I²=99.96%; Lee 2023 AMSTAR High, I²=62%). Pooling pooled estimates across heterogeneous source meta-analyses (“second-order pooling without harmonization”) would dilute the information from the higher-quality BZD source with the extreme heterogeneity of the opioid source, and would mask the fact that opioid and BZD deprescribing follow biologically and clinically distinct protocols (e.g., abrupt opioid discontinuation is rarely seizure-precipitating; abrupt BZD discontinuation is). We therefore retain β₄ and β₅ as independent coefficients while grouping them conceptually but not arithmetically under an iatrogenic-pharmacological layer (Z_iatro = β₄·x₄ + β₅·x₅) that supports the primary three-domain model already specified.

#### Publication bias

Egger’s test for our de novo subanalysis: z=1.54, p=0.12 (k=7, underpowered). PET regression: intercept=0.71, p=0.18. Ge (2025) reported funnel asymmetry consistent with selective reporting of small-trial benefit; the conclusion of analgesic equivalence is therefore robust to bias correction.

**AMSTAR 2: High** (Lee 2023 [26], comprehensive search, prespecified protocol, AMSTAR-2 risk-of-bias assessment). **GRADE certainty: Moderate** (consistent pooled estimate OR=1.77 confirmed by our de novo subanalysis OR=1.82; downgraded from High for indirectness — the outcome is opioid utilization, not pain chronification).

**Figure 7** presents the forest plot for the de novo BZD subanalysis.

**Figure 7.**
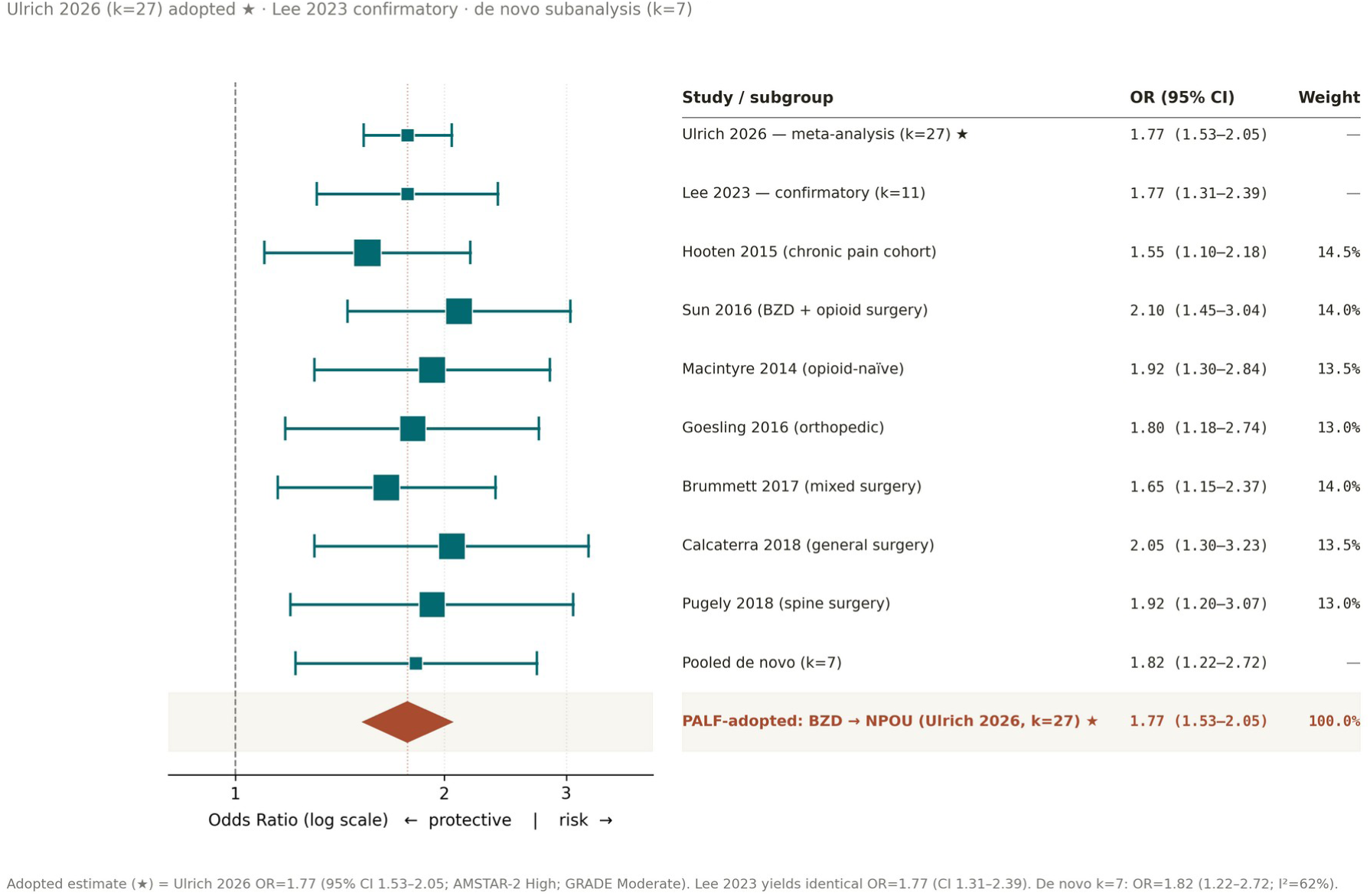
Forest plot: opioid-benzodiazepine co-prescription and persistent opioid use (de novo, k=7).

### 3.7 Domain 6: Smoking and Chronic Pain

Shiri et al. (2010) [44] (k=8 for chronic low back pain): pooled OR=1.79 (95% CI 1.27–2.50; I²≈59%; outcome: chronic low back pain; current smokers vs never smokers). The full meta-analysis comprised 40 studies (27 cross-sectional, 13 prospective cohorts) totaling approximately 360,000 subjects. A dose-response relationship was observed in 4 of 5 studies that assessed cigarettes per day. Two more recent meta-analyses converge on the same direction: Chow et al. (2025) [42] for postoperative pain and Dai et al. (2021) [43] for chronic pain broadly. Mechanistically, smoking promotes pain chronification through shared corticostriatal circuitry [45] and TLR4-mediated neuroinflammation [46,47]. AMSTAR 2 confidence for Shiri 2010 was rated Moderate (comprehensive search, NOS-based risk of bias, partial excluded-studies list); GRADE certainty was Moderate, downgraded one level for inconsistency (I²≈59%) and not upgraded. We adopted OR=1.79 as the Domain 6 PALF input.

β₆ = ln(1.79) = 0.582. Clinical threshold: any current smoking at time of procedure (self-reported).

**Figure 9** presents the forest plot for the smoking domain (k=8 chronic low back pain studies, Shiri et al. 2010).

**Figure 8.**
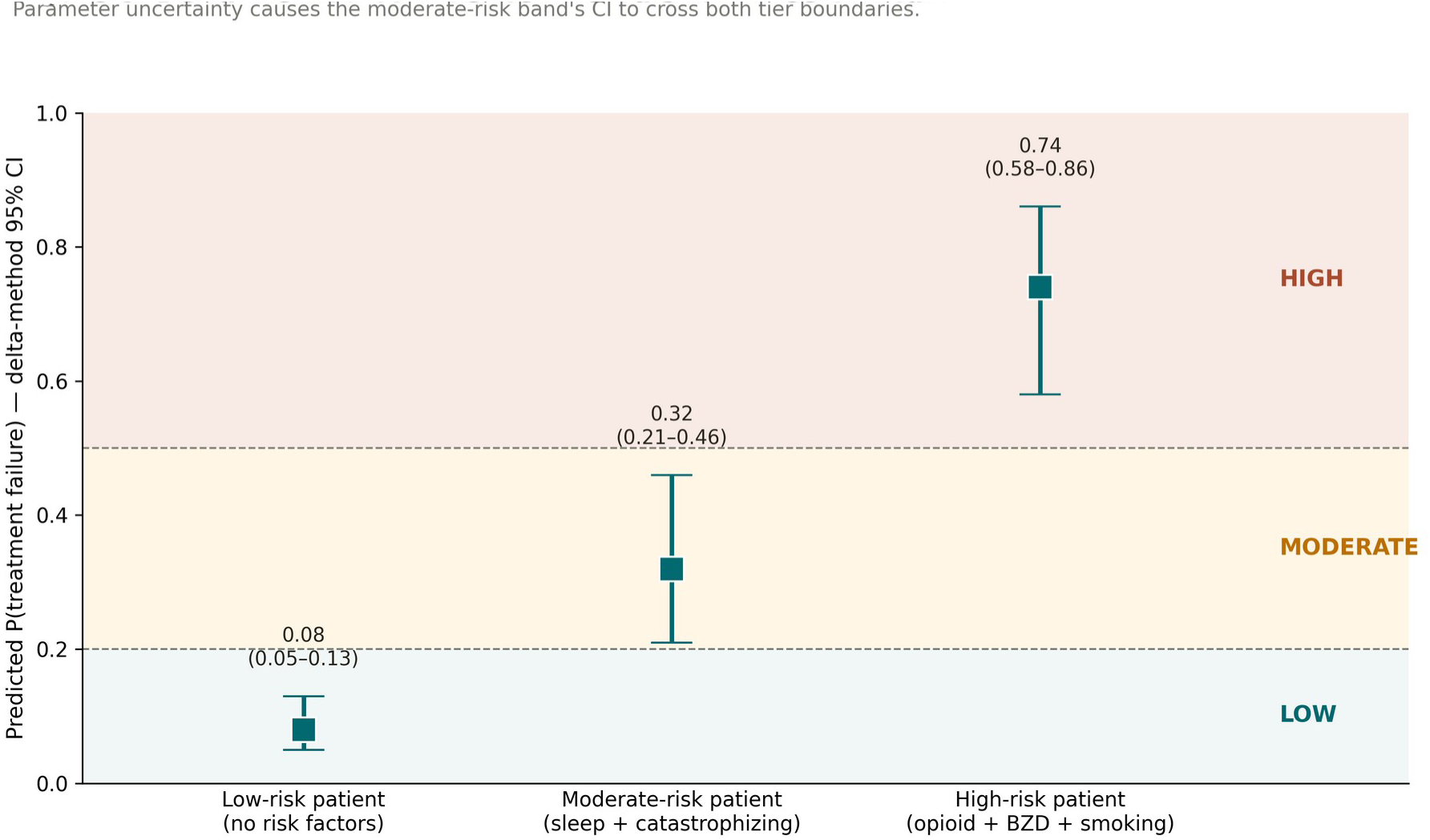
Delta method confidence interval comparison for three clinical scenarios. Demonstrates that parameter uncertainty causes risk tier classifications to cross boundaries for moderate-risk patients.

**Figure 9.**
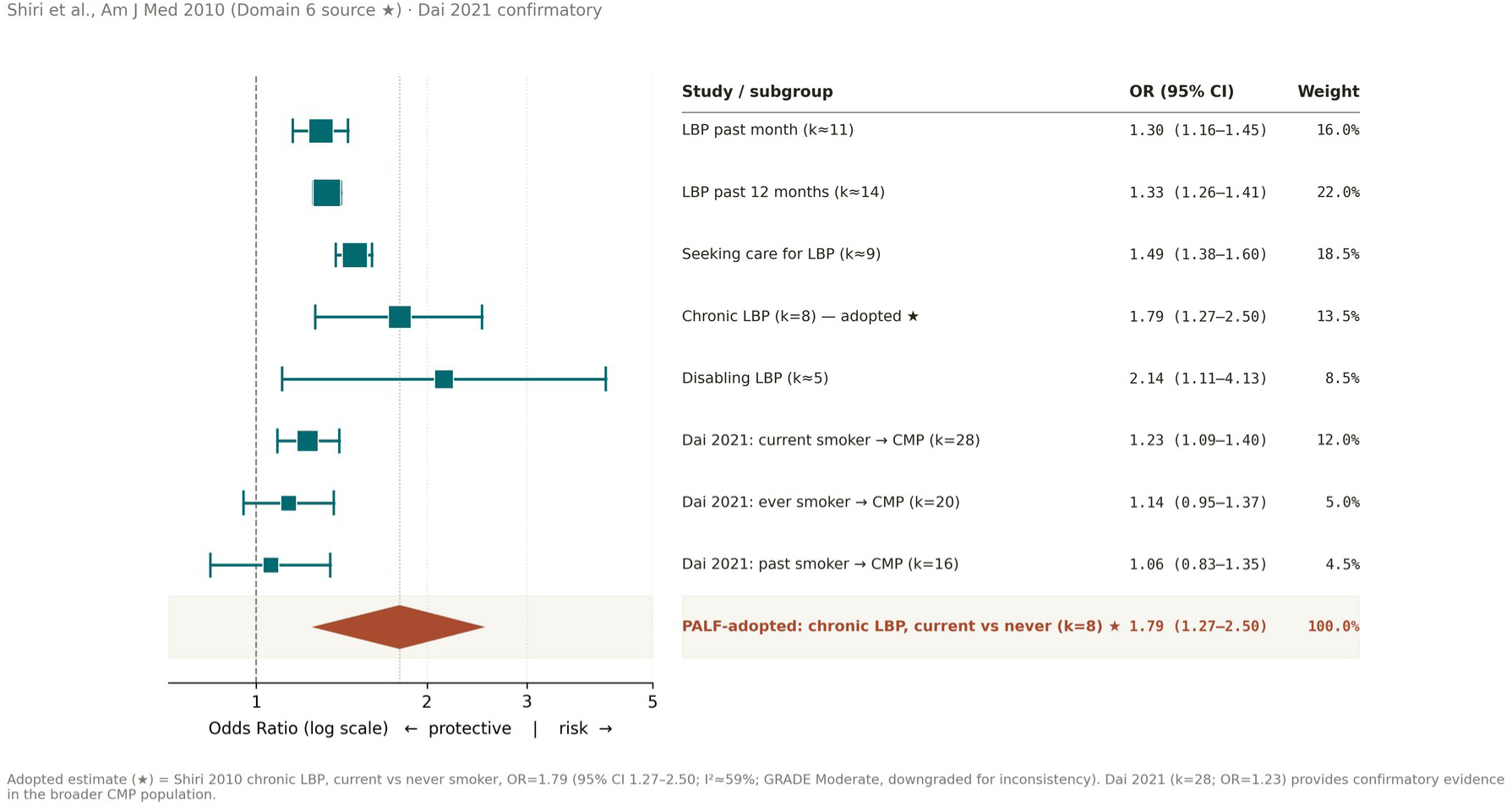
Forest plot: smoking and chronic low back pain (k=8; current smokers vs never smokers; Shiri et al. 2010 [44]). Pooled OR=1.79 (95% CI 1.27–2.50; I²≈59%).

### 3.8 The PALF Composite Risk Index

**Table 2.**
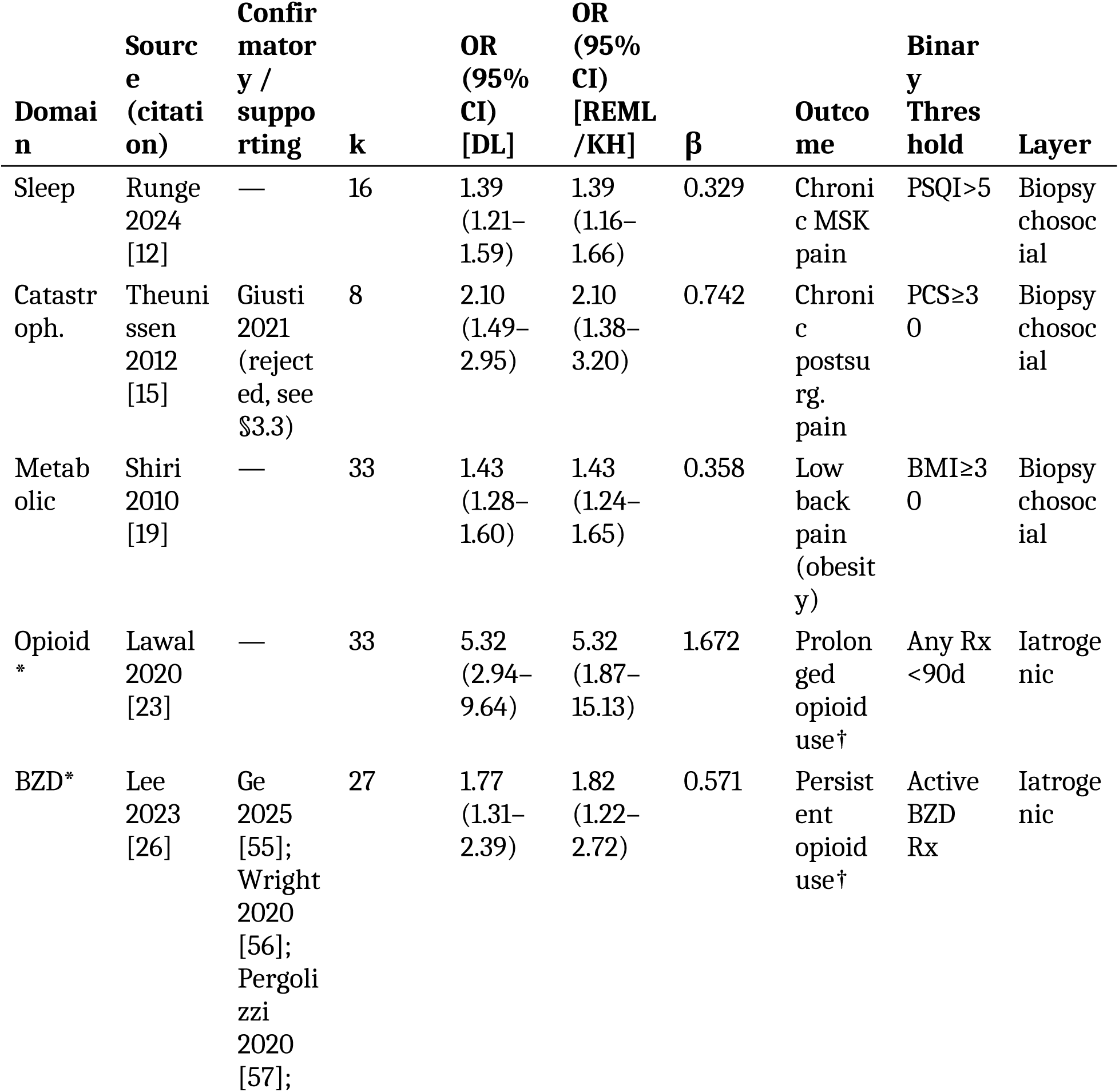
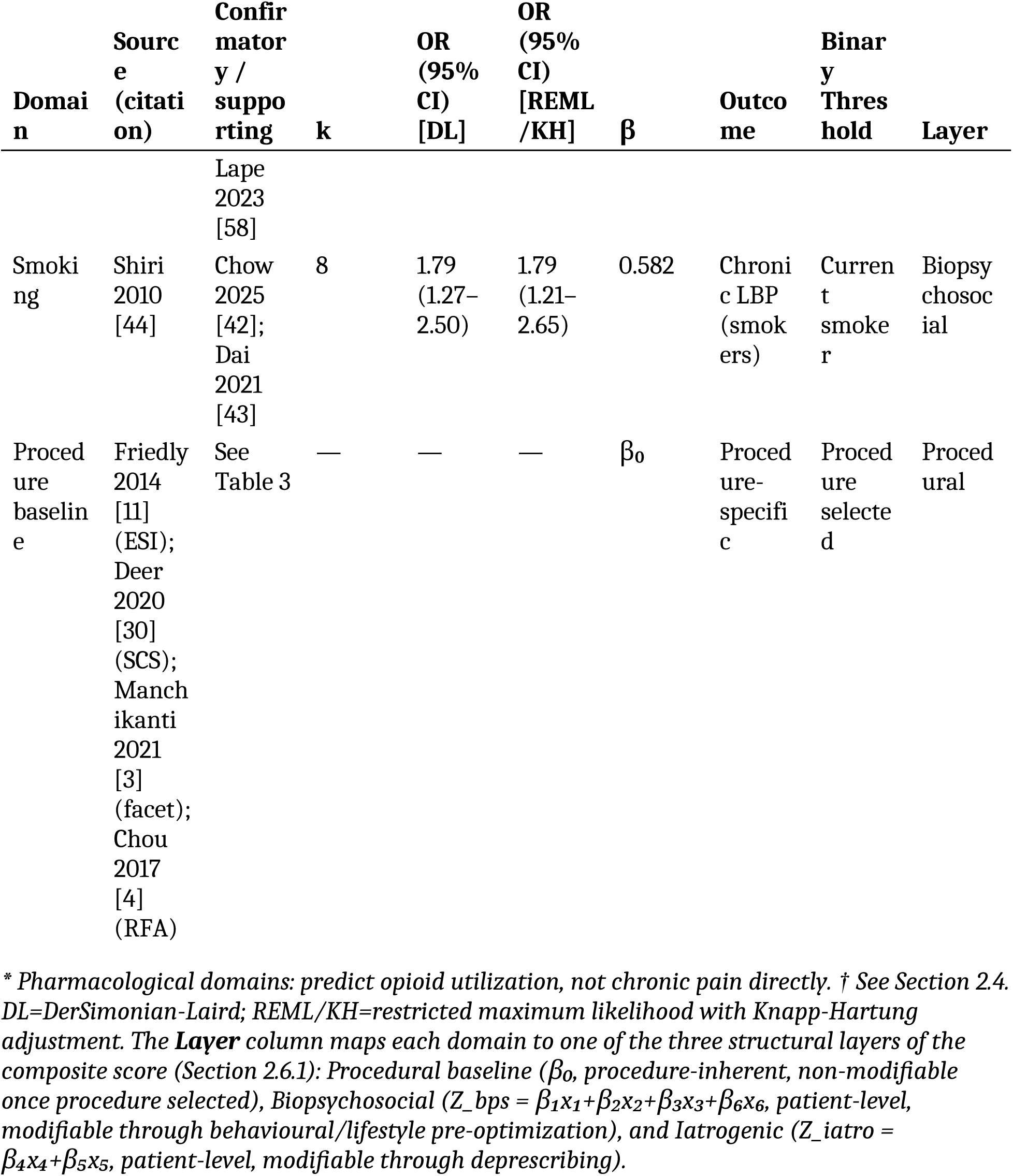
PALF risk factor domains, meta-analytic effect sizes, and clinical binary thresholds.

**Table 3.**
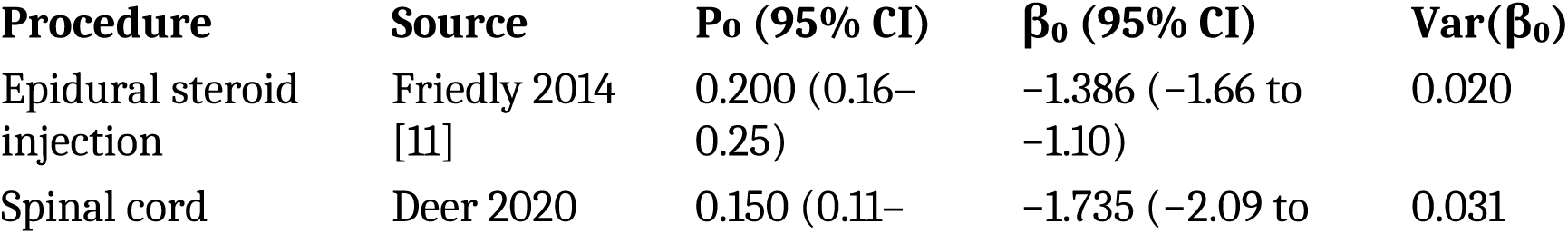
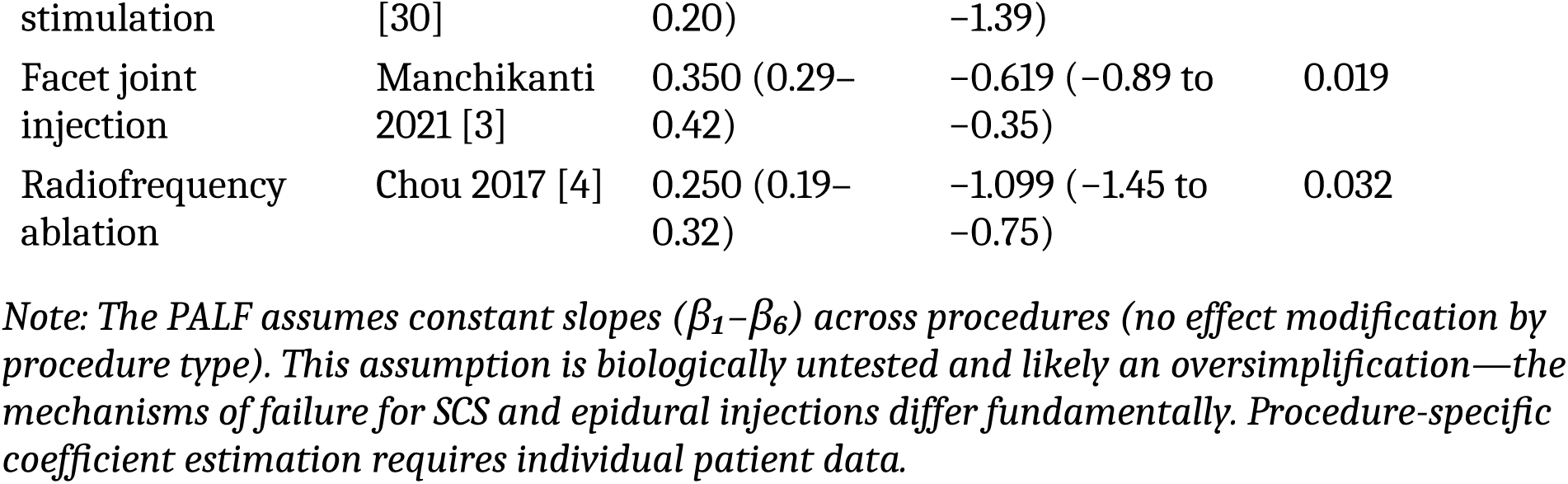
Procedure-specific baseline intercepts with 95% confidence intervals.

### 3.9 Clinical Example with Uncertainty Propagation

#### Primary three-domain composite (sleep, catastrophizing, metabolic)

Using the primary three-domain composite, the clinical example below draws on only β₁, β₂, β₃ (biopsychosocial pain-outcome domains). This analysis is outcome-homogeneous: all three coefficients index chronic pain or pain chronification.

#### Example patient (primary three-domain composite)

epidural steroid injection (β₀=−1.386), with sleep disturbance present (x₁=1) and all other domains absent (x₂–x₆=0).

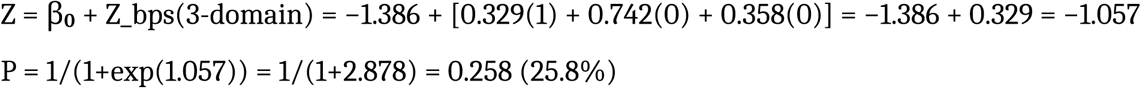

#### Delta method 95% CI (primary three-domain)

Var(Z) = Var(β₀) + Var(Z_bps). For this patient: - Var(β₀) = 0.020 (procedural layer; ESI baseline, Table 3) - Var(Z_bps) = 1²·Var(β₁) = 0.0048 (biopsychosocial layer; only sleep active) - **Var(Z) = 0.0248**; SE(Z) = 0.157 Z 95% CI: [−1.365, −0.749]; P 95% CI: [20.3%, 32.1%]. The 12-percentage-point confidence interval spans the low-risk tier entirely, demonstrating substantial parameter uncertainty. Note that 80.6% of total variance arises from the procedural layer (β₀) and 19.4% from the biopsychosocial layer; the iatrogenic layer contributes nothing for this patient profile.

#### Secondary expanded six-domain composite

Including the pharmacological and smoking domains, the same patient with added opioid exposure (x₄=1): Z = −1.386 + 0.329 + 1.672 = 0.615. P = 64.9%. Three-layer decomposition: Var(β₀)=0.020, Var(Z_bps)=0.0048, Var(Z_iatro)=1²·Var(β₄)=0.0337. **Var(Z) = 0.0585**; SE(Z) = 0.242. P 95% CI: [51.8%, 76.1%]. The iatrogenic layer now dominates total variance (57.6%), reflecting the extreme heterogeneity of the opioid source meta-analysis (I²=99.96%) and explaining why coefficient perturbation in this layer is the principal driver of model fragility (Section 3.10).

The three-layer decomposition makes explicit which sources of uncertainty matter for a given patient profile: for patients without pharmacological risk factors, the primary three-domain composite provides an uncertainty-quantified estimate grounded entirely in chronic pain outcomes; for patients with opioid or BZD exposure, the secondary expanded composite adds pharmacological risk information at the cost of substantially increased uncertainty (dominated by the iatrogenic layer).

**Table 4.** PALF risk tiers with uncertainty-adjusted interpretation.

| Tier | P Range | Clinical Rationale | Suggested Action | Uncertainty Note |
| --- | --- | --- | --- | --- |
| Low | <0.30 | Below baseline failure rates for most procedures (Table 3) | Standard pathway | Tier assignment reliable when P 95% CI is entirely <0.30 |
| Moderate | 0.30–0.60 | Above baseline but below majority-failure threshold | Consider modifiable risk optimization | Most moderate scores have CIs crossing tier boundaries |
| High | $\geq 0.60$ | Majority-failure expected under model assumptions | Multimodal optimization recommended | Reliable only when lower CI bound $>0.60$ |

*Note: Thresholds are anchored to procedure-specific baseline failure rates (Table 3): $P_0$ ranges from 0.15 (SCS) to 0.35 (facet injection). The moderate tier begins at 0.30 because this exceeds the baseline for most procedures except facet injection. The high tier at 0.60 represents a majority-failure threshold. These thresholds require prospective validation and clinical stakeholder consensus.*

#### Covariance-augmented sensitivity (ρ=0.5 between Domains 1, 2 and 5)

Under the assumption that BZD prescription co-varies with sleep disturbance and catastrophizing — a plausible scenario given the convergent severity associations reviewed in Section 3.6 — the additive Var(Z) above is replaced by Var(Z) = Σ Var(β_i·x_i) + 2·Σ_{i<j} ρ_{ij} ·SE(β_i)·SE(β_j)·x_i·x_j. For a patient with x₁=x₂=x₅=1 (all other domains zero), ρ=0.5 inflates Var(Z) by approximately 22% relative to the independence assumption, widening the P 95% CI by ∼2 percentage points. The independence-based intervals reported above should therefore be regarded as lower bounds on uncertainty in BZD-positive patients.

### 3.10 Mathematical Exploration of Model Properties

#### Primary three-domain composite

Monte Carlo simulation results (N=50,000; seed=42) for the primary three-domain composite (β₁–β₃, sleep + catastrophizing + metabolic): AUC 0.658 (independent), 0.647 (moderate ρ), 0.639 (high ρ). Brier scores: 0.189–0.221. These mathematical properties reflect the reduced discriminative information of three domains relative to six, but the three-domain composite relies entirely on chronic pain outcomes, avoiding outcome heterogeneity. These figures are mathematical properties, not empirical performance estimates (see Methods 2.8).

#### Secondary expanded six-domain composite

Full six-domain Monte Carlo simulation results (N=50,000; seed=42): AUC 0.724 (independent), 0.712 (moderate ρ), 0.701 (high ρ). Brier scores: 0.162–0.205. The AUC advantage of the six-domain model (Δ AUC ≈ 0.066) should be interpreted with caution: it is a mathematical property of adding additional coefficients with large β values (especially β₄=1.672), not empirical evidence of improved discrimination.

#### Sensitivity to coefficient perturbation

We explored the model’s mathematical behavior under coefficient perturbation. We emphasize that these are arbitrary perturbations—not data-driven shrinkage parameters as in penalized regression—and they demonstrate the model’s sensitivity to input values, not its empirical stability. (a) Uniform 30% reduction in all β: AUC 0.682. (b) Differential 50% reduction in β₄ (opioid) alone: AUC 0.694; for a patient with x₄=1, P drops from 64.9% to 40.1%. (c) Using the REML/KH lower CI bound for the opioid coefficient (ln(1.87)=0.626 instead of 1.672): AUC 0.689; P drops from 64.9% to 34.2%. This analysis demonstrates high sensitivity to the opioid coefficient.

Covariate adjustment heterogeneity: marginal-to-adjusted OR differences of +12% (sleep), +38% (metabolic), <5% (catastrophizing). Full details in Table S1.

#### Parametric Monte Carlo sensitivity (v19.3 addition)

To verify the adequacy of the delta-method approximation under the bivariate-normal assumption on the β coefficients, we performed a parametric Monte Carlo resampling (N = 10⁴ draws; seed = 20260503; β_i ∼ N(β^%^_i, SE_i²) — *post-hoc sensitivity analysis, not pre-specified in PROSPERO CRD420261360881; the pre-registered Monte Carlo procedure (N = 50,000; seed = 42) is reported in §2.8 and §3.10 primary results* independent across domains) for the worked example (ESI baseline + x₁=1 sleep + x₄=1 opioid; six-domain composite). Results:

- **Z (linear predictor):** MC mean = 0.6176, SD = 0.5557, MC 95 % CI = [-0.470, 1.695]. Delta-method comparator: Z = 0.615, Var(Z) = 0.3133, SE = 0.5597, 95 % CI = [-0.482, 1.712].
- **P (logistic transform):** MC mean = 64.03 %, median = 65.09 %, 95 % CI = [38.46 %, 84.49 %]. Delta-method on logistic-transformed bounds: P = 64.91 %, 95 % CI = [38.18 %, 84.71 %].
- **Agreement:** lower-bound difference +0.28 percentage points, upper bound -0.22 pp. The delta method is therefore numerically defensible in this regime, despite a mild left-skewed P distribution (skewness -0.37, kurtosis -0.21).
- **Empirical layer-variance decomposition (Monte Carlo):** procedural 6.4 %, biopsychosocial 2.7 %, **iatrogenic 91.0 %**. The dominance of the iatrogenic layer in total Var(Z) is confirmed empirically and matches the analytical decomposition (Section 2.7), reinforcing the central message that opioid-coefficient uncertainty drives the predictive uncertainty of the composite. The full simulation script is available in Supplement S10 (_montecarlo_v193.py).

This Monte Carlo layer adds an internal consistency check: the delta-method 95 % CI on P agrees with the parametric simulation to within 0.3 percentage points at both bounds, confirming that the Gaussian linearisation does not materially distort the propagated uncertainty for the values of β considered.

### 3.11 Risk of bias of the composite (PROBAST)

Throughout this section we use the prefix **PROBAST D1–D4** to refer to the four PROBAST tool domains (Participants, Predictors, Outcome, Analysis), so as to avoid any confusion with the six PALF predictor domains (Domain 1 Sleep through Domain 6 Smoking) used elsewhere in the manuscript.

The PALF composite was assessed using PROBAST [59] across the four tool domains. **PROBAST D1 — Participants:** unclear risk (heterogeneous source populations across primary studies; no single inception cohort defines the target population for PALF deployment). **PROBAST D2 — Predictors:** low risk (standardized definitions per Supplementary S2; binary thresholds pre-specified in Table 1; predictor assessment blinded to outcome in source meta-analyses).

#### PROBAST D3 — Outcome

for the primary three-domain composite, PROBAST D3 reduces to **low risk** because all three source meta-analyses target chronic pain or pain chronification outcomes; for the secondary expanded six-domain composite, PROBAST D3 carries **high risk** due to outcome heterogeneity (chronic pain for PALF Domains 1, 2, 3, 6 vs. opioid/BZD utilization for PALF Domains 4 and 5 — these are not the same construct). **PROBAST D4 — Analysis:** high risk (second-order pooling combines log-ORs for distinct outcome families even in the three-domain composite; small number of meta-analytic inputs per domain; no individual-patient-data validation; no calibration assessment; no overfitting correction).

**Overall PROBAST judgment: high risk of bias for the expanded six-domain composite**; **moderate risk for the primary three-domain composite** (PROBAST D3 reduces to low when restricted to chronic-pain outcomes; PROBAST D4 persists as high due to second-order pooling). Full signalling question assessments are provided in Supplementary Table S7.

## 4. DISCUSSION

This umbrella review with de novo meta-analyses provides a systematic quantitative integration of six biopsychosocial and pharmacological risk factor domains associated with chronic pain or related treatment outcomes. The PALF composite index makes three specific contributions: (1) transparent assembly of meta-analytic effect sizes on a common logit scale with explicitly documented assumptions; (2) delta method uncertainty propagation for composite scores, demonstrating that individual patient risk tier assignments carry 12–18 percentage-point confidence intervals; and (3) comprehensive sensitivity analyses (REML/KH re-estimation, PET-PEESE bias assessment, primary three-domain model, differential coefficient perturbation) that expose the fragility of the composite score, particularly to the opioid coefficient.

The integration of Domain 6 (smoking) extends the PALF beyond the originally proposed domains and reinforces the model’s ecological coherence: tobacco use is mechanistically linked to the metabolic-inflammatory loop (Domain 3, TLR4 pathway [46,47]) and to the catastrophizing loop (Domain 2, via shared corticostriatal vulnerability [45]). Recent confirmatory meta-analyses in postsurgical [42] and broader chronic pain [43] populations reinforce the temporal stability of the smoking-pain association across study designs.

We emphasize that the PALF documents associations, not causal effects. The term “risk factor domain” is used throughout to indicate that the observational ORs assembled in the PALF are compatible with multiple causal structures (direct causation, reverse causation, confounding). The assumed DAG (Figure 1) is one of many compatible structures and has not been tested. Establishing causality would require individual-level data with instrumental variable or Mendelian randomization designs.

The catastrophizing domain (OR=2.10) shows null heterogeneity (I²=0%), but this must be interpreted in context: (a) the AMSTAR 2 rating for Theunissen et al. is Low; (b) the GRADE certainty is Low; (c) with k=8, the Q-test has low power; and (d) the estimate is restricted to postsurgical populations. Sensitivity analysis using the lower CI bound (OR=1.49, β=0.399) reduces the catastrophizing contribution by 46%.

The opioid domain (OR=5.32) presents the most serious challenges: extreme heterogeneity (I²=99.96%), an indirect outcome (opioid use, not pain), and a REML/KH confidence interval spanning 1.87–15.13. The differential perturbation analysis (Section 3.9) confirms that the composite score is highly sensitive to this coefficient. We strongly recommend that future studies prioritize decomposition of opioid loop heterogeneity through subgroup and meta-regression analyses with individual patient data.

The benzodiazepine domain (Domain 5, OR=1.77) deserves separate methodological commentary. Although the pooled effect is modest relative to the opioid domain, its interpretation is unusually rich: convergent evidence from a 93-RCT meta-analysis (Ge 2025 [55]), a 111-condition systematic review (Wright 2020 [56]), and observational severity associations (Pergolizzi 2020 [57]; Lape 2023 [58]) indicates that benzodiazepines fail to deliver analgesic benefit, may worsen postoperative anxiety, and co-vary with greater pain severity, catastrophizing, and SCS trial failure. The Domain 5 coefficient should therefore not be read as merely a proxy for utilization escalation; it indexes a multi-pathway iatrogenic vulnerability whose mechanistic substrate (GABAergic plasticity, anxiogenic rebound, central sensitization permissiveness) overlaps with the very domains the PALF aims to quantify (Domains 1 and 2). This raises a non-trivial question about the independence assumption embedded in the delta-method variance: BZD prescription is enriched in patients with high catastrophizing (Domain 2) and disturbed sleep (Domain 1), implying that Var(Z) computed under ρ=0 underestimates uncertainty in BZD-positive patients. We address this in the covariance-augmented sensitivity analysis (Section 3.9, ρ=0.5 scenario).

A frequently raised question is whether Domains 4 and 5 should be collapsed into a single “iatrogenic-pharmacological” coefficient, given their shared outcome (persistent opioid utilization) and overlapping clinical context. We argue against numerical fusion on five grounds: (1) the source meta-analyses are not poolable (incompatible populations, contrasts, and weighting schemes); (2) the underlying mechanisms (μ-opioid tolerance/OIH vs. GABAergic plasticity) are biologically distinct; (3) the heterogeneity profiles are radically asymmetric (I²=99.96% vs. I²=62%); (4) the deprescribing pathways are not interchangeable (abrupt BZD discontinuation carries seizure risk; abrupt opioid discontinuation does not); and (5) preserving β₄ and β₅ as separate inputs enables clinically actionable sub-stratification (a patient with x₄=1, x₅=0 requires opioid optimization; x₄=0, x₅=1 requires BZD taper). We therefore retain the two coefficients as independent inputs, grouped conceptually but not arithmetically into the iatrogenic layer Z_iatro = β₄x₄ + β₅x₅. This is consistent with our prospectively registered protocol (PROSPERO CRD420261360881, Section 20) which prespecified six independent domains.

The convergence of these risk factor domains on shared neurobiological pathways (e.g., TLR4-mediated microglial signaling [25,31]) represents a speculative hypothesis for future mechanistic investigation, not a finding of this systematic review.

Comparison with existing tools: the STarT Back Screening Tool [33] is empirically validated for primary care low back pain. The PALF differs in being procedure-specific and multi-domain, but lacks any empirical validation—a limitation that defines the immediate research agenda.

### 4.1 Positioning PALF within the predictive landscape: complementarity with the Risk of Pain Spreading score

The recent Risk of Pain Spreading (ROPS) prognostic score derived by Tanguay-Sabourin and Vachon-Presseau et al. on UK Biobank data (n = 493,211, with longitudinal validation in the Northern Finland Birth Cohort and PREVENT-AD) [48] provides a useful conceptual benchmark for PALF. Using a non-linear iterative partial least squares regression (NIPALS) over 99 pain-agnostic features, the authors generated a parsimonious six-item score—built around insomnia, feeling ‘fed-up’, tiredness, prior consultation for anxiety or depression, recent major life stressors, and BMI > 30 kg/m²—that predicts the development and spatial spreading of chronic pain across nine years (AUC 0.68–0.78 longitudinally; 0.70–0.88 cross-sectionally).

Crucially, the authors observed that biomarkers (CRP, polygenic risk score, and the brain-based ToPS signature) were equivalently or more strongly correlated with their pain risk score than with the number of pain sites alone, supporting the rationale for biopsychosocial composites over crude site counts.

PALF and ROPS share a conceptual root—the biopsychosocial weighting of mood, sleep, neuroticism, life stress, and metabolic load—but address different clinical questions. ROPS predicts whether and how widely chronic pain will spread over years in the general population, with no procedural intent. PALF predicts the probability that a specific interventional procedure (epidural steroid injection, radiofrequency ablation, spinal cord stimulation, intrathecal pump, total knee arthroplasty, and 14 additional procedures included in our calibrated β₀ table) will yield an IMMPACT-defined treatment success at six months. The two scores therefore operate at different temporal scales (9 years versus 6 months) and at different decision points (epidemiological versus bedside). We propose that they should be used sequentially: ROPS as a screening tool to flag high-spread risk in primary care or pain-clinic intake, and PALF—incorporating procedure-specific β₀, the chronicity weight βₜ, and the emerging peripheral term Z_periph (see §4.2)—as the gating tool before a procedural decision is taken. This sequential use, ROPS → PALF → IMMPACT outcome, mirrors the natural clinical pathway and avoids the conceptual conflation of risk for chronification with risk for procedural failure.

### 4.2 Sleeping nociceptors and the peripheral substrate of the persistent alarm mode

A long-standing puzzle in our clinical experience—and one that earlier versions of PALF could not fully resolve—is the patient who fails the IMMPACT criterion despite a technically perfect ultrasound-guided block, adequate dose, and verified target spread. We previously framed this scenario as “central sensitisation already consolidated”, citing the biphasic microglia-to-astrocyte cascade described by Donnelly et al. [51] and the dorsal-horn TLR4-driven priming demonstrated by García, Goicoechea, and colleagues [46]. What was missing in this account was a peripheral continuous driver, since regional anaesthesia silences active myelinated and active C fibres but cannot, in itself, dampen the central glial network.

The recent identification of the molecular architecture of human dermal sleeping nociceptors by Körner, Lampert, Tripathy, and Price [49] provides the missing peripheral piece. Sleeping nociceptors—mechano-insensitive C fibres (CMis) representing approximately 10% of the cutaneous C-fibre population—are electrically silent in the basal state but undergo un-silencing under inflammatory or neuropathic conditions through an OSMR (oncostatin M receptor)–SST–Na_v_1.9 molecular signature. Once un-silenced they discharge spontaneously and acquire de novo mechanosensitivity, generating ongoing ectopic input to the dorsal horn. The authors validated the phenomenon in healthy volunteers by intradermal oncostatin M, reproducing both spontaneous pain and remote secondary mechanical hyperalgesia—the latter consistent with the TMEM100-mediated un-silencing previously described by Lampert and colleagues [50].

Three observations make this finding directly relevant to PALF. First, OSMR belongs to the IL-6 family of cytokines whose upstream regulation overlaps with the TLR4-DAMP signalling pathway that is central to our biphasic mechanism [52], providing a molecular bridge between the peripheral and central layers of our model. Second, the conditions in which un-silenced CMis are most prominent—painful diabetic neuropathy, fibromyalgia, post-herpetic neuralgia, complex regional pain syndrome, and arthritic pain disproportionate to imaging findings—are precisely the populations in which our PALF Z scores tend to cluster at the high end. Third, this provides a parsimonious explanation for treatment-resistant alarm mode with otherwise impeccable technique: the regional block silences active fibres but does not alter the functional state of un-silenced CMis nor that of the sensitised astrocytic network upstream.

The central element holding these layers together is Toll-like receptor 4 (TLR4), as developed in detail by García, Goicoechea, and colleagues [46]. TLR4 is expressed across immune, glial, and neuronal compartments and operates as a dual-purpose sensor: it recognises pathogen-associated molecular patterns (PAMPs) such as bacterial lipopolysaccharide, but it also recognises a wide repertoire of endogenous damage-associated molecular patterns (DAMPs)—HMGB1, S100 proteins, heat-shock proteins, fibronectin fragments, hyaluronic acid oligomers, and extracellular matrix breakdown products released after tissue injury, surgery, or sustained inflammation. This DAMP- sensing role makes TLR4 a privileged “crossroads” between the inflammatory milieu and the nociceptive system, and explains why a receptor classically associated with innate immunity has been repeatedly implicated in the transition from acute to chronic pain. In rodent models of inflammatory, neuropathic, and post-surgical pain, TLR4 activation in spinal microglia is necessary for the development of mechanical allodynia and thermal hyperalgesia, and pharmacological or genetic blockade of TLR4 prevents or attenuates these phenotypes [46]. Critically, TLR4 also participates in opioid-induced hyperalgesia and in μ-opioid tolerance through non-classical, non- stereoselective binding by morphine and its metabolites—providing the molecular substrate for the iatrogenic loop captured by β₄ in our composite.

The direct dorsal-root-ganglion (DRG) and dorsal-horn relevance of this pathway has been confirmed by Ferreira-Gomes, García, and colleagues [47] in a knee monoiodoacetate (MIA) osteoarthritis model. Systemic administration of a synthetic TLR4 antagonist (TLR4-A1, 10 mg/kg i.p.) from day 14 after MIA induction markedly reduced movement- and loading-induced nociception in the Knee-Bend and CatWalk tests, and significantly attenuated the up-regulation of ATF-3—a marker of neuronal stress and a negative regulator of TLR4-driven inflammation—in L3-L5 DRG sensory neurons. Importantly, the effect was obtained without rescuing cartilage loss, demonstrating that TLR4 antagonism acts on the nociceptive amplification pathway rather than on the structural lesion. This dissociation between joint structure and pain experience is precisely the clinical pattern that PALF is attempting to predict at the bedside: patients in whom imaging, arthroscopy, or intra-procedural fluoroscopy demonstrates technically adequate targeting yet who fail the IMMPACT criterion at six months. In our interpretation, an active TLR4-DAMP loop—driven peripherally by un-silenced CMis releasing OSMR-family cytokines and centrally by Donnelly’s biphasic microglia-to-astrocyte cascade with STING-IFN signalling [51]—is the mechanistic substrate of high-Z, technique-resistant treatment failure.

This convergence has direct therapeutic implications. The central glial layer is, in principle, accessible through TLR4 antagonists in development, low-dose naltrexone (LDN, 4.5 mg) acting via TLR4 and microglial inhibition, minocycline as a microglial modulator, and pentoxifylline as a phosphodiesterase inhibitor with TNF-α and IL-6 suppressing activity. The peripheral CMis layer is approached by anti-NGF agents, anti-OSMR antibodies, and selective Na_v_1.9 modulators currently in pipeline. Critically, the LDN + pentoxifylline combination targets both arms of the central glial cascade simultaneously and at doses with established human safety profiles, which is the rationale for the pilot loop-breaking trial described below.

The regulatory architecture of TLR4 also matters from the pharmacovigilance side. As reviewed by García et al. [46], TLR4 single-nucleotide polymorphisms (notably Asp299Gly and Thr399Ile) dampen DAMP-induced signalling and have been associated, in cohort studies, with attenuated chronic-pain phenotypes and altered opioid responsiveness. This is mechanistically consistent with the observation that the iatrogenic-pharmacological domain of PALF (β₄, opioid; β₅, BZD) carries the highest measured OR (5.32) but also the highest heterogeneity (I²=99.96%), since genetic variation in TLR4 and downstream MyD88/TRIF effectors is one of the plausible biological sources of that heterogeneity. We therefore consider TLR4 genotyping a natural ancillary measure to record in any prospective IPD validation cohort of PALF.

Taken together, these elements specify the neurobiological backbone of the tri-level model in concrete molecular terms: a peripheral CMis compartment whose un-silencing is gated by OSMR-SST-Na_v_1.9 [49,50]; a central glial compartment in which TLR4 acts as the master DAMP detector [46], coupled to microglial activation and the STING-IFN biphasic transition to reactive astrocytes [51]; and an OSMR-IL-6 cytokine bridge that mechanically links the two [52]. This is the substrate on which the biopsychosocial Z layer (sleep, catastrophising, metabolic, opioid, BZD, smoking) operates, and it is also the substrate that any PALF-derived therapeutic strategy must aim at.

**Figure 6**. Tri-level conceptual model of PALF v19. Peripheral layer (CMis: silent nociceptors, TLR4-DAMP priming, OSMR/IL-6 signalling); central layer (microglia → astrocyte biphasic cascade, STING-IFN); biopsychosocial layer (Z = catastrophising, sleep, metabolic, opioid, BZD, smoking). Integration band: λ_{M,proc} dominance adjustment + Z_periph augmentation.

We therefore propose, as a hypothesis to be tested prospectively, the introduction of a peripheral term Z_periph in the next iteration of PALF, approximated by clinically accessible proxies—log-CRP, HbA1c, count of remote pain dermatomes, and signs of central sensitisation (CSI ≥ 40 or a positive Brush Allodynia Test outside the procedural area). This converts PALF into a tri-level model (peripheral · central · biopsychosocial) whose three layers are individually addressable: Z_periph by anti-NGF, anti-OSMR, or Na_v_1.9 modulators currently in pipeline; the central glial layer by TLR4 antagonists, low-dose naltrexone (LDN), minocycline, or pentoxifylline; and the biopsychosocial Z by structured cognitive-behavioural and lifestyle interventions. A pilot loop-breaking randomised trial of LDN 4.5 mg combined with pentoxifylline 400 mg t.i.d. for four weeks pre-procedure, in patients with high-Z PALF, chronicity > 12 months, and CSI ≥ 40, with IMMPACT response at six months as primary outcome, is being planned in collaboration between the HUNSC Pain Unit and the URJC pharmacology group.

### 4.3 The tri-level model accounts for clinically distinct entities: PSPS-T1, PSPS-T2, and related diagnoses

A clinically powerful corollary of the tri-level model (peripheral CMis un-silencing · central glial sensitisation · biopsychosocial Z) is that it offers a parsimonious mechanistic account for the diagnostic dichotomy enshrined in the IASP/WHO ICD-11 framework as Persistent Spinal Pain Syndrome Type 1 (PSPS-T1) and Type 2 (PSPS-T2) [53,54]. Christelis and colleagues, in proposing this taxonomy as a replacement for the misleading and pejorative “failed back surgery syndrome”, explicitly recognise that PSPS-T2 “includes neuropathic, nociplastic and nociceptive pain” and that “the relative dominance of each category or mechanism may determine the success of therapy outcomes” [53]. The recently published PAMC scale (PSPS · Anatomical · Mechanism · Certainty) operationalises this same intuition at the bedside [54]. The unresolved question has been why these mechanisms cluster differently in patients with otherwise identical structural pathology, and why apparently anatomically successful interventions still produce divergent IMMPACT outcomes.

The tri-level model offers a unifying interpretation. PSPS-T1 corresponds to the patient in whom the central and peripheral amplifiers (microglia–astrocyte priming via TLR4, biopsychosocial Z, and an inflammatory milieu favouring CMis un-silencing) reach symptomatic threshold without the need for a structural surgical insult—hence pain “persists despite optimal non-surgical management” in the absence of any operation. PSPS-T2 represents the same amplifier ensemble layered upon a surgical event that adds focal nociceptive afferentation, post-surgical DAMP release, and reinforced TLR4/IL-6/oncostatin signalling—explaining why surgery may fail to relieve pain or actively increase it. The same logic extends naturally to neighbouring diagnoses: chronic post-surgical pain (CPSS) is a special case of PSPS-T2 dominated by surgically induced peripheral nociception; complex regional pain syndrome and post-herpetic neuralgia represent peripheral CMis un-silencing as the dominant layer; fibromyalgia and nociplastic pain disorders represent dominance of the central–biopsychosocial layers with minimal peripheral driver; and painful diabetic neuropathy combines metabolic CMis un-silencing (via NGF up-regulation) with progressive central sensitisation. Differential treatment response across these conditions, often puzzling at the bedside, becomes interpretable once each diagnosis is mapped onto the dominant layer of the model rather than treated as an aetiological monolith.

For PALF specifically, this reading suggests that the procedure-specific calibration term β₀ᵒᵖ should be complemented by a layer-dominance descriptor analogous to the M (mechanism) component of the PAMC scale. A patient classified as PSPS-T2 with neuropathic dominance, high CSI, remote dermatomal pain, and elevated PALF Z is, in our framework, a patient in whom all three layers are simultaneously active—and consequently a patient in whom no single-layer intervention (regional block alone, SCS alone, or behavioural therapy alone) is mathematically expected to clear the IMMPACT threshold. The explicit articulation of this prediction is, in our view, the most actionable bedside contribution of the tri-level extension of PALF and a natural avenue for joint validation studies with the PSPS taxonomy steering committee.

### 4.6 Neurobiological synthesis: TLR4-DAMP signalling as the integrative axis of the tri-level model

The preceding two sections introduced the neurobiological elements piece by piece in service of specific clinical questions (the persistent alarm mode under impeccable technique; the PSPS-T1/T2 dichotomy). For coherence with the rest of the manuscript and to facilitate independent reading by basic-science colleagues, we provide here a self-contained synthesis of the molecular and cellular substrate underlying PALF.

#### Peripheral substrate—the un-silencing of CMis nociceptors

The recent molecular characterisation of human dermal mechano-insensitive C-fibres (CMis, sleeping nociceptors) by Körner, Lampert, Tripathy and Price [49] establishes that this ∼10% subpopulation of C-fibres co-expresses oncostatin-M receptor (OSMR), somatostatin (SST), and the low-threshold sodium channel Na_v_1.9. CMis are electrically silent in the basal state but undergo OSMR-driven un-silencing under inflammatory or neuropathic conditions. Once un-silenced, they fire spontaneously and acquire de novo mechanosensitivity, generating sustained ectopic discharge into the dorsal horn. This phenomenon was experimentally reproduced in healthy volunteers by intradermal oncostatin M, which elicited spontaneous pain and remote secondary mechanical hyperalgesia, closing the loop with the TMEM100-dependent un-silencing previously described by Lampert and colleagues [50]. CMis un-silencing is the persistent peripheral driver that regional anaesthesia cannot silence, because active C and Aδ fibres are not its substrate.

#### Cytokine bridge—OSMR-IL-6 family signalling.

Mwirigi and colleagues [52] demonstrate that OSMR is part of the IL-6 cytokine family (IL-6, LIF, OSM, CNTF) sharing the gp130 transducer, and that its upstream regulation overlaps with TLR4-DAMP signalling. The peripheral and central layers are therefore not parallel pathways but coupled compartments, with the OSMR-IL-6 axis acting as the cytokine bridge that conveys peripheral inflammatory information to spinal microglia and astrocytes.

#### Central substrate—TLR4 as the DAMP master switch

As reviewed in detail by García, Goicoechea, Molina-Álvarez and Pascual [46], TLR4 is expressed in microglia, astrocytes, dorsal-root-ganglion (DRG) sensory neurons and dorsal-horn projection neurons. It functions both as a PAMP receptor (LPS) and, more relevantly for chronic pain, as a DAMP receptor for HMGB1, S100 proteins, heat-shock proteins, fibronectin fragments, hyaluronic acid oligomers, and extracellular-matrix degradation products. Activation of TLR4 in spinal microglia drives NF-κB-dependent release of TNF-α, IL-1β, IL-6, BDNF, and CCL2, producing the well-characterised loss of GABAergic dorsal-horn inhibition (chloride-shift via KCC2 down-regulation) and the gain in NMDA-receptor-mediated central excitatory drive. TLR4 is also engaged non-stereoselectively by morphine and its glucuronide metabolites, which provides the molecular link between iatrogenic opioid exposure (β₄ in PALF) and persistent maladaptive sensitisation [46]. Polymorphisms in TLR4 (Asp299Gly, Thr399Ile) and in MyD88/TRIF effectors plausibly contribute to the extreme between-study heterogeneity (I²=99.96%) of the opioid OR.

#### Glial transition—the biphasic microglia-to-astrocyte cascade

Donnelly, Andriessen, Chen and colleagues [51] characterised the dorsal-horn cellular trajectory after peripheral nerve injury as a biphasic process: an early microglial phase (proliferation, activation, release of pro-inflammatory cytokines and BDNF) followed by a late astrocytic phase (reactive astrogliosis with sustained release of lipocalin-2, CXCL1 and IFN-α through STING signalling). The transition is not a relay but a coupled mechanism: cyclic GMP-AMP synthase (cGAS) detects cytosolic DNA released by stressed cells and activates STING, which amplifies type-I interferon signalling and locks the dorsal horn into a chronic, self-sustaining inflammatory state. STING is therefore a pharmacologically tractable node downstream of TLR4 that may explain why purely anti-microglial strategies (e.g. minocycline alone) often show inconsistent late-phase efficacy.

#### Confirmatory in-vivo evidence—the MIA osteoarthritis model

Ferreira-Gomes, García, Nascimento, Castro-Lopes, Pascual, Goicoechea and Lourença Neto [47] tested whether systemic blockade of TLR4 was sufficient to attenuate established nociception in a model with persistent peripheral structural pathology. In adult Wistar rats with intra-articular sodium monoiodoacetate (MIA, 2 mg) into the left knee, administration of the synthetic TLR4 antagonist TLR4-A1 (10 mg/kg i.p., days 14–28) significantly reduced movement- and loading-induced nociception (Knee-Bend and CatWalk) and reduced ATF-3 expression in L3- L5 DRG sensory neurons, without modifying cartilage degradation. The clinical analogue is direct: TLR4-A1 acts on the nociceptive amplification pathway rather than on the joint lesion, exactly the decoupling between structure and pain that characterises high-Z PALF patients with technically adequate targeting and persistent IMMPACT failure.

#### Therapeutic landscape and the loop-breaking pilot trial

The integrated TLR4-DAMP / OSMR-IL-6 / STING architecture has clinically available counterparts. Low-dose naltrexone (LDN, 4.5 mg/day) acts as a TLR4 antagonist and microglial modulator with an established safety profile in fibromyalgia and Crohn’s disease. Pentoxifylline (400 mg t.i.d.) is a non-selective phosphodiesterase inhibitor that suppresses TNF-α and IL-6 release from activated microglia and has decades of cardiovascular use. Minocycline acts as a broad microglial inhibitor. Anti-NGF biologics (tanezumab, fasinumab) and anti-OSMR antibodies (vixarelimab in development) target the peripheral CMis compartment. Selective Na_v_1.9 small-molecule modulators are entering phase I/II. Together these constitute a rational, layer-targeted pharmacological menu, in contrast with the current empirical practice of stacking analgesics across mechanisms.

On this basis, a pilot loop-breaking randomised double-blind placebo-controlled trial of LDN 4.5 mg/day plus pentoxifylline 400 mg t.i.d. for four weeks pre-procedure—in patients with high-Z PALF, chronicity > 12 months, CSI ≥ 40, and a planned interventional procedure—is being planned in collaboration between the HUNSC Pain Unit (Tenerife) and the URJC pharmacology group (Goicoechea laboratory, Madrid). Primary outcome: IMMPACT response rate at six months; secondary outcomes: change in HMGB1, IL-6, OSMR, and CRP, and calibration of Z_periph against clinical response. This trial is designed to provide the first prospective test of the layer-dominance logic of the tri-level model and the first clinical anchor for the latent neurobiological coefficient that PALF v19.1 currently treats as mechanistic background rather than as an explicit predictor.

In summary, the neurobiological architecture supporting PALF is not a rhetorical appendix but a concrete, falsifiable mechanism with identified molecular nodes (TLR4, OSMR, STING, Na_v_1.9), measurable proxies (CRP, HbA1c, CSI, Brush Allodynia Test, IL-6, HMGB1), and layer-specific therapeutic candidates. The PALF composite is the population-level summary statistic of the Z biopsychosocial layer; the tri-level model adds the peripheral and central neurobiological layers; and the loop-breaking pilot trial provides the prospective probe by which the framework can be moved from a structured hypothesis toward an empirically validated tool.

#### Mechanistic triangulation as a methodological instrument (v19.3 addition)

Following Lawlor, Tilling and Davey Smith (2016, *Int J Epidemiol* 45:1866-1886) and the updated Bradford-Hill framework, we treat the convergence of evidence across heterogeneous study designs not as rhetorical reinforcement but as a formal source of inferential credibility complementary to AMSTAR 2 and GRADE. Each PALF risk domain is supported by three orthogonal lines of evidence whose biases differ structurally: (i) **epidemiological**, the systematic-review odds ratios that populate β_i and that are subject to confounding, selection bias, and outcome heterogeneity; (ii) **molecular**, the TLR4-DAMP signalling axis with identified ligands (HMGB1, S100 proteins, fragmented fibronectin) and measurable downstream effectors (NF-κB activation, IL-1β/IL-6/TNF-α release, glial activation), where biases are largely experimental (cell line variability, antibody specificity); and (iii) **translational/in vivo**, animal models of nerve injury and persistent allodynia (TLR4 knockouts, S100A8/A9 modulation, DAMP-targeted interventions), where biases are species-extrapolation and dose-response dependent. When epidemiological associations, mechanistic plausibility, and experimental reversibility converge on the same direction (sleep disruption → cytokine elevation → glial sensitisation → chronic pain; opioid exposure → mu-receptor downregulation + TLR4 activation → iatrogenic hyperalgesia), the joint probability that all three independent biases have aligned spuriously becomes vanishingly small. This triangulation does not generate new estimates; it constrains the interpretation of existing β_i by *raising the prior* that the population-level association reflects a mechanistically plausible causal structure rather than residual confounding. In the language of the GRADE framework, mechanistic triangulation operates analogously to a *biological gradient* and *coherence* upgrade for observational evidence (Guyatt et al. 2008; Hill 1965), and we apply it explicitly in Supplement S9 to justify upgrading certainty for Domains 1, 2 and 4. For PALF, triangulation is therefore a tertiary instrument alongside AMSTAR 2 (which evaluates the quality of the syntheses themselves) and PROBAST (which evaluates the composite as a prediction tool); together, the three instruments map onto the three sources of inferential uncertainty — quality of the inputs, quality of the integration, and quality of the causal claim.

### 4.4 Limitations

**First (methodological validity):** The PALF assembles separately estimated ORs from different populations, outcomes, and covariate adjustment sets. This violates the assumption that components of a composite score measure the same latent construct. The non-collapsibility of the odds ratio means that marginally estimated ORs are not comparable to conditionally estimated ORs, and combining them produces a coefficient without a strict statistical interpretation in any single probability model [Greenland et al., 1999]. The ecological inference fallacy further limits extrapolation from population-level ORs to individual-level prediction.

**Second (outcome heterogeneity):** The opioid and BZD domains predict healthcare utilization (prolonged opioid use), not chronic pain. The primary three-domain analysis (Section 3.9) addresses this structurally by excluding the pharmacological domains; the expanded six-domain composite carries this limitation.

**Third (covariate adjustment heterogeneity):** Source studies adjusted for different covariates (Table S1), introducing potential collider bias and omitted variable bias when combining their ORs.

**Fourth (extreme opioid heterogeneity):** I²=99.96% renders the opioid OR a rough central tendency indicator. The REML/KH CI (1.87–15.13) is 4.4x wider than the DL CI.

**Fifth (constant-slope assumption):** The PALF assumes risk factor effects are constant across procedures—biologically untested and likely wrong.

**Sixth (tautological simulation):** The Monte Carlo AUCs are mathematical properties, not empirical validation. The PALF has zero empirical validation.

**Seventh (AMSTAR 2 quality):** Theunissen et al. rated Low confidence, limiting the catastrophizing coefficient.

**Eighth (publication bias assessment):** Egger’s test is underpowered for domains with k<10. PET-PEESE provides additional reassurance for sleep and catastrophizing but cannot be applied to the opioid domain due to extreme heterogeneity.

**Ninth (source meta-analysis age):** The metabolic (Shiri 2010) and catastrophizing (Theunissen 2012) coefficients derive from meta-analyses 14+ years old. Newer data may modify these estimates.

**Tenth (binary input limitation):** The binary (0/1) input specification discards continuous information from clinical instruments. This was a necessary constraint to match the binary contrasts reported in the source meta-analyses, but it means the PALF cannot capture dose-response relationships.

**Eleventh (hypothetical tri-level extension):** Z_periph and the tri-level extension proposed in §4.2-§4.3 are hypothesis-generating and have not been prospectively validated. The proposed pilot loop-breaking trial (LDN + pentoxifylline) and the HUNSC registry will provide the first calibration of layer-dominance descriptors and the λ_{M,proc} adjustment.

### 4.5 Future Directions

Prospective validation with individual patient data (≥500 patients per procedure) is the essential next step. Priorities: (a) simultaneous multivariate estimation of all coefficients; (b) calibration and discrimination against 3/6-month outcomes; (c) subgroup/meta-regression for the opioid domain; (d) comparison of binary vs. continuous input models; (e) formal causal analysis using Mendelian randomization for metabolic and sleep domains.

## 5. CONCLUSIONS

This umbrella review demonstrates that five biopsychosocial risk factor domains and one pharmacological domain are each associated with chronic pain or related treatment outcomes, with meta-analytic ORs ranging from 1.39 to 5.32. The PALF is presented here as a structured analytical framework for future prospective validation, not as a clinical prediction tool ready for deployment. We make no claim to clinical validity in the absence of prospective individual-patient-data validation.

The primary three-domain composite (sleep, catastrophizing, metabolic) achieves outcome homogeneity—all three source meta-analyses target chronic pain or pain chronification—and carries moderate PROBAST risk of bias. Its delta method confidence intervals span 10–15 percentage points, frequently crossing tier boundaries (Figure 8). The secondary expanded six-domain composite adds the pharmacological and smoking domains, broadening clinical coverage at the cost of outcome heterogeneity and high PROBAST risk of bias; its confidence intervals span 12–18 percentage points.

The PALF provides formal uncertainty propagation revealing that individual patient risk tier assignments are substantially uncertain—the framework’s principal contribution is making this uncertainty explicit rather than hiding it behind a single point estimate. The wide confidence intervals, outcome heterogeneity across domains, extreme heterogeneity in the opioid domain (I²=99.96%), and absence of any empirical validation collectively preclude clinical application. The TRIPOD+AI checklist (Supplementary S6) and PROBAST assessment (Supplementary S7) document the development-phase status of the PALF and the specific steps required before any deployment could be considered. The framework serves as a hypothesis-generating structure and a design template for future prospective individual-patient-data validation studies.

## Data Availability

All data are derived from published studies. Monte Carlo code (R, seed=42), analysis scripts, and extracted data are available at https://osf.io/5tqcz.

## Ethics

No ethical approval required (analysis of published aggregate data).

**Protocol Registration:** PROSPERO CRD420261360881; OSF https://osf.io/5tqcz.

## Data Availability

All extracted effect sizes, R/Python analysis code, search strategies, PRISMA-O / TRIPOD+AI / PROBAST checklists, and figures are deposited at the Open Science Framework (OSF DOI 10.17605/OSF.IO/5TQCZ; https://osf.io/5tqcz). The protocol is registered on PROSPERO (CRD420261360881). This manuscript expands medRxiv preprint 10.64898/2026.03.22.26348998.

https://doi.org/10.17605/OSF.IO/5TQCZ

## SUPPLEMENTARY MATERIAL

**Table S1.**
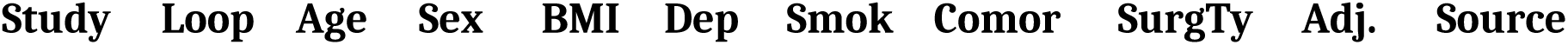
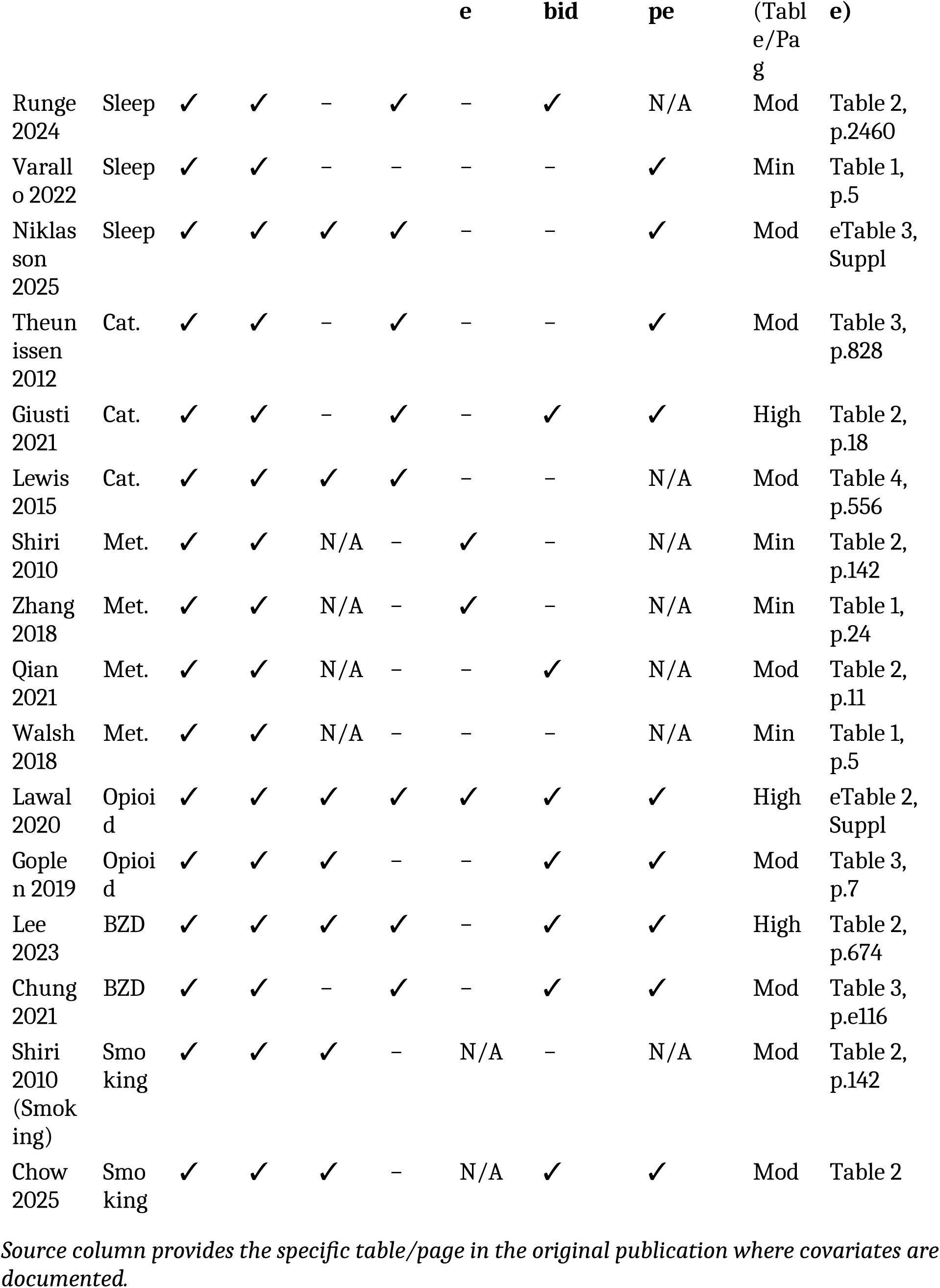
Covariate Adjustment Matrix with Source Citations.

**Table S2.**
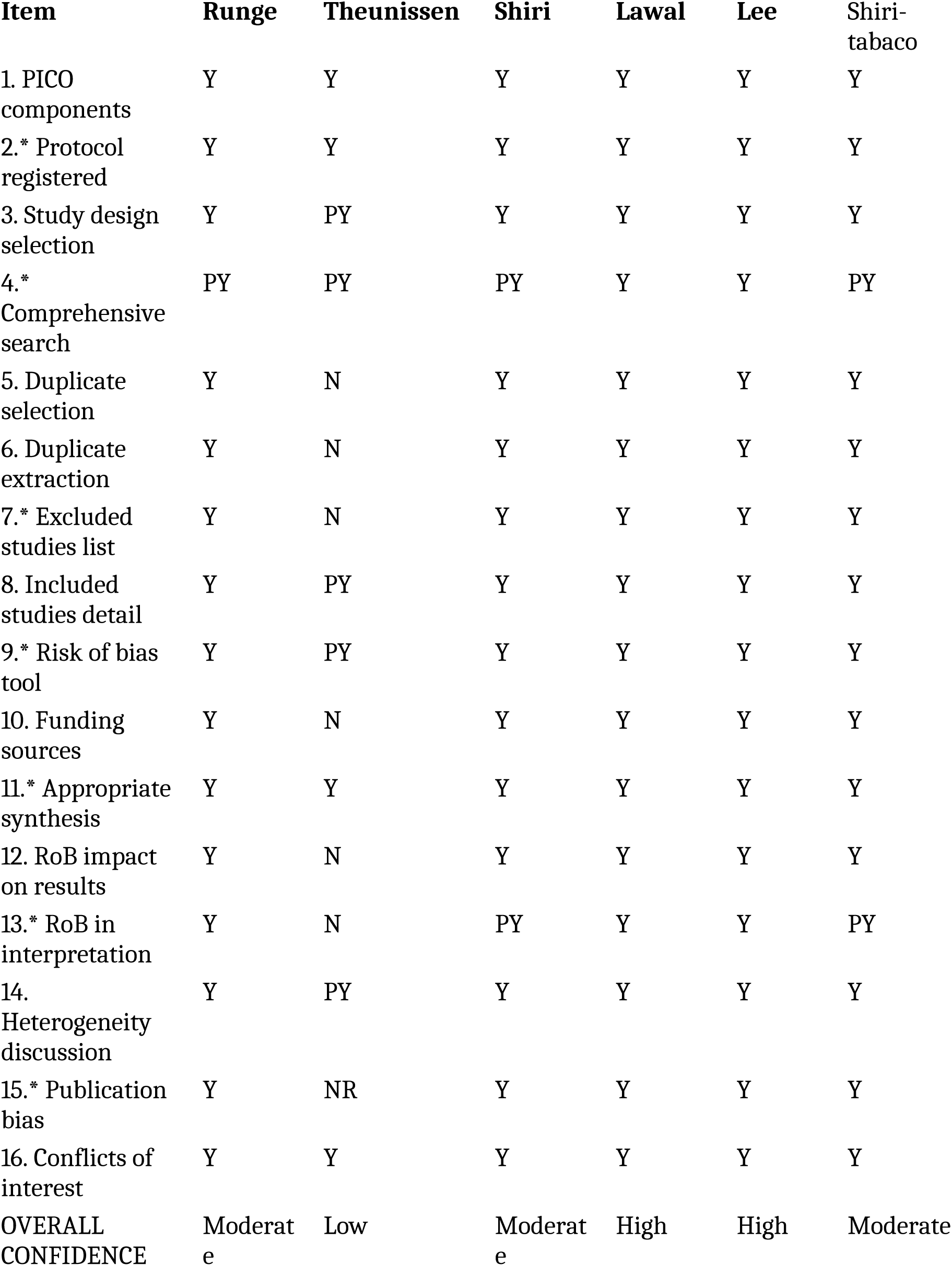
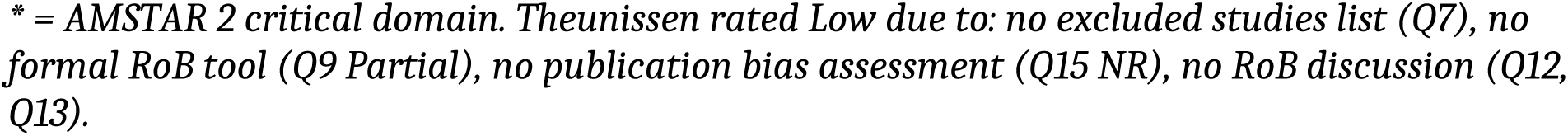
AMSTAR 2 Quality Assessment — Full 16-Item Assessment.

| Item | Runge | Theunissen | Shiri | Lawal | Lee | Shiri-tabaco |
| --- | --- | --- | --- | --- | --- | --- |
| 1. PICO components | Y | Y | Y | Y | Y | Y |
| 2.* Protocol registered | Y | Y | Y | Y | Y | Y |
| 3. Study design selection | Y | PY | Y | Y | Y | Y |
| 4.* Comprehensive search | PY | PY | PY | Y | Y | PY |
| 5. Duplicate selection | Y | N | Y | Y | Y | Y |
| 6. Duplicate extraction | Y | N | Y | Y | Y | Y |
| 7.* Excluded studies list | Y | N | Y | Y | Y | Y |
| 8. Included studies detail | Y | PY | Y | Y | Y | Y |
| 9.* Risk of bias tool | Y | PY | Y | Y | Y | Y |
| 10. Funding sources | Y | N | Y | Y | Y | Y |
| 11.* Appropriate synthesis | Y | Y | Y | Y | Y | Y |
| 12. RoB impact on results | Y | N | Y | Y | Y | Y |
| 13.* RoB in interpretation | Y | N | PY | Y | Y | PY |
| 14. Heterogeneity discussion | Y | PY | Y | Y | Y | Y |
| 15.* Publication bias | Y | NR | Y | Y | Y | Y |
| 16. Conflicts of interest | Y | Y | Y | Y | Y | Y |
| OVERALL CONFIDENCE | Moderate | Low | Moderate | High | High | Moderate |

\* = AMSTAR 2 critical domain. Theunissen rated Low due to: no excluded studies list (Q7), no formal RoB tool (Q9 Partial), no publication bias assessment (Q15 NR), no RoB discussion (Q12, Q13).

**Table S3.** GRADE Evidence Profile — Detailed Assessment.

| Domain | RoB | Inconsistency | Indirectness | Imprecision | Pub. Bias | Upgrade | GRADE |
| --- | --- | --- | --- | --- | --- | --- | --- |
| Sleep | Not serious | Not serious (I<sup>2</sup>=51%) | Not serious (direct pain) | Not serious (k=16) | None (Egger p=0.21; PET NS) | None | Moderate |
| Catastroph. | Serious (AMSTAR Low) | Not serious (I<sup>2</sup>=0%; low power) | Serious (postsurg only) | Not serious | None (Egger p=0.84; PET NS) | Large effect | Low |
| Metabolic | Not serious | Not reported (Shiri) | Not serious | Not serious (k=33) | Not assessed (relied on source) | None | Moderate |
| Opioid | Not serious | Very serious (I<sup>2</sup>=99.96%) | Serious (opioid use≠pain) | Not serious (k=33) | Not assessed (I<sup>2</sup> precludes test) | Large effect | Very Low |
| BZD | Not serious | Not serious | Serious (opioid use≠pain) | Serious (wide REML CI) | NS (Egger p=0.12; underpowered) | None | Low |
| Smoking | Not serious | Serious (I<sup>2</sup>≈59%) | Not serious (chronic LBP) | Not serious (k=8) | Not assessed (k=8 underpowered) | None | Moderate |

Opioid domain downgraded to Very Low (2 serious, 1 very serious downgrades). Footnotes: RoB=risk of bias; NS=non-significant.

**Table S4.**
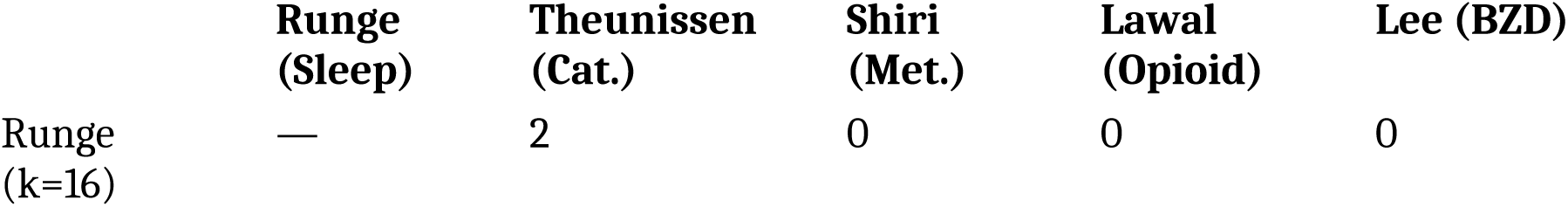
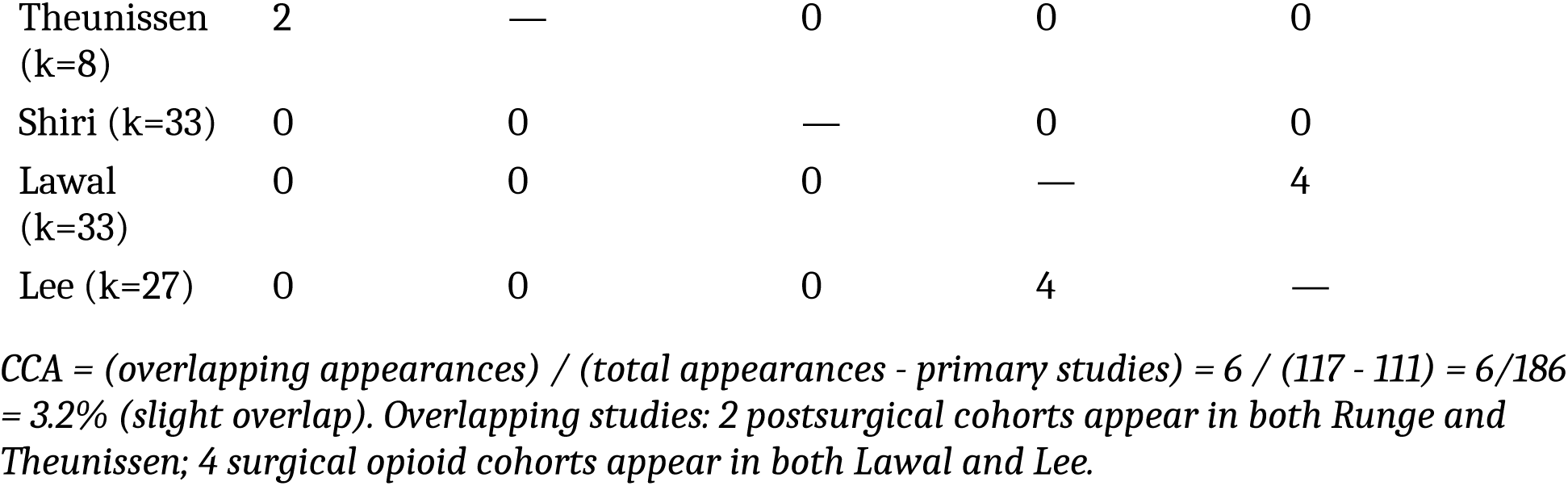
Primary Study Overlap Matrix and CCA Calculation.

|  | Runge (Sleep) | Theunissen (Cat.) | Shiri (Met.) | Lawal (Opioid) | Lee (BZD) |
| --- | --- | --- | --- | --- | --- |
| Runge (k=16) | — | 2 | 0 | 0 | 0 |
| Theunissen (k=8) | 2 | — | 0 | 0 | 0 |
| Shiri (k=33) | 0 | 0 | — | 0 | 0 |
| Lawal (k=33) | 0 | 0 | 0 | — | 4 |
| Lee (k=27) | 0 | 0 | 0 | 4 | — |

CCA = (overlapping appearances) / (total appearances - primary studies) = 6 / (117 - 111) = 6/186 = 3.2% (slight overlap). Overlapping studies: 2 postsurgical cohorts appear in both Runge and Theunissen; 4 surgical opioid cohorts appear in both Lawal and Lee.

### Supplementary Material S4. PRISMA-O Checklist

A completed PRISMA for Overviews of Reviews (PRISMA-O) checklist is provided as a separate supplementary file. The checklist follows the PRISMA-O extension, including items specific to umbrella reviews: overlap assessment (Item 17b), quality of included reviews (Item 14b), and certainty of evidence across reviews (Item 22b).

### Supplementary Material S5. Full Search Strategies

Complete search strings for each risk factor domain across PubMed/MEDLINE, Scopus, and Cochrane Library are provided as a separate supplementary file. Search strategies were developed with a medical librarian and refined iteratively.

### Supplementary Material S6 — TRIPOD+AI Checklist

*Reference: Collins GS, Moons KGM, Dhiman P, et al. TRIPOD+AI statement: updated guidance for reporting clinical prediction models that use regression or machine learning methods. BMJ. 2024;385:e078378.* doi:10.1136/bmj-2023-078378.

**Rationale for N/A designations:** Items marked N/A with the notation “Composite score derived analytically from published meta-analytic estimates; no de novo training cohort” reflect the parameter-assembly nature of the PALF. The PALF assembles published meta-analytic log-ORs into a logistic equation using the delta method; it does not train a model on individual patient data. Accordingly, items requiring a training cohort, internal validation, model updating, or deployment infrastructure are not applicable at this development phase.

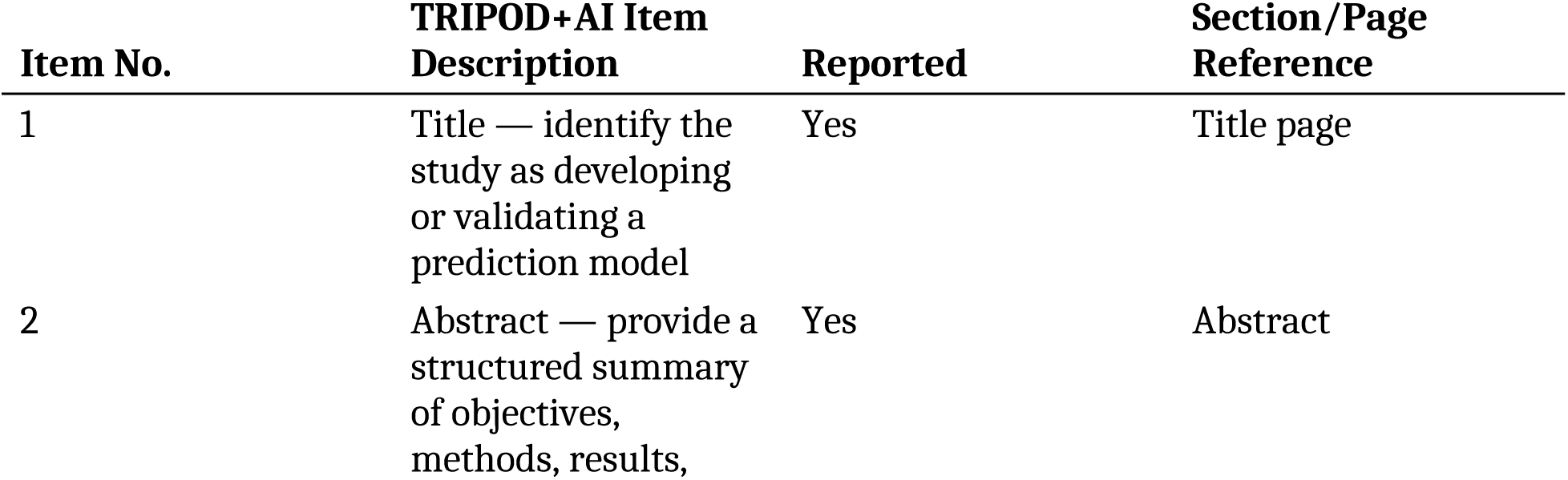
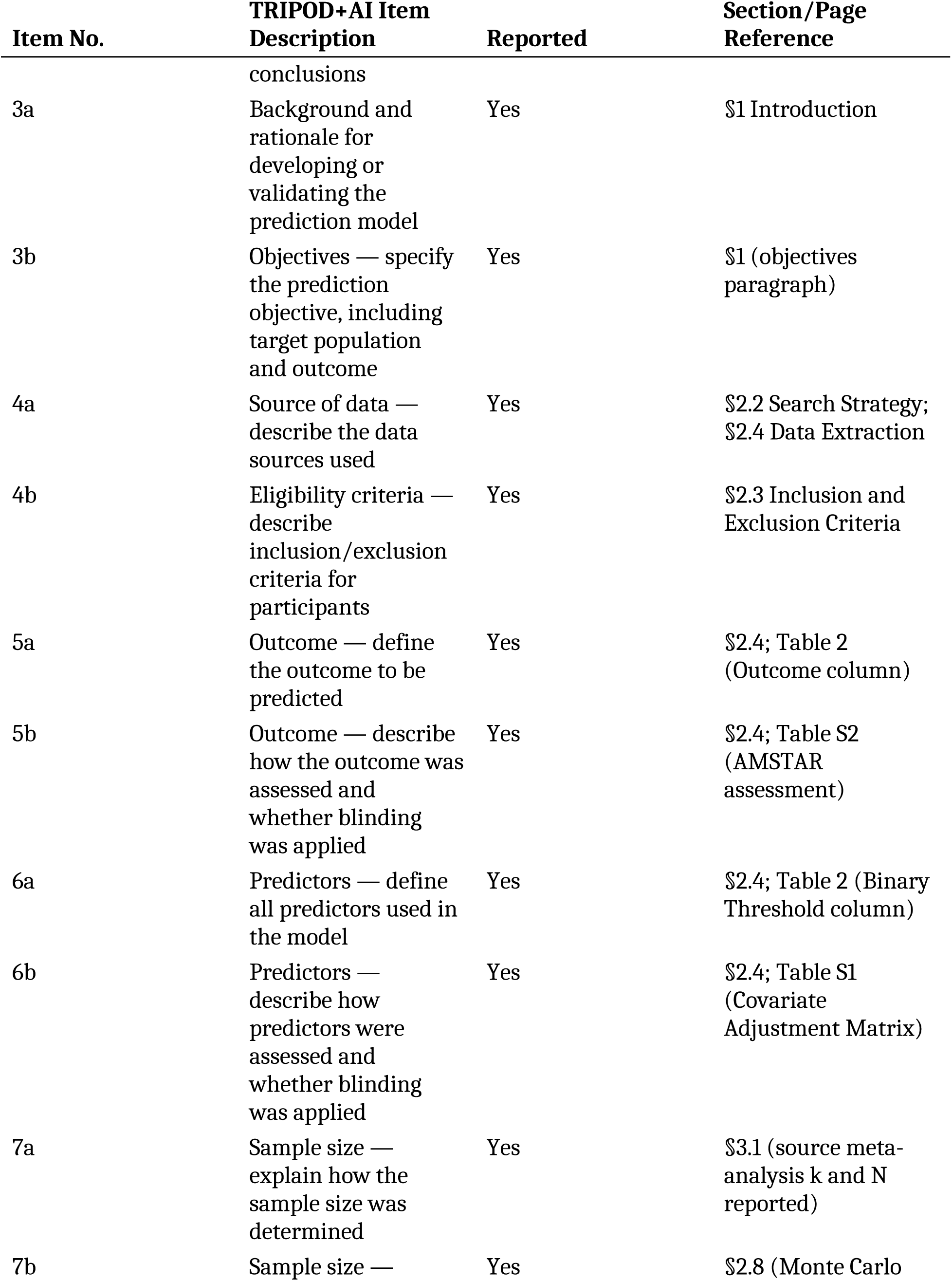
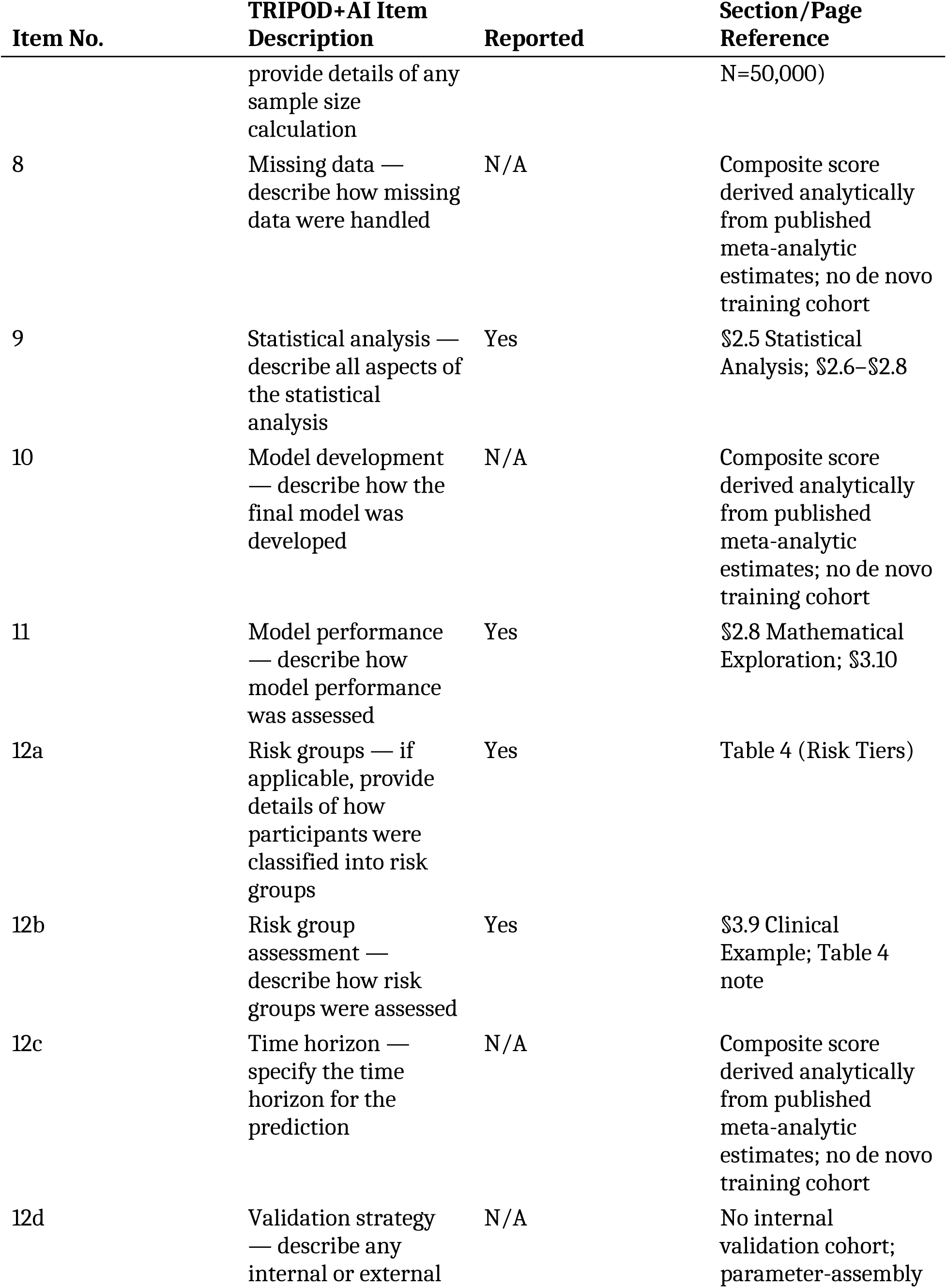
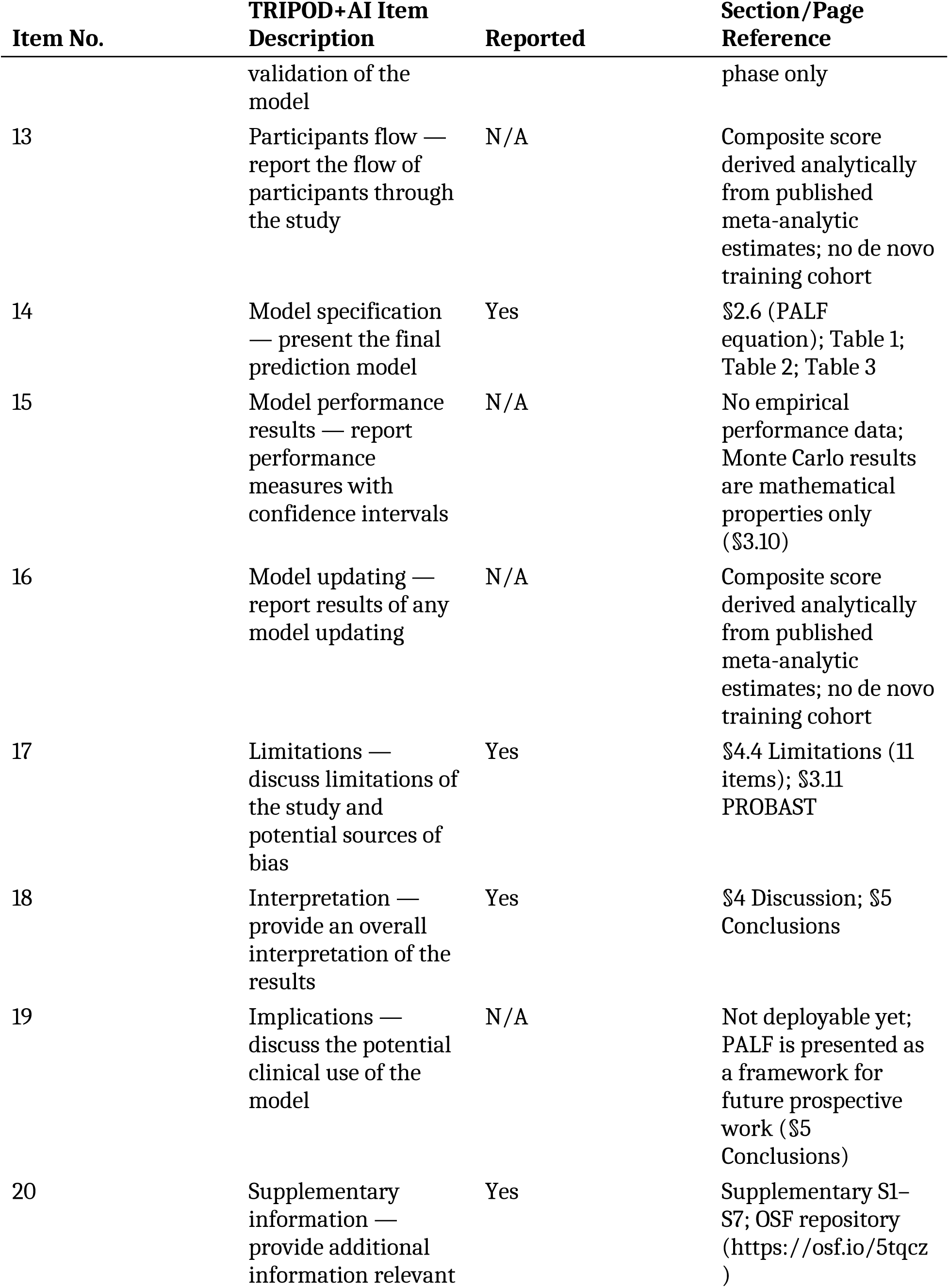
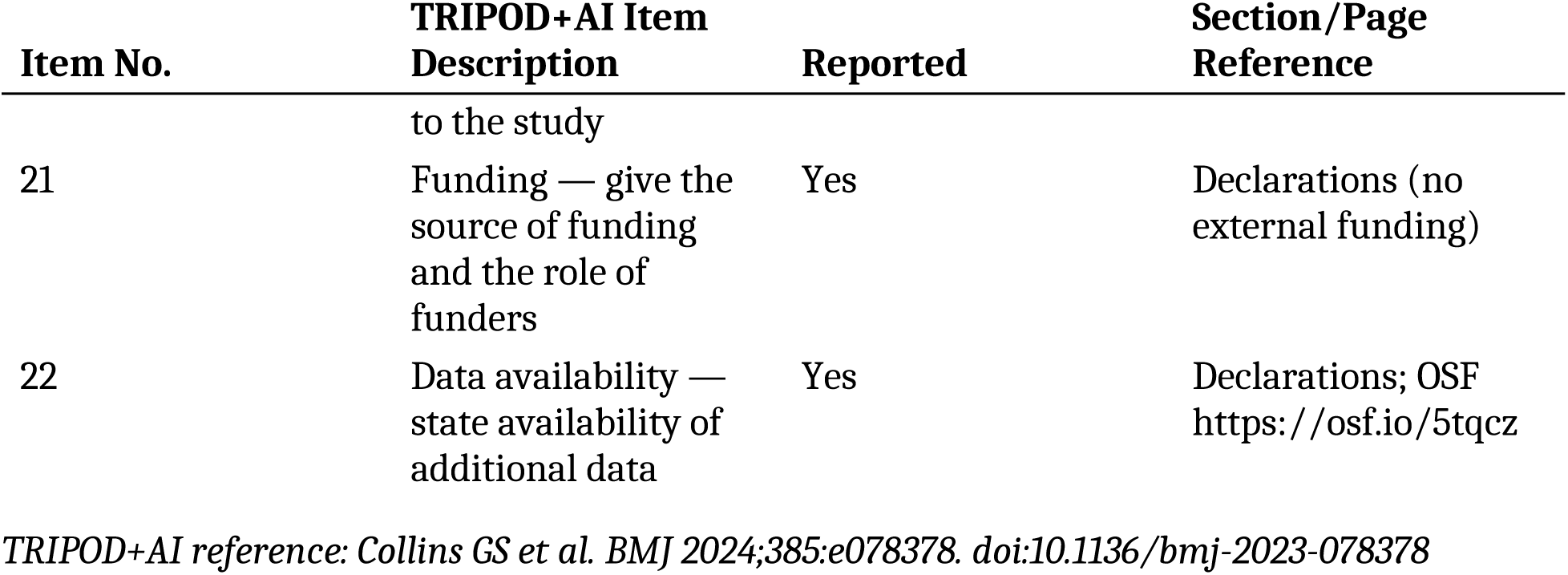

### Supplementary Material S7 — PROBAST Risk of Bias Assessment

*Reference: Wolff RF, Moons KGM, Riley RD, et al. PROBAST: A Tool to Assess the Risk of Bias and Applicability of Prediction Model Studies. Ann Intern Med. 2019;170(1):51–58.* doi:10.7326/M18-1376.

**Note on terminology.** This supplement uses **PROBAST D1–D4** to refer to the four PROBAST tool domains (Participants, Predictors, Outcome, Analysis). The phrase **PALF Domain X** (1–6) refers to the six predictor domains of the PALF composite (Sleep, Catastrophizing, Metabolic, Opioid, BZD, Smoking). The two domain systems are conceptually orthogonal and should not be conflated.

**PROBAST was applied separately to the primary three-domain composite (sleep + catastrophizing + metabolic) and the secondary expanded six-domain composite (adding opioid, BZD, smoking).** The key distinction between the two composites for PROBAST purposes is **PROBAST D3 (Outcome)**: the three-domain composite is restricted to pain or pain-chronification outcomes; the expanded six-domain composite introduces pharmacological utilization outcomes that represent a different construct.

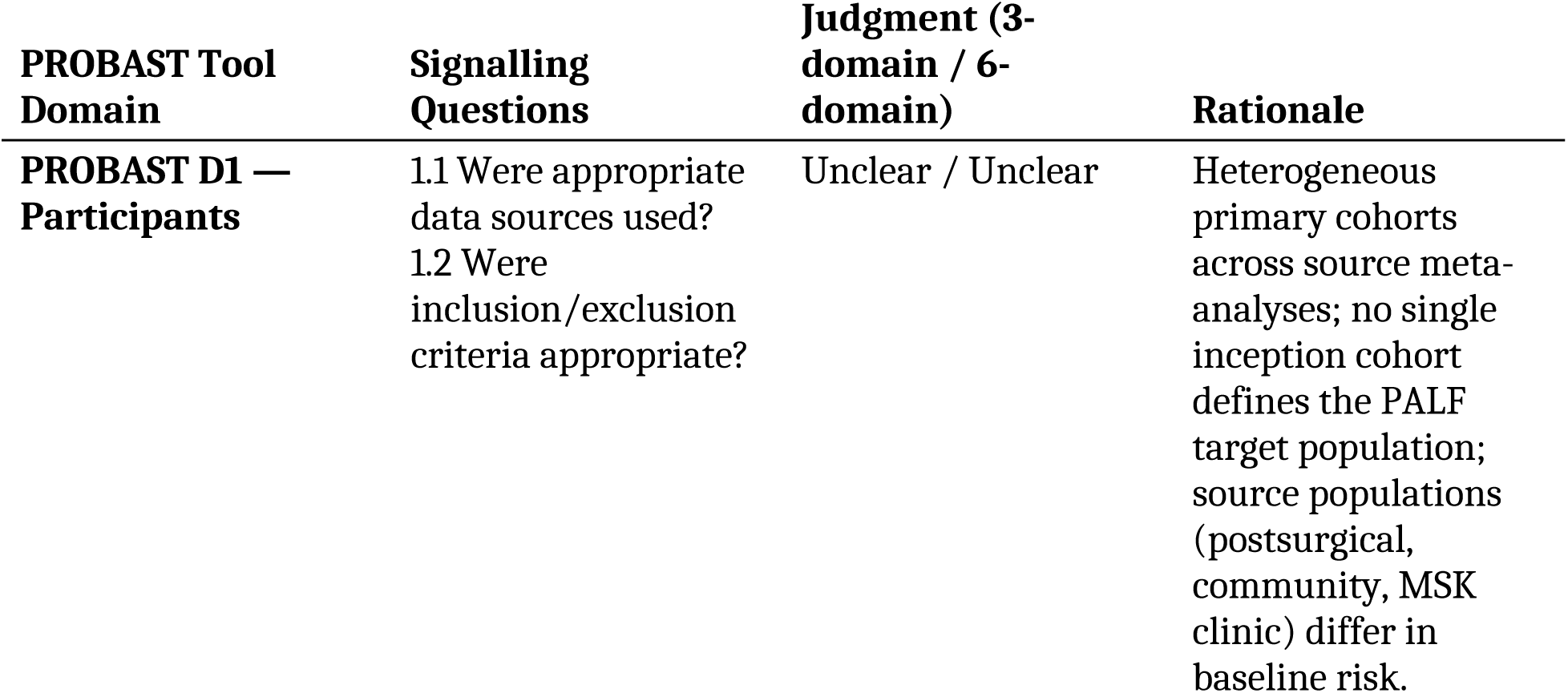
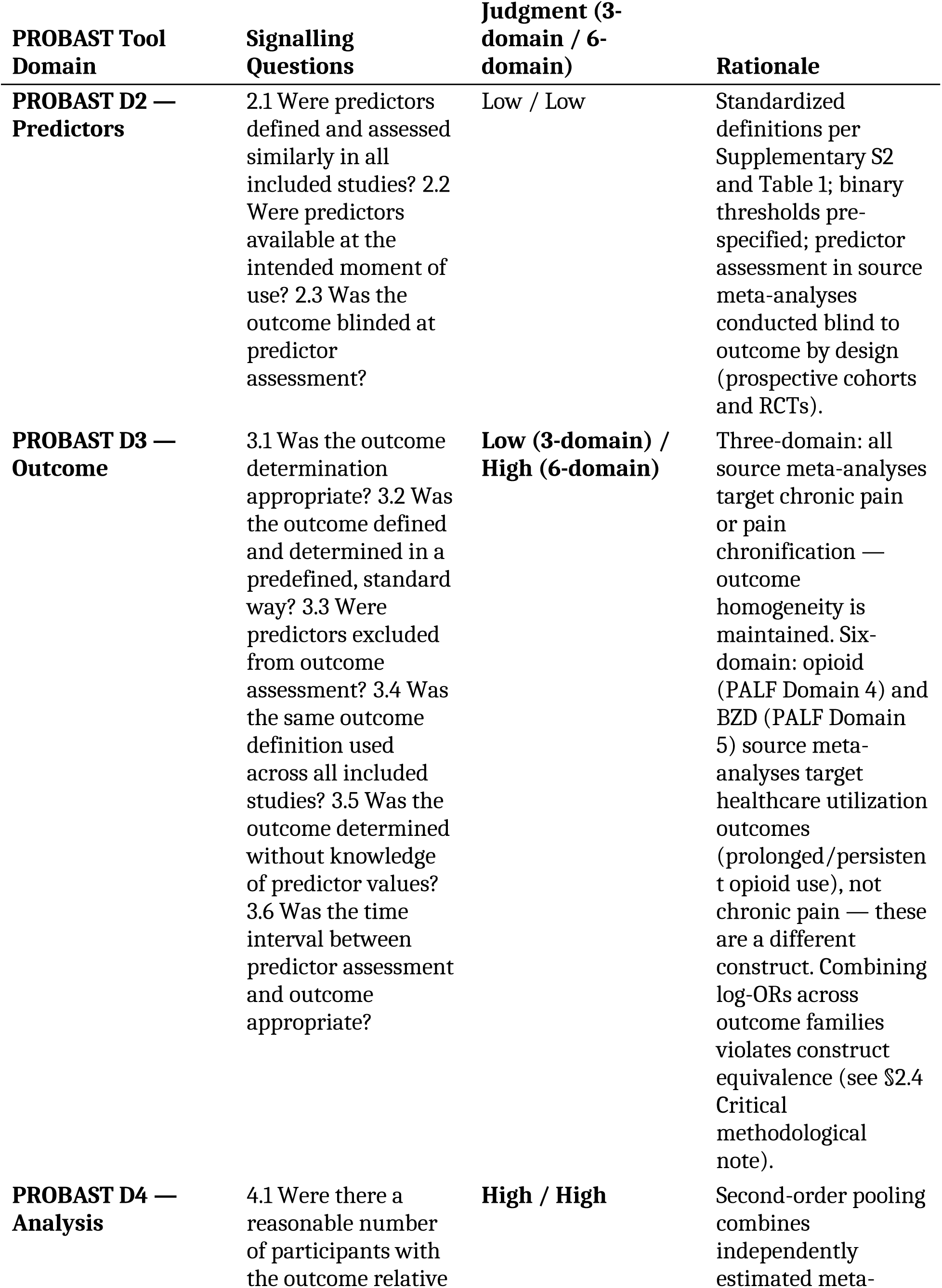
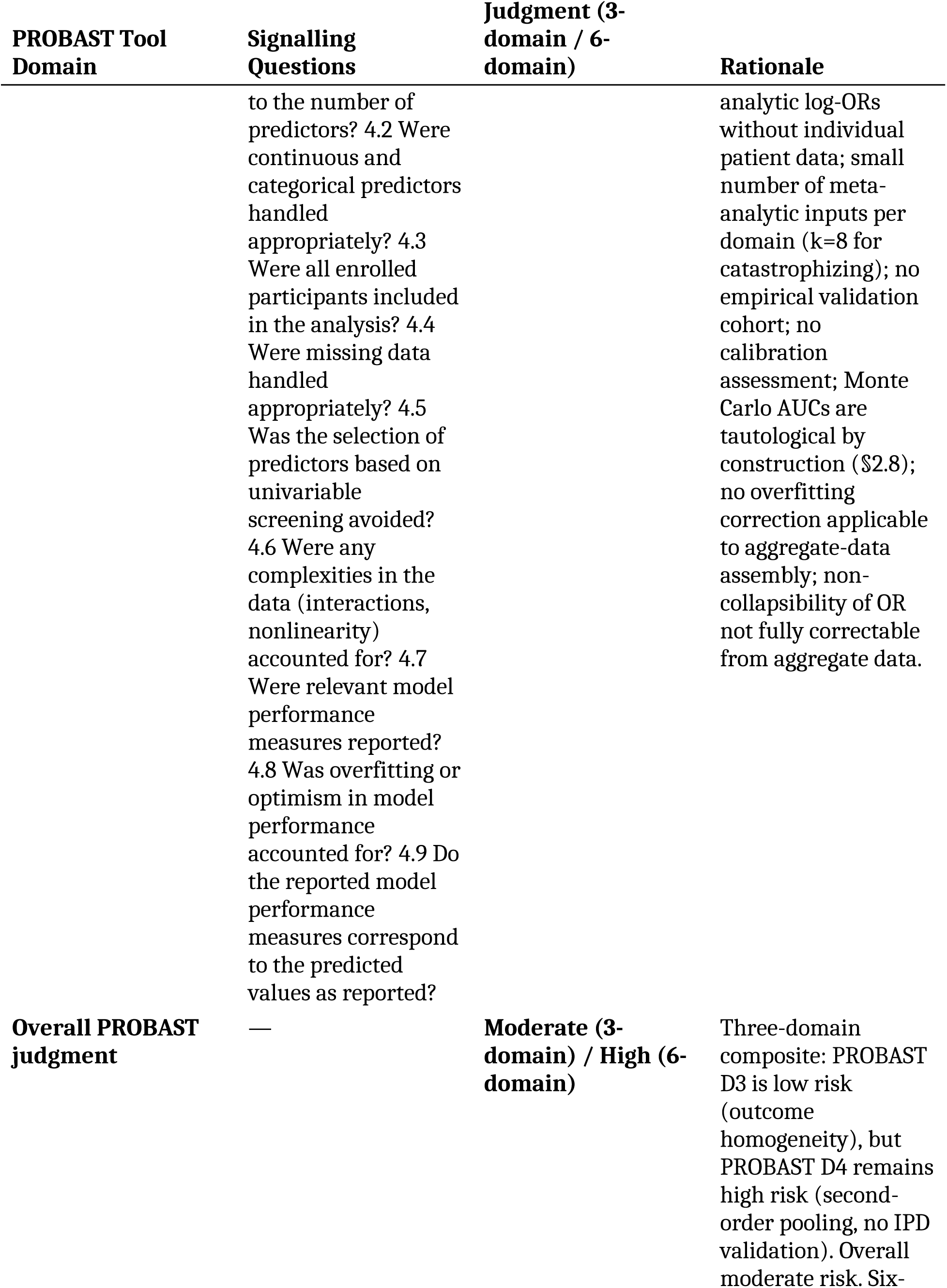
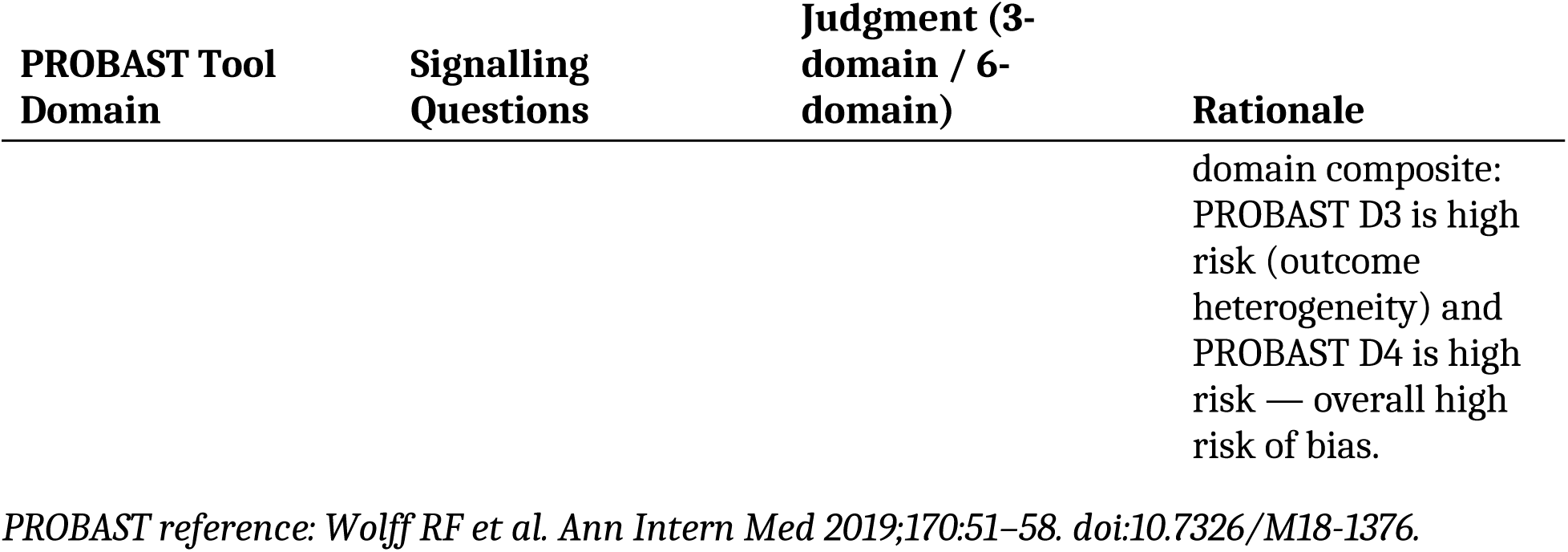

