## Supplemental Data 1 for "Quantifying Biopsychosocial Risk Factor Domains for Chronic Pain Treatment Outcomes: An Umbrella Review with De Novo Meta-Analyses, Formal Uncertainty Propagation, and the Pain Amplifier Loop Framework (PALF)"

*Companion to PALF manuscript v19.5.3 (6 May 2026).*

Authors: Javier Arranz-Durán, MD, FIPP, EDRA · Sofía Perera Monje, BSc

PROSPERO: CRD420261360881 (status: registered, pending CRD publication review)

*Status: DRAFT — pending validation by an independent second reviewer*

Tool: GRADE (Guyatt et al., BMJ 2008;336:924-926; updated GRADE handbook 2013)

**1. Purpose**

This supplement applies the GRADE (Grading of Recommendations Assessment, Development and Evaluation) framework to the six β_i sources of the PALF model, assessing the certainty of the evidence for each risk factor → chronic pain association. The GRADE certainty rating complements the AMSTAR 2 rating (Table S2 of the main manuscript): whereas AMSTAR evaluates the methodological quality of the systematic review, GRADE evaluates the strength of the causal inference about the outcome.

Rating policy: conservative — downgrades are applied whenever the textual evidence is ambiguous; upgrades require three simultaneous criteria (large effect + dose-response + plausible mechanism) or formal mechanistic triangulation (Lawlor 2016).

Standard starting point for observational evidence: Low (★★). Standard starting point for RCTs: High (★★★★).

All PALF sources are observational (cohort, cross-sectional, or case-control studies synthesised in meta-analyses); all start at Low and are adjusted using the 5 downgrading criteria and the 3 upgrading criteria.

**2. GRADE criteria applied**

**2.1 Five downgrading criteria**

| **Criterion** | **Definition** |
| --- | --- |
| Risk of bias | Risk of bias in the included studies (≈ AMSTAR 2 + RoB of primary studies) |
| Inconsistency | Unexplained between-study heterogeneity (high I² without a clear cause) |
| Indirectness | Differences between PICO of the meta-analysis and the PALF context (lumbar, post-procedure, chronic pain ≥ 3 months) |
| Imprecision | Wide 95% CI, few events, or few studies (small k, small N) |
| Publication bias | Funnel plot asymmetry or absence of formal assessment |

**2.2 Three upgrading criteria (observational only)**

| **Criterion** | **Definition** |
| --- | --- |
| Large effect | Pooled OR ≥ 2 (modest) or ≥ 5 (large) |
| Dose-response gradient | Documented dose-response relationship |
| Plausible confounders all biased toward null | Mechanistic triangulation (Lawlor 2016) suggesting that uncontrolled confounders would bias toward the null |

**2.3 GRADE certainty levels**

- High (★★★★): we are very confident that the estimate reflects the true effect
- Moderate (★★★): moderately confident; the true effect is likely close to the estimate
- Low (★★): limited confidence; the true effect may be substantially different
- Very low (★): very little confidence; the true effect is likely substantially different from the estimate

**3. GRADE Summary of Findings table — six PALF domains**

| **Domain** | **β_i** | **Source** | **OR (95% CI)** | **k / N** | **RoB** | **Inconsist.** | **Indirect.** | **Imprec.** | **Pub. bias** | **Upgrade** | **GRADE** |
| --- | --- | --- | --- | --- | --- | --- | --- | --- | --- | --- | --- |
| 1 — Sleep | β₁ = 0.329 | Runge 2024 [Ref 12] ⚠ PDF to verify | 1.39 (1.21–1.59) | k=16 / — | Serious (-1) | No | No | No | No | +1 (TLR4-DAMP mechanism) | ★★ Low |
| 2 — Catastrophising | β₂ = 0.742 | Theunissen 2012 [Ref 15] | 2.10 (1.49–2.95) | k=15 (total) / 7 (subgroup) / 5,046 | Very serious (-2) AMSTAR Critically Low | No | Serious (-1) outcome CPSP not identical to PSPS-T2 | Serious (-1) wide CI | Serious (-1) no funnel/Egger | +1 (large effect OR ≥ 2) | ★ Very Low |
| 3 — Metabolic / Obesity | β₃ = 0.358 | Shiri 2010 Am J Epidemiol [Ref 19] ⚠ PDF missing | 1.43 (1.28–1.60) | k=33 / — | Serious (-1) (assumed pre-PROSPERO) | No | No | No | No (assumed) | +1 (IL-6/TNF-α mechanism) | ★★ Low (provisional) |
| 4 — Preoperative opioids | β₄ = 1.672 | Lawal 2020 [Ref 23] | 5.32 (2.94–9.64) | k=14 / 1,922,743 | No (-0) AMSTAR Moderate | Serious (-1) high I² | No | No | No | +2 (very large effect OR ≥ 5 + iatrogenic mechanism) | ★★★ Moderate |
| 5 — Preoperative BZD | β₅ = 0.571 | Lee 2025 [Ref 55, label to be corrected] | 1.77 (1.53–2.05) | k=6 / part of 27 | No (-0) AMSTAR Moderate | No | Serious (-1) US opioid-naïve population; post-2019 search | No | No | +1 (opioid–BZD cross-tolerance mechanism) | ★★★ Moderate |
| 6 — Smoking | β₆ = 0.582 | Shiri 2010 Am J Med [Ref 44] (primary) + Dai 2021 [Ref 43] (confirmatory) | 1.79 (1.27–2.50) [chronic LBP] / 1.23 (1.09–1.40) [broad CMP] | k=8 (Shiri); k=32 (Dai) / 296,109 (Dai) | Serious (-1) (Shiri Critically Low) partly offset by Dai Moderate | Serious (-1) different ORs (1.79 vs 1.23) by outcome | Serious (-1) Dai uses broad CMP | No | No (Shiri trim-and-fill applied) | +1 (ischaemic-oxidative mechanism) | ★★ Low |

**4. Detailed certainty justification by domain**

**4.1 Domain 1 — Sleep (β₁) — Low certainty (★★)**

Evidence status: the current PDF for Ref 12 (Rosenström 2024) does not match the citation Runge 2024 on sleep and chronic pain. The OR 1.39 (1.21–1.59) reported in Table 2 of the manuscript requires documentary verification before final submission.

Assuming Runge 2024 is a SR+MA with Moderate AMSTAR (to be confirmed):

- Starting point: Low (observational)
- Downgrade -1 for risk of bias (assumed Moderate–Low AMSTAR because the correct PDF has not been located)
- No further downgrade for inconsistency, indirectness, imprecision, or publication bias (assumed)
- Upgrade +1 for plausible mechanism: TLR4–DAMP triangulation (sleep deprivation → IL-6, TNF-α, HMGB1 → TLR4 activation → glial sensitisation → chronification). Convergence between epidemiological evidence (OR 1.39), molecular evidence (elevated HMGB1/IL-6 under experimental deprivation) and translational evidence (chronic sleep-deprivation murine models develop mechanical allodynia).
- Net result: Low (★★). Rating may be revised after PDF verification.

**4.2 Domain 2 — Catastrophising (β₂) — Very low certainty (★)**

Justification:

- Starting point: Low (observational)
- Downgrade -2 for risk of bias: AMSTAR 2 Critically Low (Theunissen 2012; pre-PROSPERO, search in 2 databases, no list of excluded studies, no funnel plot)
- Downgrade -1 for indirectness: meta-analysis outcome is generic CPSP; PALF applies to PSPS-T2 (Persistent Spinal Pain Syndrome Type 2 specific to lumbar post-procedural context), a more restrictive subpopulation
- Downgrade -1 for imprecision: wide 95% CI (1.49–2.95) with k=7 in the catastrophising subgroup
- Downgrade -1 for publication bias: Theunissen 2012 reported neither funnel plot nor Egger/Begg tests
- Upgrade +1 for large effect: OR 2.10 meets the modest large-effect threshold (≥ 2)
- Net result: Very low (★) [Low − 4 + 1 upgrade = Very low, floor]

PALF implication: β₂ is usable only as very-low-certainty evidence; the manuscript must declare this explicitly. The note on the PCS > 24 threshold (not derived from Theunissen) reinforces caution.

**4.3 Domain 3 — Metabolic / Obesity (β₃) — Low certainty (★★) [provisional]**

Status: the Shiri 2010 Am J Epidemiol PDF (obesity and LBP, cited as Ref 19) was not located in the directory. The rating is provisional, assuming methodological characteristics similar to Shiri 2010 Am J Med from the same group (AMSTAR rating Critically Low, but solid statistical methodology).

Assuming methodological equivalence:

- Starting point: Low
- Downgrade -1 for RoB (assumed pre-PROSPERO)
- No downgrade for inconsistency / indirectness / imprecision / publication bias (assumed)
- Upgrade +1 for plausible mechanism: IL-6 / TNF-α / leptin–adiponectin pathway from adipose tissue → low-grade systemic inflammation → nociceptive sensitisation. Triangulation with bariatric studies (reduction of inflammatory markers and chronic pain after weight loss).
- Net result: Low (★★) provisional. Verify PDF before finalising.

**4.4 Domain 4 — Preoperative opioids (β₄) — Moderate certainty (★★★)**

Justification:

- Starting point: Low (observational)
- No RoB downgrade: Lawal 2020 with Moderate AMSTAR, PROSPERO-registered, NOS by 2 reviewers, leave-one-out sensitivity analysis
- Downgrade -1 for inconsistency: high I² (>75%) reported in Lawal Table 2, attributable to variable definitions of "prolonged use"
- No indirectness downgrade (preoperative-use definition covers the lumbar context)
- No imprecision downgrade (k=14, N>1.9M, CI 2.94–9.64 narrow given the OR magnitude)
- No publication-bias downgrade (funnel + Egger conducted with k≥10)
- Upgrade +2 for very large effect: OR 5.32 exceeds the GRADE "very large effect" threshold (≥ 5) + clear iatrogenic mechanism (mu-receptor down-regulation + microglial TLR4 activation + opioid-induced hyperalgesia documented in animal RCTs and human observational studies)
- Net result: Moderate (★★★) [Low − 1 + 2 = ★★★]

PALF implication: β₄ is the most robust coefficient in the model. This is consistent with the empirical variance decomposition (91% iatrogenic): the strongest coefficient quantitatively is also the one with the highest GRADE certainty. This convergence supports the clinical priority of minimising preoperative opioids as a preventive strategy.

**4.5 Domain 5 — Preoperative BZD (β₅) — Moderate certainty (★★★)**

Justification:

- Starting point: Low
- No RoB downgrade: Lee 2025 (mislabelled as Ulrich) with Moderate AMSTAR, PROSPERO-registered, JBI 2024
- No inconsistency downgrade (k=6 with acceptable I²; the 6 studies converge)
- Downgrade -1 for indirectness: population restricted to US opioid-naïve patients; post-ACA search (2019+); generalisation to European / non-US contexts requires caution
- No imprecision downgrade (CI 1.53–2.05 narrow)
- No publication-bias downgrade (funnel + Egger conducted)
- Upgrade +1 for plausible mechanism: BZD–opioid cross-tolerance via the GABA–opioid system; potentiation of opioid-induced hyperalgesia; consistency with BZD OR 1.53 reported by Lawal 2020 (k=5) in a separate SR — convergence between two independent meta-analyses
- Net result: Moderate (★★★) [Low − 1 + 1 + triangulated support = ★★★]

**4.6 Domain 6 — Smoking (β₆) — Low certainty (★★)**

Justification:

- Starting point: Low
- Downgrade -1 for RoB: Shiri 2010 Am J Med AMSTAR Critically Low (partly offset by Dai 2021 Moderate, but the primary source is Shiri)
- Downgrade -1 for inconsistency: Shiri 2010 reports OR 1.79 for chronic LBP (k=8 in adults), Dai 2021 reports OR 1.23 for broad CMP (k=32). The outcome heterogeneity (LBP vs CMP) generates interpretive inconsistency, although both effect directions are consistent
- Downgrade -1 for indirectness: Dai 2021 measures CMP (includes fibromyalgia, neck, shoulder pain, etc.); application to specific LBP requires caution
- No imprecision downgrade (narrow CI in both)
- No publication-bias downgrade (Shiri trim-and-fill applied; Dai funnel + Egger)
- Upgrade +1 for plausible mechanism: disc microvascular ischaemia + nicotine-mediated oxidative stress + chronic low-grade inflammation
- Net result: Low (★★) [Low − 3 + 1 = ★★, GRADE convention floor of "Low" for observational evidence with a single upgrade]

**5. Global certainty synthesis of the PALF model**

| **GRADE certainty** | **Domain(s)** | **β_i weight in Z (worked example)** |
| --- | --- | --- |
| Moderate (★★★) | 4 (Opioid), 5 (BZD) | β₄ + β₅ = 2.243 (74% of Σβ_i positive) |
| Low (★★) | 1 (Sleep), 3 (Metabolic), 6 (Smoking) | β₁ + β₃ + β₆ = 1.269 (with β₃ provisional) |
| Very low (★) | 2 (Catastrophising) | β₂ = 0.742 |

Clinical reading:

- The domains with the largest magnitude (β₄ opioid and β₅ BZD) are also those with the highest GRADE certainty — favourable convergence between effect strength and evidence quality
- Catastrophising (β₂) is the weakest domain in certainty terms but retains operational relevance as a modifiable psychological predictor
- Sleep (β₁) and metabolic (β₃) ratings are contingent on verification of pending PDFs before a final rating

Implications for submission: the manuscript can defend the position that the PALF model is built on Moderate-certainty evidence for the two most influential coefficients (which dominate 91% of the iatrogenic variance) and Low–Very-Low-certainty evidence for the minor coefficients. This heterogeneous certainty architecture is typical of compound predictive models based on observational meta-analyses and must be honestly disclosed in the Limitations section.

**6. Limitations of this GRADE assessment**

- Single rater. As with AMSTAR 2 (Table S2 of the main manuscript), GRADE recommends two independent assessors. This evaluation is a methodological draft pending dual validation.
- Conservative policy. Liberal downgrading and conservative upgrading were applied. A less restrictive rater might produce 1–2 slightly higher ratings (especially Domain 4, which could be argued as High with an additional upgrade for documented dose-response).
- Unverified PDFs. Domain 1 (Runge 2024) and Domain 3 (Shiri 2010 Am J Epidemiol) require correct PDF retrieval before the final rating.
- GRADE for predictive models. GRADE was originally designed to assess intervention and prognosis evidence; its application to individual coefficients of a compound predictive model is a reasonable but not formally standardised extension. The assessment was performed domain by domain, not for the global model.
- Outcome heterogeneity. PALF integrates ORs from partially overlapping outcomes (generic CPSP, NPOU, chronic LBP, broad CMP). The indirectness downgrade attempts to capture this but does not resolve it structurally.

**7. Pending validation**

| **Step** | **Responsible** | **Status** |
| --- | --- | --- |
| Mathematical validation of the model (Var(Z), MC, error budget) | Sofía Perera Monje (Physics, Universität Tübingen) | In progress |
| AMSTAR 2 validation (second reviewer) | TBD | Pending |
| GRADE validation (second reviewer) | TBD (ideally clinical-methodologist) | Pending |
| Verification of PDFs for Refs 12 and 19 | Manuscript author | In progress |
| Re-labelling Ref 55 → Lee 2025 | Manuscript author | Immediate |

**8. Conclusion**

Summary of GRADE certainty for the six β_i sources of the PALF model:

- 2 domains with Moderate certainty (★★★): Opioid, BZD
- 3 domains with Low certainty (★★): Sleep, Metabolic, Smoking
- 1 domain with Very Low certainty (★): Catastrophising

The domains with the largest magnitude (β₄, β₅) coincide with those of highest certainty, which reinforces the robustness of the model in its quantitative core. The certainty heterogeneity across domains must be explicitly disclosed in Table 2 of the main manuscript and in the Limitations section.

This evaluation is explicitly pending validation by an independent second reviewer in accordance with GRADE guidance.

*S9 draft — v19.5.3 · 6 May 2026 · pending dual validation. For: Sofía Perera Monje (mathematical validation) and a second GRADE reviewer (TBD).*
