## Supplemental Data 2 for "Quantifying Biopsychosocial Risk Factor Domains for Chronic Pain Treatment Outcomes: An Umbrella Review with De Novo Meta-Analyses, Formal Uncertainty Propagation, and the Pain Amplifier Loop Framework (PALF)"

**Authors:** Javier Arranz-Durán, MD, FIPP, EDRA · Sofía Perera Monje, BSc

**PROSPERO:** CRD420261360881 (status: registered, pending CRD publication review) **OSF:** https://osf.io/5tqcz · DOI 10.17605/OSF.IO/5TQCZ **Manuscript version:** v19.5.3.3 · Compiled 5 May 2026

**Provenance and validation status:** Preliminary single-reviewer assessment (JAD); pending dual independent validation by SPM + external third reviewer (to be identified). A triple-validated version of this document will be deposited at OSF before peer-reviewed submission.

*Reference: Wolff RF, Moons KGM, Riley RD, et al. PROBAST: A Tool to Assess the Risk of Bias and Applicability of Prediction Model Studies. Ann Intern Med. 2019;170(1):51–58. doi:10.7326/M18-1376.*

**Note on terminology.** This supplement uses **PROBAST D1–D4** to refer to the four PROBAST tool domains (Participants, Predictors, Outcome, Analysis). The phrase **PALF Domain X** (1–6) refers to the six predictor domains of the PALF composite (Sleep, Catastrophizing, Metabolic, Opioid, BZD, Smoking). The two domain systems are conceptually orthogonal and should not be conflated.

**PROBAST was applied separately to the primary three-domain composite (sleep + catastrophizing + metabolic) and the secondary expanded six-domain composite (adding opioid, BZD, smoking).** The key distinction between the two composites for PROBAST purposes is **PROBAST D3 (Outcome)**: the three-domain composite is restricted to pain or pain-chronification outcomes; the expanded six-domain composite introduces pharmacological utilization outcomes that represent a different construct.

| **PROBAST Tool Domain** | **Signalling Questions** | **Judgment (3-domain / 6-domain)** | **Rationale** |
| --- | --- | --- | --- |
| **PROBAST D1 — Participants** | 1.1 Were appropriate data sources used? 1.2 Were inclusion/exclusion criteria appropriate? | Unclear / Unclear | Heterogeneous primary cohorts across source meta-analyses; no single inception cohort defines the PALF target population; source populations (postsurgical, community, MSK clinic) differ in baseline risk. |
| **PROBAST D2 — Predictors** | 2.1 Were predictors defined and assessed similarly in all included studies? 2.2 Were predictors available at the intended moment of use? 2.3 Was the outcome blinded at predictor assessment? | Low / Low | Standardized definitions per Supplementary S2 and Table 1; binary thresholds pre-specified; predictor assessment in source meta-analyses conducted blind to outcome by design (prospective cohorts and RCTs). |
| **PROBAST D3 — Outcome** | 3.1 Was the outcome determination appropriate? 3.2 Was the outcome defined and determined in a predefined, standard way? 3.3 Were predictors excluded from outcome assessment? 3.4 Was the same outcome definition used across all included studies? 3.5 Was the outcome determined without knowledge of predictor values? 3.6 Was the time interval between predictor assessment and outcome appropriate? | **Low (3-domain) / High (6-domain)** | Three-domain: all source meta-analyses target chronic pain or pain chronification — outcome homogeneity is maintained. Six-domain: opioid (PALF Domain 4) and BZD (PALF Domain 5) source meta-analyses target healthcare utilization outcomes (prolonged/persistent opioid use), not chronic pain — these are a different construct. Combining log-ORs across outcome families violates construct equivalence (see §2.4 Critical methodological note). |
| **PROBAST D4 — Analysis** | 4.1 Were there a reasonable number of participants with the outcome relative to the number of predictors? 4.2 Were continuous and categorical predictors handled appropriately? 4.3 Were all enrolled participants included in the analysis? 4.4 Were missing data handled appropriately? 4.5 Was the selection of predictors based on univariable screening avoided? 4.6 Were any complexities in the data (interactions, nonlinearity) accounted for? 4.7 Were relevant model performance measures reported? 4.8 Was overfitting or optimism in model performance accounted for? 4.9 Do the reported model performance measures correspond to the predicted values as reported? | **High / High** | Second-order pooling combines independently estimated meta-analytic log-ORs without individual patient data; small number of meta-analytic inputs per domain (k=8 for catastrophizing); no empirical validation cohort; no calibration assessment; Monte Carlo AUCs are tautological by construction (§2.8); no overfitting correction applicable to aggregate-data assembly; non-collapsibility of OR not fully correctable from aggregate data. |
| **Overall PROBAST judgment** | — | **Moderate (3-domain) / High (6-domain)** | Three-domain composite: PROBAST D3 is low risk (outcome homogeneity), but PROBAST D4 remains high risk (second-order pooling, no IPD validation). Overall moderate risk. Six-domain composite: PROBAST D3 is high risk (outcome heterogeneity) and PROBAST D4 is high risk — overall high risk of bias. |

*PROBAST reference: Wolff RF et al. Ann Intern Med 2019;170:51–58. doi:10.7326/M18-1376.*
