## Supplemental Data 3 for "Quantifying Biopsychosocial Risk Factor Domains for Chronic Pain Treatment Outcomes: An Umbrella Review with De Novo Meta-Analyses, Formal Uncertainty Propagation, and the Pain Amplifier Loop Framework (PALF)"

**Authors:** Javier Arranz-Durán, MD, FIPP, EDRA · Sofía Perera Monje, BSc

**PROSPERO:** CRD420261360881 (status: registered, pending CRD publication review) **OSF:** https://osf.io/5tqcz · DOI 10.17605/OSF.IO/5TQCZ **Manuscript version:** v19.5.3.3 · Compiled 5 May 2026

**Provenance and validation status:** Preliminary single-reviewer assessment (JAD); pending dual independent validation by SPM + external third reviewer (to be identified). A triple-validated version of this document will be deposited at OSF before peer-reviewed submission.

*Reference: Collins GS, Moons KGM, Dhiman P, et al. TRIPOD+AI statement: updated guidance for reporting clinical prediction models that use regression or machine learning methods. BMJ. 2024;385:e078378. doi:10.1136/bmj-2023-078378.*

**Rationale for N/A designations:** Items marked N/A with the notation “Composite score derived analytically from published meta-analytic estimates; no de novo training cohort” reflect the parameter-assembly nature of the PALF. The PALF assembles published meta-analytic log-ORs into a logistic equation using the delta method; it does not train a model on individual patient data. Accordingly, items requiring a training cohort, internal validation, model updating, or deployment infrastructure are not applicable at this development phase.

| **Item No.** | **TRIPOD+AI Item Description** | **Reported** | **Section/Page Reference** |
| --- | --- | --- | --- |
| 1 | Title — identify the study as developing or validating a prediction model | Yes | Title page |
| 2 | Abstract — provide a structured summary of objectives, methods, results, conclusions | Yes | Abstract |
| 3a | Background and rationale for developing or validating the prediction model | Yes | §1 Introduction |
| 3b | Objectives — specify the prediction objective, including target population and outcome | Yes | §1 (objectives paragraph) |
| 4a | Source of data — describe the data sources used | Yes | §2.2 Search Strategy; §2.4 Data Extraction |
| 4b | Eligibility criteria — describe inclusion/exclusion criteria for participants | Yes | §2.3 Inclusion and Exclusion Criteria |
| 5a | Outcome — define the outcome to be predicted | Yes | §2.4; Table 2 (Outcome column) |
| 5b | Outcome — describe how the outcome was assessed and whether blinding was applied | Yes | §2.4; Table S2 (AMSTAR assessment) |
| 6a | Predictors — define all predictors used in the model | Yes | §2.4; Table 2 (Binary Threshold column) |
| 6b | Predictors — describe how predictors were assessed and whether blinding was applied | Yes | §2.4; Table S1 (Covariate Adjustment Matrix) |
| 7a | Sample size — explain how the sample size was determined | Yes | §3.1 (source meta-analysis k and N reported) |
| 7b | Sample size — provide details of any sample size calculation | Yes | §2.8 (Monte Carlo N=50,000) |
| 8 | Missing data — describe how missing data were handled | N/A | Composite score derived analytically from published meta-analytic estimates; no de novo training cohort |
| 9 | Statistical analysis — describe all aspects of the statistical analysis | Yes | §2.5 Statistical Analysis; §2.6–§2.8 |
| 10 | Model development — describe how the final model was developed | N/A | Composite score derived analytically from published meta-analytic estimates; no de novo training cohort |
| 11 | Model performance — describe how model performance was assessed | Yes | §2.8 Mathematical Exploration; §3.10 |
| 12a | Risk groups — if applicable, provide details of how participants were classified into risk groups | Yes | Table 4 (Risk Tiers) |
| 12b | Risk group assessment — describe how risk groups were assessed | Yes | §3.9 Clinical Example; Table 4 note |
| 12c | Time horizon — specify the time horizon for the prediction | N/A | Composite score derived analytically from published meta-analytic estimates; no de novo training cohort |
| 12d | Validation strategy — describe any internal or external validation of the model | N/A | No internal validation cohort; parameter-assembly phase only |
| 13 | Participants flow — report the flow of participants through the study | N/A | Composite score derived analytically from published meta-analytic estimates; no de novo training cohort |
| 14 | Model specification — present the final prediction model | Yes | §2.6 (PALF equation); Table 1; Table 2; Table 3 |
| 15 | Model performance results — report performance measures with confidence intervals | N/A | No empirical performance data; Monte Carlo results are mathematical properties only (§3.10) |
| 16 | Model updating — report results of any model updating | N/A | Composite score derived analytically from published meta-analytic estimates; no de novo training cohort |
| 17 | Limitations — discuss limitations of the study and potential sources of bias | Yes | §4.4 Limitations (11 items); §3.11 PROBAST |
| 18 | Interpretation — provide an overall interpretation of the results | Yes | §4 Discussion; §5 Conclusions |
| 19 | Implications — discuss the potential clinical use of the model | N/A | Not deployable yet; PALF is presented as a framework for future prospective work (§5 Conclusions) |
| 20 | Supplementary information — provide additional information relevant to the study | Yes | Supplementary S1–S7; OSF repository (https://osf.io/5tqcz) |
| 21 | Funding — give the source of funding and the role of funders | Yes | Declarations (no external funding) |
| 22 | Data availability — state availability of additional data | Yes | Declarations; OSF https://osf.io/5tqcz |

*TRIPOD+AI reference: Collins GS et al. BMJ 2024;385:e078378. doi:10.1136/bmj-2023-078378*
