## Supplemental Data 4 for "Quantifying Biopsychosocial Risk Factor Domains for Chronic Pain Treatment Outcomes: An Umbrella Review with De Novo Meta-Analyses, Formal Uncertainty Propagation, and the Pain Amplifier Loop Framework (PALF)"

Companion to PALF manuscript v19.5.3

**Authors.** Javier Arranz Durán (JAD; guarantor), Sofia Perera Monje (SPM)

**PROSPERO registration.** CRD420261360881 — *registered, pending CRD publication review* (record submitted 5 May 2026)

**OSF repository.** https://osf.io/5tqcz — DOI [10.17605/OSF.IO/5TQCZ](https://doi.org/10.17605/OSF.IO/5TQCZ)

**Compilation date.** 5 May 2026 — **Document version.** v19.5.3

### Scope and conventions (consistent with PROSPERO CRD420261360881)

- **Information sources searched.** PubMed/MEDLINE, Scopus, and Cochrane Database of Systematic Reviews (CDSR / Cochrane Library).
- **Date coverage.** Inception to **March 2026** (no lower-date restriction; upper search date executed in March 2026, as registered in PROSPERO).
- **Language.** **English only**, applied at the database level (English[lang] in PubMed; LANGUAGE("English") in Scopus; English limit in Cochrane Library).
- **Population filter.** Adults (≥18 y); humans only. PubMed humans/adult filters applied; Scopus and CDSR equivalent age/human filters applied where supported.
- **Publication types of interest.** Systematic reviews and meta-analyses (umbrella review parent search), plus primary cohort/case-control studies for de novo meta-analyses (metabolic and BZD subanalysis, per PROSPERO study-design field).
- **Other methods.** Backward citation searching (reference-list checking) of every adopted source meta-analysis (per PROSPERO “Other methods of identifying studies”).
- **Provenance.** Search strings below were executed by JAD (guarantor) in March 2026 as a single-reviewer preliminary pass. Independent re-execution by SPM and a third external reviewer (TBD) is **pending** before the triple-validated version (see *Search re-execution and dual-validation plan* at end).
- **Hit counts.** All record counts in this document are reported as **preliminary placeholders (N=__)** — final counts will be reported in the triple-validated version. **No fabricated counts are reported here.**

**Note on outcome-to-domain mapping.** Each domain’s exposure–outcome contrast below is identical to the corresponding row in the PROSPERO “Main outcomes” field (CRD420261360881).

### Domain 1 — Sleep disturbance → Chronic musculoskeletal pain

- **Exposure.** Sleep disturbance — operationalised as ≥3-month sleep impairment by validated instruments: PSQI (Pittsburgh Sleep Quality Index >5), ISI (Insomnia Severity Index), or ESS (Epworth Sleepiness Scale).
- **Comparator.** Unexposed reference (e.g., PSQI ≤5).
- **Primary outcome.** Chronic musculoskeletal pain (≥3-month follow-up).

#### 1a. PubMed/MEDLINE

```
((("Sleep Initiation and Maintenance Disorders"[MeSH] OR "Sleep Wake Disorders"[MeSH]
   OR "sleep disorders"[tiab] OR "sleep disturbance"[tiab] OR "insomnia"[tiab]
   OR "PSQI"[tiab] OR "Pittsburgh Sleep Quality Index"[tiab]
   OR "Insomnia Severity Index"[tiab] OR "ISI"[tiab]
   OR "Epworth Sleepiness Scale"[tiab] OR "ESS"[tiab])
  AND ("Chronic Pain"[MeSH] OR "Musculoskeletal Pain"[MeSH]
       OR "chronic pain"[tiab] OR "musculoskeletal pain"[tiab]
       OR "pain chronification"[tiab] OR "chronification"[tiab])
  AND ("Meta-Analysis"[Publication Type] OR "Systematic Review"[Publication Type]
       OR "meta-analysis"[tiab] OR "meta analysis"[tiab] OR "systematic review"[tiab]))
 AND English[lang]
 AND (humans[MeSH] AND adult[MeSH]))
```

*Filters applied:* Humans; Adult (19+); English. *Date executed:* March 2026.

#### 1b. Scopus (TITLE-ABS-KEY)

```
( TITLE-ABS-KEY ( "sleep disorder*" OR "sleep disturbance*" OR "insomnia"
   OR "PSQI" OR "Pittsburgh Sleep Quality Index"
   OR "Insomnia Severity Index" OR "ISI"
   OR "Epworth Sleepiness Scale" OR "ESS" )
 AND TITLE-ABS-KEY ( "chronic pain" OR "musculoskeletal pain"
   OR "chronification" OR "pain chronification" )
 AND TITLE-ABS-KEY ( "meta-analysis" OR "meta analysis" OR "systematic review" ) )
 AND ( LIMIT-TO ( LANGUAGE , "English" ) )
 AND ( LIMIT-TO ( DOCTYPE , "re" ) OR LIMIT-TO ( DOCTYPE , "ar" ) )
```

*Date executed:* March 2026.

#### 1c. Cochrane Library (CDSR)

```
#1  MeSH descriptor: [Sleep Wake Disorders] explode all trees
#2  (sleep NEAR/3 (disorder* OR disturb* OR quality)):ti,ab,kw
#3  (insomnia OR PSQI OR "Pittsburgh Sleep Quality Index"
     OR "Insomnia Severity Index" OR ISI OR "Epworth Sleepiness Scale"):ti,ab,kw
#4  #1 OR #2 OR #3
#5  MeSH descriptor: [Chronic Pain] explode all trees
#6  ("chronic pain" OR "musculoskeletal pain" OR chronification):ti,ab,kw
#7  #5 OR #6
#8  #4 AND #7        — restrict to Cochrane Reviews (CDSR)
```

*Date executed:* March 2026. Limits: English; Cochrane Reviews.

#### 1d. Records identified (preliminary)

Records identified — **preliminary single-reviewer count, pending dual-reviewer verification**: PubMed N=__; Scopus N=__; Cochrane (CDSR) N=__; total before deduplication N=__; after deduplication N=__. Final counts will be reported in the triple-validated version.

### Domain 2 — Pain catastrophizing → Chronic postsurgical pain

- **Exposure.** Preoperative Pain Catastrophizing Scale (PCS) ≥75th percentile (operationalised in source meta-analyses as PCS ≥30, or equivalent threshold).
- **Comparator.** PCS ≤24.
- **Primary outcome.** Chronic postsurgical pain (≥3-month postoperative).

#### 2a. PubMed/MEDLINE

```
((("Catastrophization"[MeSH] OR "catastrophizing"[tiab] OR "catastrophising"[tiab]
   OR "pain catastrophizing"[tiab] OR "PCS"[tiab]
   OR "Pain Catastrophizing Scale"[tiab])
  AND ("Pain, Postoperative"[MeSH] OR "Chronic Pain"[MeSH]
       OR "chronic postsurgical pain"[tiab] OR "chronic post-surgical pain"[tiab]
       OR "persistent postsurgical pain"[tiab] OR "CPSP"[tiab]
       OR "postoperative pain"[tiab])
  AND ("Meta-Analysis"[Publication Type] OR "Systematic Review"[Publication Type]
       OR "meta-analysis"[tiab] OR "meta analysis"[tiab] OR "systematic review"[tiab]))
 AND English[lang]
 AND (humans[MeSH] AND adult[MeSH]))
```

*Filters:* Humans; Adult; English. *Date executed:* March 2026.

#### 2b. Scopus (TITLE-ABS-KEY)

```
( TITLE-ABS-KEY ( "catastroph*" OR "pain catastrophizing"
   OR "Pain Catastrophizing Scale" OR "PCS" )
 AND TITLE-ABS-KEY ( "chronic postsurgical pain" OR "chronic post-surgical pain"
   OR "persistent postsurgical pain" OR "CPSP" OR "postoperative pain" )
 AND TITLE-ABS-KEY ( "meta-analysis" OR "meta analysis" OR "systematic review" ) )
 AND ( LIMIT-TO ( LANGUAGE , "English" ) )
 AND ( LIMIT-TO ( DOCTYPE , "re" ) OR LIMIT-TO ( DOCTYPE , "ar" ) )
```

*Date executed:* March 2026.

#### 2c. Cochrane Library (CDSR)

```
#1  MeSH descriptor: [Catastrophization] explode all trees
#2  (catastroph* OR "pain catastrophizing" OR "Pain Catastrophizing Scale" OR PCS):ti,ab,kw
#3  #1 OR #2
#4  MeSH descriptor: [Pain, Postoperative] explode all trees
#5  ("chronic postsurgical pain" OR "persistent postsurgical pain"
     OR CPSP OR "postoperative pain"):ti,ab,kw
#6  #4 OR #5
#7  #3 AND #6        — restrict to Cochrane Reviews (CDSR)
```

*Date executed:* March 2026. Limits: English; Cochrane Reviews.

#### 2d. Records identified (preliminary)

Records identified — **preliminary single-reviewer count, pending dual-reviewer verification**: PubMed N=__; Scopus N=__; Cochrane (CDSR) N=__; total before deduplication N=__; after deduplication N=__. Final counts will be reported in the triple-validated version.

### Domain 3 — Metabolic / obesity exposure → Chronic low back pain

- **Exposure.** BMI ≥30 kg/m² **or** metabolic syndrome (per source criteria).
- **Comparator.** BMI <30; absence of metabolic syndrome.
- **Primary outcome.** Chronic low back pain (≥3-month).

#### 3a. PubMed/MEDLINE

```
((("Obesity"[MeSH] OR "Body Mass Index"[MeSH] OR "Metabolic Syndrome"[MeSH]
   OR "obesity"[tiab] OR "obese"[tiab] OR "BMI"[tiab]
   OR "body mass index"[tiab] OR "metabolic syndrome"[tiab]
   OR "metabolic-inflammatory"[tiab])
  AND ("Low Back Pain"[MeSH] OR "Chronic Pain"[MeSH]
       OR "low back pain"[tiab] OR "lumbar pain"[tiab]
       OR "chronic low back pain"[tiab] OR "CLBP"[tiab])
  AND ("Meta-Analysis"[Publication Type] OR "Systematic Review"[Publication Type]
       OR "meta-analysis"[tiab] OR "meta analysis"[tiab] OR "systematic review"[tiab]
       OR "Cohort Studies"[MeSH] OR "Case-Control Studies"[MeSH]))
 AND English[lang]
 AND (humans[MeSH] AND adult[MeSH]))
```

*Filters:* Humans; Adult; English. Cohort/case-control study types retained because Domain 3 includes a **de novo meta-analysis** per PROSPERO. *Date executed:* March 2026.

#### 3b. Scopus (TITLE-ABS-KEY)

```
( TITLE-ABS-KEY ( "obesity" OR "obese" OR "BMI" OR "body mass index"
   OR "metabolic syndrome" OR "metabolic-inflammatory" )
 AND TITLE-ABS-KEY ( "low back pain" OR "lumbar pain" OR "chronic low back pain" OR "CLBP" )
 AND TITLE-ABS-KEY ( "meta-analysis" OR "systematic review"
   OR "cohort study" OR "case-control" OR "prospective cohort" ) )
 AND ( LIMIT-TO ( LANGUAGE , "English" ) )
 AND ( LIMIT-TO ( DOCTYPE , "re" ) OR LIMIT-TO ( DOCTYPE , "ar" ) )
```

*Date executed:* March 2026.

#### 3c. Cochrane Library (CDSR)

```
#1  MeSH descriptor: [Obesity] explode all trees
#2  MeSH descriptor: [Body Mass Index] this term only
#3  MeSH descriptor: [Metabolic Syndrome] explode all trees
#4  (obesity OR obese OR BMI OR "body mass index" OR "metabolic syndrome"):ti,ab,kw
#5  #1 OR #2 OR #3 OR #4
#6  MeSH descriptor: [Low Back Pain] explode all trees
#7  ("low back pain" OR "lumbar pain" OR CLBP OR "chronic low back pain"):ti,ab,kw
#8  #6 OR #7
#9  #5 AND #8        — restrict to Cochrane Reviews (CDSR)
```

*Date executed:* March 2026. Limits: English; Cochrane Reviews.

#### 3d. Records identified (preliminary)

Records identified — **preliminary single-reviewer count, pending dual-reviewer verification**: PubMed N=__; Scopus N=__; Cochrane (CDSR) N=__; total before deduplication N=__; after deduplication N=__. Final counts will be reported in the triple-validated version.

### Domain 4 — Preoperative opioid exposure → Prolonged postoperative opioid use

- **Exposure.** Chronic opioid use ≥3 months pre-surgery.
- **Comparator.** No preoperative opioid prescription.
- **Primary outcome.** Prolonged postoperative opioid use.

#### 4a. PubMed/MEDLINE

```
((("Analgesics, Opioid"[MeSH] OR "Opioid-Related Disorders"[MeSH]
   OR "opioid"[tiab] OR "opioids"[tiab]
   OR "preoperative opioid"[tiab] OR "chronic opioid"[tiab]
   OR "opioid exposure"[tiab] OR "opioid use"[tiab])
  AND ("postoperative opioid"[tiab] OR "prolonged opioid"[tiab]
       OR "persistent opioid use"[tiab] OR "chronic postoperative opioid"[tiab]
       OR "new persistent opioid use"[tiab] OR "NPOU"[tiab])
  AND ("Meta-Analysis"[Publication Type] OR "Systematic Review"[Publication Type]
       OR "meta-analysis"[tiab] OR "meta analysis"[tiab] OR "systematic review"[tiab]))
 AND English[lang]
 AND (humans[MeSH] AND adult[MeSH]))
```

*Filters:* Humans; Adult; English. *Date executed:* March 2026.

#### 4b. Scopus (TITLE-ABS-KEY)

```
( TITLE-ABS-KEY ( "preoperative opioid" OR "chronic opioid"
   OR "opioid exposure" OR "opioid use" )
 AND TITLE-ABS-KEY ( "prolonged postoperative opioid"
   OR "persistent opioid use" OR "new persistent opioid"
   OR "chronic postoperative opioid" OR "NPOU" )
 AND TITLE-ABS-KEY ( "meta-analysis" OR "systematic review" ) )
 AND ( LIMIT-TO ( LANGUAGE , "English" ) )
 AND ( LIMIT-TO ( DOCTYPE , "re" ) OR LIMIT-TO ( DOCTYPE , "ar" ) )
```

*Date executed:* March 2026.

#### 4c. Cochrane Library (CDSR)

```
#1  MeSH descriptor: [Analgesics, Opioid] explode all trees
#2  MeSH descriptor: [Opioid-Related Disorders] explode all trees
#3  ("preoperative opioid" OR "chronic opioid" OR "opioid exposure"):ti,ab,kw
#4  #1 OR #2 OR #3
#5  ("persistent opioid use" OR "prolonged postoperative opioid"
     OR "new persistent opioid" OR NPOU):ti,ab,kw
#6  #4 AND #5        — restrict to Cochrane Reviews (CDSR)
```

*Date executed:* March 2026. Limits: English; Cochrane Reviews.

#### 4d. Records identified (preliminary)

Records identified — **preliminary single-reviewer count, pending dual-reviewer verification**: PubMed N=__; Scopus N=__; Cochrane (CDSR) N=__; total before deduplication N=__; after deduplication N=__. Final counts will be reported in the triple-validated version.

### Domain 5 — Benzodiazepine co-prescription → Persistent postoperative opioid use

- **Exposure.** Concurrent benzodiazepine (BZD) prescription during opioid therapy.
- **Comparator.** No active BZD prescription.
- **Primary outcome.** Persistent postoperative opioid use.

#### 5a. PubMed/MEDLINE

```
((("Benzodiazepines"[MeSH] OR "Hypnotics and Sedatives"[MeSH]
   OR "benzodiazepine"[tiab] OR "benzodiazepines"[tiab] OR "BZD"[tiab]
   OR "diazepam"[tiab] OR "lorazepam"[tiab] OR "alprazolam"[tiab]
   OR "clonazepam"[tiab])
  AND ("Analgesics, Opioid"[MeSH]
       OR "opioid"[tiab] OR "opioids"[tiab]
       OR "co-prescription"[tiab] OR "concurrent use"[tiab]
       OR "persistent opioid use"[tiab] OR "prolonged opioid"[tiab])
  AND ("Meta-Analysis"[Publication Type] OR "Systematic Review"[Publication Type]
       OR "meta-analysis"[tiab] OR "meta analysis"[tiab] OR "systematic review"[tiab]
       OR "Cohort Studies"[MeSH] OR "Case-Control Studies"[MeSH]))
 AND English[lang]
 AND (humans[MeSH] AND adult[MeSH]))
```

*Filters:* Humans; Adult; English. Cohort/case-control retained because Domain 5 includes a **de novo BZD subanalysis** per PROSPERO. *Date executed:* March 2026.

#### 5b. Scopus (TITLE-ABS-KEY)

```
( TITLE-ABS-KEY ( "benzodiazepine*" OR "BZD" OR "hypnotic*" OR "sedative*"
   OR "diazepam" OR "lorazepam" OR "alprazolam" OR "clonazepam" )
 AND TITLE-ABS-KEY ( "opioid*" OR "co-prescription" OR "concurrent"
   OR "persistent opioid use" OR "prolonged opioid" )
 AND TITLE-ABS-KEY ( "meta-analysis" OR "systematic review"
   OR "cohort study" OR "case-control" ) )
 AND ( LIMIT-TO ( LANGUAGE , "English" ) )
 AND ( LIMIT-TO ( DOCTYPE , "re" ) OR LIMIT-TO ( DOCTYPE , "ar" ) )
```

*Date executed:* March 2026.

#### 5c. Cochrane Library (CDSR)

```
#1  MeSH descriptor: [Benzodiazepines] explode all trees
#2  MeSH descriptor: [Hypnotics and Sedatives] explode all trees
#3  (benzodiazepine* OR BZD OR diazepam OR lorazepam OR alprazolam OR clonazepam):ti,ab,kw
#4  #1 OR #2 OR #3
#5  MeSH descriptor: [Analgesics, Opioid] explode all trees
#6  (opioid* OR "co-prescription" OR "concurrent use"
     OR "persistent opioid use"):ti,ab,kw
#7  #5 OR #6
#8  #4 AND #7        — restrict to Cochrane Reviews (CDSR)
```

*Date executed:* March 2026. Limits: English; Cochrane Reviews.

#### 5d. Records identified (preliminary)

Records identified — **preliminary single-reviewer count, pending dual-reviewer verification**: PubMed N=__; Scopus N=__; Cochrane (CDSR) N=__; total before deduplication N=__; after deduplication N=__. Final counts will be reported in the triple-validated version.

### Domain 6 — Smoking → Chronic low back pain

- **Exposure.** Current daily smoking.
- **Comparator.** Never smoker / former smoker.
- **Primary outcome.** Chronic low back pain.

#### 6a. PubMed/MEDLINE

```
((("Smoking"[MeSH] OR "Tobacco Smoking"[MeSH] OR "Cigarette Smoking"[MeSH]
   OR "Tobacco Use"[MeSH]
   OR "smoking"[tiab] OR "smoker"[tiab] OR "smokers"[tiab]
   OR "tobacco"[tiab] OR "cigarette"[tiab] OR "nicotine"[tiab])
  AND ("Low Back Pain"[MeSH] OR "Chronic Pain"[MeSH]
       OR "low back pain"[tiab] OR "lumbar pain"[tiab]
       OR "chronic low back pain"[tiab] OR "CLBP"[tiab]
       OR "musculoskeletal pain"[tiab])
  AND ("Meta-Analysis"[Publication Type] OR "Systematic Review"[Publication Type]
       OR "meta-analysis"[tiab] OR "meta analysis"[tiab] OR "systematic review"[tiab]))
 AND English[lang]
 AND (humans[MeSH] AND adult[MeSH]))
```

*Filters:* Humans; Adult; English. *Date executed:* March 2026.

#### 6b. Scopus (TITLE-ABS-KEY)

```
( TITLE-ABS-KEY ( "smoking" OR "smoker*" OR "tobacco" OR "cigarette*" OR "nicotine" )
 AND TITLE-ABS-KEY ( "low back pain" OR "lumbar pain"
   OR "chronic low back pain" OR "CLBP" OR "musculoskeletal pain" )
 AND TITLE-ABS-KEY ( "meta-analysis" OR "systematic review" ) )
 AND ( LIMIT-TO ( LANGUAGE , "English" ) )
 AND ( LIMIT-TO ( DOCTYPE , "re" ) OR LIMIT-TO ( DOCTYPE , "ar" ) )
```

*Date executed:* March 2026.

#### 6c. Cochrane Library (CDSR)

```
#1  MeSH descriptor: [Smoking] explode all trees
#2  MeSH descriptor: [Tobacco Use] explode all trees
#3  (smoking OR smoker* OR tobacco OR cigarette* OR nicotine):ti,ab,kw
#4  #1 OR #2 OR #3
#5  MeSH descriptor: [Low Back Pain] explode all trees
#6  ("low back pain" OR "lumbar pain" OR "chronic low back pain"
     OR CLBP OR "musculoskeletal pain"):ti,ab,kw
#7  #5 OR #6
#8  #4 AND #7        — restrict to Cochrane Reviews (CDSR)
```

*Date executed:* March 2026. Limits: English; Cochrane Reviews.

#### 6d. Records identified (preliminary)

Records identified — **preliminary single-reviewer count, pending dual-reviewer verification**: PubMed N=__; Scopus N=__; Cochrane (CDSR) N=__; total before deduplication N=__; after deduplication N=__. Final counts will be reported in the triple-validated version.

### Other sources of identification — backward citation searching

Per PROSPERO CRD420261360881 (“Other methods of identifying studies: reference list checking — backward citation searching”), the reference list of every adopted source meta-analysis was screened by JAD against the same eligibility criteria. Specifically, backward citation searching was applied to:

1. The source meta-analysis adopted for **Domain 1 (Sleep → chronic musculoskeletal pain)**.
2. The source meta-analysis adopted for **Domain 2 (Catastrophizing → chronic postsurgical pain)**.
3. The source meta-analyses contributing to the **Domain 3 (Metabolic → CLBP)** *de novo* meta-analysis.
4. The source meta-analysis adopted for **Domain 4 (Preoperative opioid → prolonged postoperative opioid use)**.
5. The source meta-analyses contributing to the **Domain 5 (BZD co-prescription → persistent opioid use)** *de novo* subanalysis.
6. The source meta-analysis adopted for **Domain 6 (Smoking → CLBP)**.

Records identified through backward citation searching — **preliminary single-reviewer count, pending dual-reviewer verification**: total additional records N=__; eligible after screening N=__. Final counts will be reported in the triple-validated version. Forward citation searching was not pre-specified in the PROSPERO record and is therefore not applied.

### Search re-execution and dual-validation plan

In accordance with the PROSPERO record (selection process: “independently by ≥2 people or person/machine combination”) and with the v19.5.3 manuscript (§2.2 Search Strategy), all six domain-specific search strings deposited in this Supplement S5 will be **re-executed independently** as part of the dual-validation procedure that produces the triple-validated version of the PALF manuscript and supplements:

1. **Step 1 — Independent re-execution by SPM.** Sofia Perera Monje will re-execute the unmodified strings (Domains 1–6, three databases each) in PubMed/MEDLINE, Scopus and Cochrane Library, recording exact hit counts, search dates, and any database interface differences.
2. **Step 2 — Independent re-execution by 3rd external reviewer (TBD).** A third reviewer external to the authorship team will re-execute the same strings independently, blind to SPM’s counts, and record their own counts.
3. **Step 3 — Discrepancy reconciliation.** Hit-count discrepancies >5% between any two independent runs will be investigated jointly (database-side updates, filter behaviour, language tagging, indexing lag). Strings will be amended only if a defect is identified; any amendment will be tracked in a versioned change log appended to this supplement.
4. **Step 4 — Dual screening of the merged record set.** Title/abstract and full-text screening will then be performed independently and in duplicate (SPM + 3rd reviewer); JAD acts as guarantor and tie-breaker only, **not** as a primary screener. Inter-rater agreement (Cohen’s κ) will be reported in the triple-validated PRISMA flow diagram.
5. **Step 5 — Triple-validated counts.** Final, reviewer-agreed PubMed / Scopus / CDSR / backward-citation counts will replace the N=__ placeholders throughout this supplement and in Figure 2 (PRISMA flow) of the manuscript.
6. **Step 6 — Public deposition.** The triple-validated S5 (with definitive counts, search-execution dates, and reviewer initials) will be deposited at osf.io/5tqcz and will accompany the peer-reviewed publication.

Until Steps 1–6 are completed, all quantitative outputs of this supplement — and any downstream PRISMA flow numbers in the manuscript — are flagged as **PRELIMINARY**.

*Document version v19.5.3 | Compiled 5 May 2026 | OSF: https://osf.io/5tqcz | PROSPERO: CRD420261360881 (registered, pending publication review)*
