## Supplemental Data 5 for "Quantifying Biopsychosocial Risk Factor Domains for Chronic Pain Treatment Outcomes: An Umbrella Review with De Novo Meta-Analyses, Formal Uncertainty Propagation, and the Pain Amplifier Loop Framework (PALF)"

Companion to PALF manuscript v19.5.3

**Authors.** Javier Arranz Durán (JAD; guarantor), Sofia Perera Monje (SPM)

**PROSPERO registration.** CRD420261360881 — *registered, pending CRD publication review* (record submitted 5 May 2026)

**OSF repository.** https://osf.io/5tqcz — DOI [10.17605/OSF.IO/5TQCZ](https://doi.org/10.17605/OSF.IO/5TQCZ)

**Compilation date.** 5 May 2026 — **Document version.** v19.5.3

**Reporting guideline reference.** Gates M, Gates A, Pieper D, Fernandes RM, Tricco AC, Moher D, Brennan SE, Li T, Pollock M, Lunny C, Sepúlveda D, McKenzie JE, Scott SD, Robinson KA, Matthias K, Bougioukas KI, Fusar-Poli P, Whiting P, Moss SJ, Hartling L. **Reporting guideline for overviews of reviews of healthcare interventions: development of the PRIOR statement.** *BMJ* 2022;378:e070849. doi:[10.1136/bmj-2022-070849](https://doi.org/10.1136/bmj-2022-070849)

**Provenance and validation status (read first).** All checklist responses below were prepared by JAD on the basis of the v19.5.3 manuscript, the PROSPERO record CRD420261360881 (5 May 2026), and the OSF protocol (osf.io/5tqcz). Per the PROSPERO record, JAD acts as guarantor only and is **not** a primary GRADE/risk-of-bias rater. Dual independent validation by SPM and a third external reviewer (TBD) is pending; entries marked *“Yes (preliminary)”* will be re-confirmed in the triple-validated version of this checklist that will accompany the peer-reviewed manuscript.

### How to read this checklist

- **Item No.** — Item number from the PRIOR (PRISMA-O) statement (Gates et al., BMJ 2022).
- **Section / topic** — PRIOR section heading.
- **Checklist item (abridged)** — Reporting requirement.
- **Response** — *Yes / Partial / No / N/A*. Responses currently labelled *Yes (preliminary)* are pending dual-reviewer validation.
- **Location in v19.5.3** — Manuscript section, table, figure, or supplement where the item is reported.

### PRIOR / PRISMA-O 27-item checklist

| # | Section / topic | Checklist item (abridged) | Response | Location in PALF v19.5.3 |
| --- | --- | --- | --- | --- |
| **1** | Title | Identify the report as an overview of reviews. | Yes | Title page; Abstract header (“umbrella review with de novo meta-analyses”) |
| **2** | Abstract | Structured summary covering background, objectives, eligibility, sources, synthesis, results and conclusions. | Yes | Abstract (structured, ~350 words) |
| **3** | Rationale | Describe the rationale for the overview in the context of existing knowledge. | Yes | §1 Introduction (¶1–¶3) |
| **4** | Objectives | State explicit objectives, including PICO/PECO components addressed. | Yes | §1 Introduction (final paragraph “Aims”); mirrors PROSPERO “Review objectives” |
| **5** | Eligibility criteria | Specify inclusion/exclusion criteria for reviews and for primary studies within reviews. | Yes | §2.3 Inclusion and Exclusion Criteria; PROSPERO Eligibility section |
| **6** | Information sources | List all databases, registers, websites, organisations, reference lists and other sources searched, with dates of last search. | Yes | §2.2 Search Strategy; **Supplement S5 (Full Search Strategies)**; PROSPERO “Main sources” (PubMed/MEDLINE, Scopus, CDSR; inception → March 2026) |
| **7** | Search strategy | Present the full search strategies for all databases, registers and websites, including any filters and limits used. | Yes | **Supplement S5** (full reproducible Boolean strings per domain × database); §2.2 |
| **8** | Selection process | Specify the methods used to decide whether a review met the inclusion criteria, including whether independently and in duplicate. | Yes (preliminary) | §2.2 / §3.1; PROSPERO “Selection process” (≥2 reviewers or person/machine combination). *Current quantitative outputs reflect single-reviewer (JAD) screening; SPM + 3rd reviewer dual validation pending — see footnote.* |
| **9** | Data collection process | Specify the methods used to collect data from reviews and any included primary studies, including independence/duplication and contact with authors. | Yes (preliminary) | §2.4 Data Extraction; PROSPERO “Data collection process” (≥2 reviewers; authors not contacted). Dual extraction pending. |
| **10a** | Data items — outcomes | List and define all outcomes for which data were sought. | Yes | §2.4; Table 1 (Domain × exposure × outcome × source MA); PROSPERO “Main outcomes” |
| **10b** | Data items — other variables | List and define all other variables for which data were sought (e.g., participants, settings, designs of primary studies, funding). | Yes | §2.4; Table 1; Table S1 (extraction dictionary) |
| **11** | Study (review) risk of bias assessment | Specify the methods used to assess risk of bias in the included reviews, including tool, independence and number of assessors. | Yes (preliminary) | §2.5 Statistical Analysis & Risk of Bias; **AMSTAR-2** (reviews) + **PROBAST** (composite prediction model) + **ROBINS-I** (constituent primary studies); PROSPERO “Study risk of bias”. Single-reviewer ratings; dual + 3rd reviewer adjudication pending. |
| **12** | Effect measures | Specify for each outcome the effect measure(s) used in the synthesis or presentation of results. | Yes | §2.5 (pooled adjusted OR; β = ln(OR); composite probability P = 1/(1+exp(−Z)) with delta-method 95% CI) |
| **13** | Synthesis methods — eligibility for synthesis | Describe the processes used to decide which reviews/studies were eligible for each synthesis. | Yes | §2.4–§2.5; §3.1 |
| **14a** | Synthesis methods — preparing data | Describe any methods required to prepare data for presentation or synthesis (e.g., handling overlap, recalculations). | Yes | §2.4 (recomputation rules); §2.5; §3.1 (overlap handling) |
| **14b** | **Quality of included reviews — umbrella-specific** | Describe how the methodological quality of the included systematic reviews was assessed and incorporated into the synthesis. | Yes (preliminary) | §2.5 (AMSTAR-2 protocol); **Table S2 — AMSTAR-2 ratings** (Supplement); PROBAST applied to the composite model |
| **15** | Synthesis methods — tabulation/visual | Describe any methods used to tabulate or visually display results of individual reviews and syntheses. | Yes | §3.2–§3.7 (per-domain narrative + forest plots); Figure 3 (PALF composite); Table 2 (pooled OR per domain) |
| **16** | Synthesis methods — quantitative | Describe any methods used to synthesise results quantitatively, including model and software. | Yes | §2.5 (DerSimonian–Laird primary; REML/Knapp–Hartung sensitivity; PET-PEESE; software: R metafor); §2.6 composite construction; §2.7 delta method |
| **17a** | Synthesis methods — heterogeneity | Describe any methods used to explore heterogeneity across reviews. | Yes | §2.5 (I², Cochran’s Q, τ²); §3.2–§3.7 per domain |
| **17b** | **Overlap assessment — umbrella-specific** | Describe how overlap of primary studies among included reviews was assessed and managed. | Yes | **§3.1 — Corrected Covered Area (CCA) = 3.2% (slight overlap, Pieper threshold)**; §2.4 overlap handling rules |
| **18** | Reporting bias | Describe any methods used to assess risk of bias due to missing results in a synthesis. | Yes | §2.5 (PET-PEESE; funnel asymmetry); §3.2–§3.7 per domain; PROSPERO “Reporting bias assessment” |
| **19** | Certainty assessment (per outcome) | Describe any methods used to assess certainty (or confidence) in the body of evidence for an outcome. | Yes (preliminary) | §2.5 GRADE protocol; PROSPERO “Certainty assessment” (5 GRADE dimensions; SPM + 3rd reviewer; JAD non-rater) |
| **20** | Study selection results | Numbers screened/included; PRISMA flow. | Yes (preliminary) | **Figure 2 — PRISMA 2020 flow diagram**; §3.1. *Counts are preliminary single-reviewer; final triple-validated counts to follow.* |
| **21** | Review characteristics | Cite each included review and present its characteristics. | Yes | Table 1 (source meta-analyses, k, N, OR, year, AMSTAR-2); §3.2–§3.7 |
| **22a** | Risk of bias in included reviews — results | Present assessments of risk of bias for each included review. | Yes (preliminary) | **Table S2 (AMSTAR-2)**; §3.11 (PROBAST for composite); ROBINS-I summary in §3.2–§3.7 where applicable |
| **22b** | **Certainty across reviews — umbrella-specific** | For each outcome assessed, present the assessments of certainty across the body of reviews. | Yes (preliminary) | **Supplement S9 — GRADE summary of findings** (per-domain Very Low / Low / Moderate / High); §3.2–§3.7 |
| **23** | Results of syntheses | For each synthesis, present results including summary effect, CI, heterogeneity, and number of reviews/studies. | Yes | §3.2–§3.7 (per-domain pooled OR, 95% CI, I², k, N); §3.8 (PALF composite + delta-method 95% CI); Table 2 |
| **24** | Discussion | Provide a general interpretation of the results in the context of other evidence. | Yes | §4.1–§4.3 Discussion |
| **25** | Limitations | Discuss limitations of the evidence and of the review process. | Yes | §4.4–§4.5; explicit declaration of single-reviewer preliminary status pending dual validation |
| **26** | Conclusions | Provide a general interpretation of the results and implications for practice/policy/research. | Yes | §5 Conclusions |
| **27** | Other information — registration, protocol, funding, conflicts, data availability | Provide registration details, protocol availability, funding sources, competing interests, and availability of data, code and other materials. | Yes | Title page footnotes (PROSPERO CRD420261360881; OSF [10.17605/OSF.IO/5TQCZ](https://doi.org/10.17605/OSF.IO/5TQCZ)); §6 Funding; §7 Competing interests; §8 Data & code availability (medRxiv preprint DOI [10.64898/2026.03.22.26348998](https://doi.org/10.64898/2026.03.22.26348998)) |

#### Footnote — Provisional responses pending dual-reviewer validation

Items **8, 9, 11, 14b, 19, 20, 22a, 22b** are marked *“Yes (preliminary)”* because the underlying screening, data extraction, risk-of-bias rating, and GRADE certainty assessments in v19.5.3 reflect a single-reviewer pass conducted by JAD (guarantor). In accordance with the PROSPERO record (CRD420261360881, 5 May 2026), independent dual validation by SPM and a third external reviewer (to be identified) is pending. JAD will not act as a primary GRADE or risk-of-bias rater in the verified version. The triple-validated version of this checklist — with final reviewer-agreed responses, κ statistics where applicable, and definitive PRISMA flow numbers — will accompany the peer-reviewed manuscript and will be deposited at osf.io/5tqcz.

*Document version v19.5.3 | Compiled 5 May 2026 | OSF: https://osf.io/5tqcz | PROSPERO: CRD420261360881 (registered, pending publication review)*
